# High Intelligence is not a Risk Factor for Mental Health Disorders

**DOI:** 10.1101/2022.05.26.22275621

**Authors:** Camille Michèle Williams, Hugo Peyre, Ghislaine Labouret, Judicael Fassaya, Adoración Guzmán García, Nicolas Gauvrit, Franck Ramus

**Affiliations:** Laboratoire de Sciences Cognitives et Psycholinguistique, Département d’Études Cognitives, École Normale Supérieure, EHESS, CNRS, PSL University, 75005, Paris, France; INSERM UMR 1141, Paris Diderot University, Paris, France; Department of Child and Adolescent Psychiatry, Robert Debré Hospital, APHP, Paris, France; Human and Artificial Cognition Lab, Ecole Pratique des Hautes Etudes, Paris, France

## Abstract

**Objective:** Studies reporting that highly intelligent individuals have more mental health disorders often have sampling bias, no or inadequate control group, or insufficient sample size. We addressed these caveats by examining the difference in the prevalence of mental health disorders between individuals with high and average general intelligence (g-factor) in the UK Biobank.

**Methods:** Participants with general intelligence (g-factor) scores standardized relative to the same-age UK population, were divided into 2 groups: a high g-factor group (g-factor 2 SD above the UK mean; N=16,137) and an average g-factor group (g-factor within 2 SD of the UK mean; N=236,273). Using self-report questionnaires and medical diagnoses, we examined group differences in prevalence across 32 phenotypes, including mental health disorders, trauma, allergies, and other traits.

**Results:** High and average g-factor groups differed across 15/32 phenotypes and did not depend on sex and/or age. Individuals with high g-factors had less general anxiety (OR=0.69) and PTSD (OR=0.67), were less neurotic (β=-0.12), less socially isolated (OR=0.85), and were less likely to have experienced childhood stressors and abuse, adulthood stressors, or catastrophic trauma (OR=0.69-0.90). They did not differ in any other mental health disorder or trait. However, they generally had more allergies (e.g., eczema; OR=1.13-1.33).

**Conclusions:** The present study provides robust evidence that highly intelligent individuals have no more mental health disorders than the average population. High intelligence even appears as a protective factor for general anxiety and PTSD.

**Key Points:** *Question:* Are high IQ individuals at increased risk of mental health disorders?

*Findings:* In the UK Biobank (N ≃ 7,266 - 252,249), highly intelligent individuals (2SD above the population mean) were less likely to suffer from general anxiety and PTSD, and no more likely to have depression, social anxiety, a drug use disorder, eating disorders, obsessive-compulsive disorder, bipolar disorder, and schizophrenia.

*Meaning:* Contrary to popular belief, high intelligence is not a risk factor for psychiatric disorders and even serves as a protective factor for general anxiety and PTSD.

## 1. Introduction

Is being intellectually gifted a strength or a handicap? Intelligence – the ability to learn, reason, and solve problems^1^ – is associated with greater physical health and longevity^2–4^ (e.g., Calvin et al., 2017; Christensen et al., 2016). And yet, some researchers report that highly intelligent individuals are at a higher risk of developing mental health and somatic disorders^5,6^. The majority of studies that report a negative effect of intelligence on mental and somatic health, however, often suffer from sampling bias, the lack of a control group, or insufficient sample size^7,8^. The present paper addresses these caveats by examining the difference in the prevalence of health and somatic disorders and other traits between the highly intelligent (2 SD above mean) and averagely intelligent (within 2 SD of the mean) individuals of the UK Biobank (N ≃ 261,500).

The notion of general intelligence (*g*) stems from the positive correlation in performance across most cognitive tests^9^. It is formally defined as the common source of variance underlying performance across a wide range of tests and is usually computed using factor analysis. Intelligence Quotient (IQ) reflects a person’s average performance across cognitive tests relative to a representative sample of the same-age national population. Henceforth, IQ and *g* will be used interchangeably and as shortcuts for general intelligence scores.

Over the last decades, intelligence has proved to be a strong predictor of education ^10,11^, occupational status, performance^12,13^, and health outcomes^14–16^. For instance, having a childhood IQ one standard deviation (SD) above the mean decreases one’s risk of accidents and developing heart, respiratory, and digestive disease by 20-25%^3,4,15,17,18^. And yet, whether intelligence also serves as a protective factor for mental health disorders is still subject to debate. While some postulate that high intelligence serves as a protective factor for several mental and somatic disorders^19–23^, others suggest that high intelligence is a risk factor for these phenotypes^5,24–29^ or that the effects of intelligence vary across mental health phenotypes^30^.

The most recent study examining the prevalence of mental health and somatic (i.e., allergies, asthma, immunodeficiencies) disorders in highly intelligent individuals reported that high IQ was a risk factor for affective disorders, neurodevelopmental disorders, and diseases related to the immune system^5^. However, the study suffers from sampling bias because participants were recruited from the American Mensa Ltd. – a society open to individuals that at some point scored in the top 2% on a verified intelligence test (N = 3,715). Since IQ tests are typically administered to children when parents or teachers notice behavioral problems or by individuals experiencing stereotypical characteristics associated with IQ, selecting individuals from a sample of individuals who actively decided to take an IQ test or become members of a highly intelligent society may exacerbate the correlation between having a high IQ and mental health disorders and/or behavioral problems^7,8^. The present paper thus aims to address these limitations.

We investigated the difference in prevalence between individuals with high (2 SD above the population mean) and average (within 2 SD from the population mean) general intelligence scores (g-factor scores^31^) in the UK Biobank across mental health disorders, somatic disorders, and certain traits. We examined group differences in the prevalence of available mental health and somatic disorders in the UK Biobank, as well as phenotypes that are thought to differ in prevalence in highly intelligent individuals, such as subjective well-being phenotypes (e.g., well-being, social isolation^32,33^), myopia^34^, chronotype^35^, and trauma^36,37^. Finally, we included sexual behaviors because studies have shown heterogeneous results concerning the relation between sexual behaviors and IQ ^38–41^. As a point of comparison, we report differences in prevalence across phenotypes between the average and low (2 SD under the population mean) g-factor groups in the exploratory analyses.

## 2. Methods

Analyses were preregistered on OSF (preregistration: https://osf.io/wcmqs/?view_only=305a3a05461c487797d2056aaf5a1460 and project with supplemental tables and files: https://osf.io/cywd6/?view_only=fa9f5091de124d96be3eb1a55a4e7f01).

### 2.1. Participants

Participants were taken from the UKBiobank, an open-access large prospective study with phenotypic, genotypic, and neuroimaging data from more than 500,000 participants recruited between 2006 and 2011 at 40 to 69 years old^42^. All participants provided informed consent (“Resources tab” at https://biobank.ctsu.ox.ac.uk/crystal/field.cgi?id=200). The UK Biobank received ethical approval from the Research Ethics Committee (reference 11/NW/0382) and the present study was conducted based on application 46 007.

UK Biobank participants were asked to complete a variety of cognitive tests upon each of their visits to the UKBiobank assessment center (http://biobank.ctsu.ox.ac.uk/crystal/label.cgi?id=100026) and online (http://biobank.ctsu.ox.ac.uk/crystal/label.cgi?id=116).

In a previous paper^31^, we calculated g-factor scores for individuals in the UK Biobank who completed at least one of the following tests: Fluid Intelligence, Matrix Pattern Completion, Tower Rearranging, Numeric Memory, Pairs Matching, Symbol Digit Substitution, Reaction Time, and Trail Making. Cognitive tests were adjusted for age and standardized using the occupational data from the 2001 census to obtain a g-Factor score relative to the general population in the UK.

We estimated the quality of the g-factor based on the combination of completed tests, and we selected participants who had a sufficiently reliable estimate of g (correlation with full g factor > 0.7; N= 261,701 participants; details in Section S1.1). Participants were excluded when there was a mismatch between the self-reported (field 31) and genetic sex (field 22001). Self-reported sex was coded -0.5 for males and 0.5 for females.

### 2.2. G-factor Groups

We created 3 g-factor groups: a high g-factor group (g-factor 2 SD above the population mean), a low g-factor group (g-factor 2 SD below the population mean), and an average g-factor group (g-factor within 2 SD from the population mean). About 90% of individuals were in the average g-factor group (236,273/261,701), 6.2% in the high g-factor group (16,137/261,701), and 3.6 % in the low g-factor group (9,291/261,701; sex differences in Section S1.2).

### 2.4. Phenotypes

To maximize the number of participants with a diagnosis, each phenotype was created from a combination of questions on diagnoses by mental health professionals, self-reported diagnoses by professionals, and probable diagnoses obtained from previous studies^43–45^ using the UK Biobank questionnaires (Tables SA, Section 1.3). We examined 32 phenotypes using one or several binary, ordinal, or continuous variables (Table 1; Table SB1-B2).

**Table 1.**
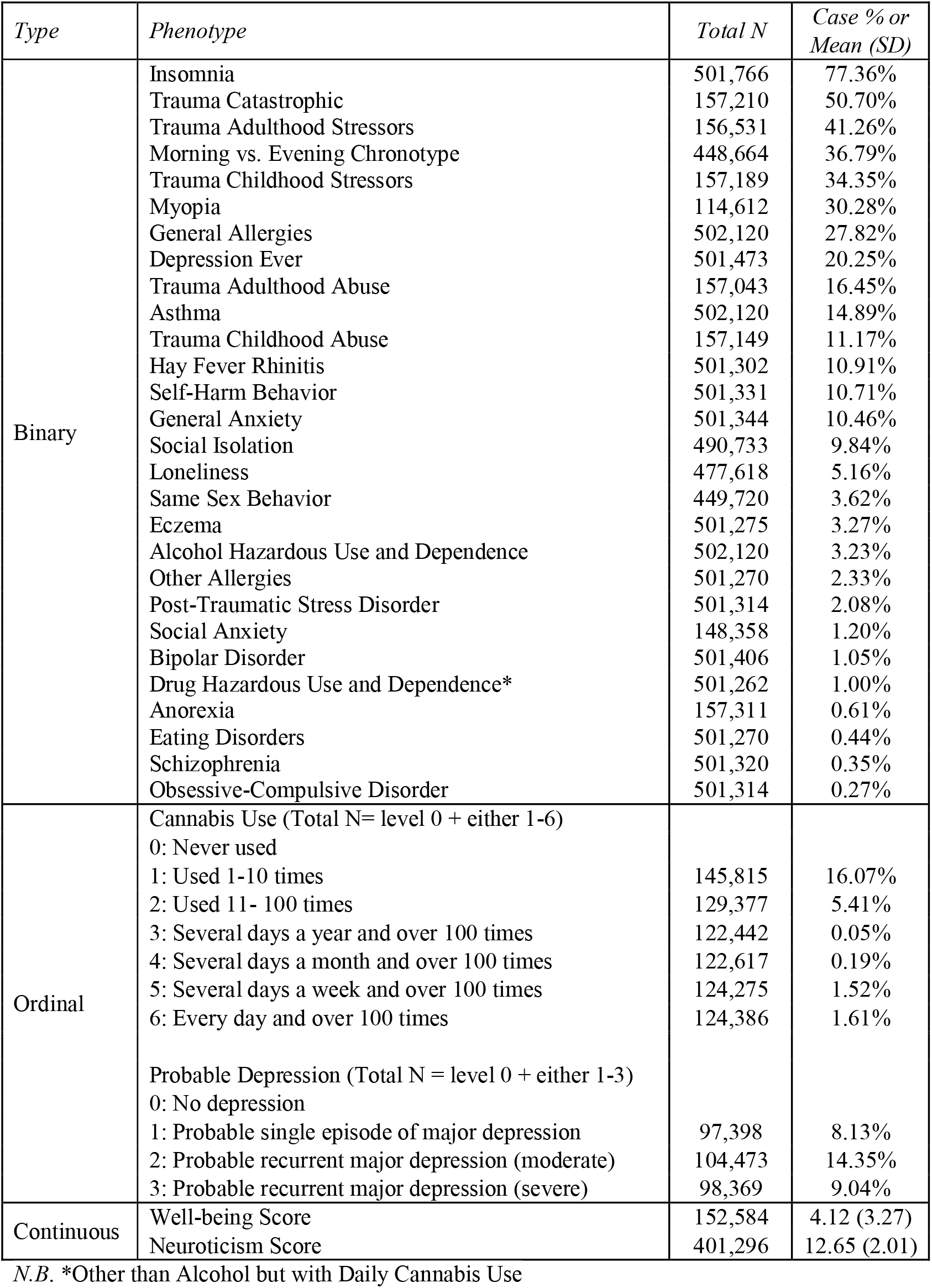
Phenotypic Prevalence in the UK Biobank

| *Type* | *Phenotype* | *Total N* | *Case % or Mean (SD)* |
| --- | --- | --- | --- |
| Binary | Insomnia | 501,766 | 77.36% |
|  | Trauma Catastrophic | 157,210 | 50.70% |
|  | Trauma Adulthood Stressors | 156,531 | 41.26% |
|  | Morning vs. Evening Chronotype | 448,664 | 36.79% |
|  | Trauma Childhood Stressors | 157,189 | 34.35% |
|  | Myopia | 114,612 | 30.28% |
|  | General Allergies | 502,120 | 27.82% |
|  | Depression Ever | 501,473 | 20.25% |
|  | Trauma Adulthood Abuse | 157,043 | 16.45% |
|  | Asthma | 502,120 | 14.89% |
|  | Trauma Childhood Abuse | 157,149 | 11.17% |
|  | Hay Fever Rhinitis | 501,302 | 10.91% |
|  | Self-Harm Behavior | 501,331 | 10.71% |
|  | General Anxiety | 501,344 | 10.46% |
|  | Social Isolation | 490,733 | 9.84% |
|  | Loneliness | 477,618 | 5.16% |
|  | Same Sex Behavior | 449,720 | 3.62% |
|  | Eczema | 501,275 | 3.27% |
|  | Alcohol Hazardous Use and Dependence | 502,120 | 3.23% |
|  | Other Allergies | 501,270 | 2.33% |
|  | Post-Traumatic Stress Disorder | 501,314 | 2.08% |
|  | Social Anxiety | 148,358 | 1.20% |
|  | Bipolar Disorder | 501,406 | 1.05% |
|  | Drug Hazardous Use and Dependence* | 501,262 | 1.00% |
|  | Anorexia | 157,311 | 0.61% |
|  | Eating Disorders | 501,270 | 0.44% |
|  | Schizophrenia | 501,320 | 0.35% |
|  | Obsessive-Compulsive Disorder | 501,314 | 0.27% |
| Ordinal | Cannabis Use (Total N= level 0 + either 1-6)<br>0: Never used |  |  |
|  | 1: Used 1-10 times | 145,815 | 16.07% |
|  | 2: Used 11- 100 times | 129,377 | 5.41% |
|  | 3: Several days a year and over 100 times | 122,442 | 0.05% |
|  | 4: Several days a month and over 100 times | 122,617 | 0.19% |
|  | 5: Several days a week and over 100 times | 124,275 | 1.52% |
|  | 6: Every day and over 100 times | 124,386 | 1.61% |
|  | Probable Depression (Total N = level 0 + either 1-3)<br>0: No depression |  |  |
|  | 1: Probable single episode of major depression | 97,398 | 8.13% |
|  | 2: Probable recurrent major depression (moderate) | 104,473 | 14.35% |
|  | 3: Probable recurrent major depression (severe) | 98,369 | 9.04% |
| Continuous | Well-being Score | 152,584 | 4.12 (3.27) |
|  | Neuroticism Score | 401,296 | 12.65 (2.01) |

*N.B.* \*Other than Alcohol but with Daily Cannabis Use

### 2.5. Age

As the age of onset of disorders was not available for all participants and/or measures, we took into account the maximum age at which an individual provided the most recent measure of a disorder to approximate lifetime prevalence (Section S1.4).

### 2.6. Statistical Analyses

To reduce the number of statistical tests performed when examining group differences in each phenotype, we first examined age and sex effects and interactions on each phenotype (Section S1.5.1). If age and sex’s main effects or interactions did not significantly predict a phenotype (p > 0.05), they were excluded from the g-factor group analyses. Equation 1 corresponds to the model with all possible predictors.

To be included in the group comparison of a phenotype, participants had to have a *g* factor measure. They had to have answered all of the questions used to create that phenotype, except when the same question was asked several times (Section S1.5.2).

We used logistic regression for binary phenotypes, multinomial regressions for ordinal phenotypes, and linear regressions for continuous phenotypes. Age was mean-centered in ordinal and binary regressions, while age and continuous phenotypes were men-centered and divided by 1sd in linear regressions to report standardized betas.

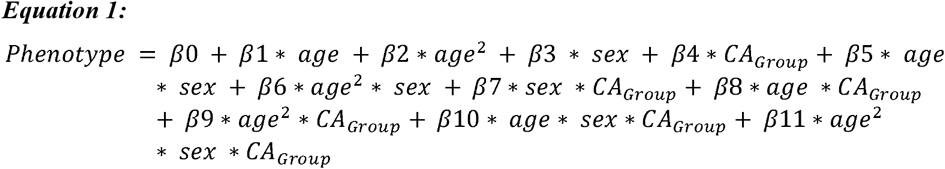

We report results with the g-factor group predictor that survived a Bonferroni correction of 0.05 divided by the number of coefficients in the equation with the g-factor group term times the number of coefficients of interest times the number of investigated phenotypes (e.g., 0.05/ (6*32) for equation 2). Since the analyses on the low g-factor group were exploratory, we applied the same multiple comparison correction as for the high g-factor group (e.g., 0.05/ (6*32) for equation 2).

## 3. Results

### 3.1. Differences in the Prevalence of Phenotypes between Participants with High and Average CA

Sex and age effects across phenotypes are described in Supplemental Section 2.1. and in Supplemental Tables S3. The prevalence of each disorder by g-factor group is available in Supplemental Tables S4 and regression results are reported in Supplemental Tables S5.

Across the 32 phenotypes, the prevalence differed between the high g-factor and average g-factor groups in 15 phenotypes (47%) and between the low g-factor and average g-factor groups in 12 phenotypes (38%; Figure 1). Phenotypic differences between the Low and Average g-factor are discussed in Supplemental Section 2.3. There were no significant interactions in the analyses comparing the average to the high g-factor groups. Low-powered phenotypes are discussed in Supplemental Section 2.4.

**Figure 1.**
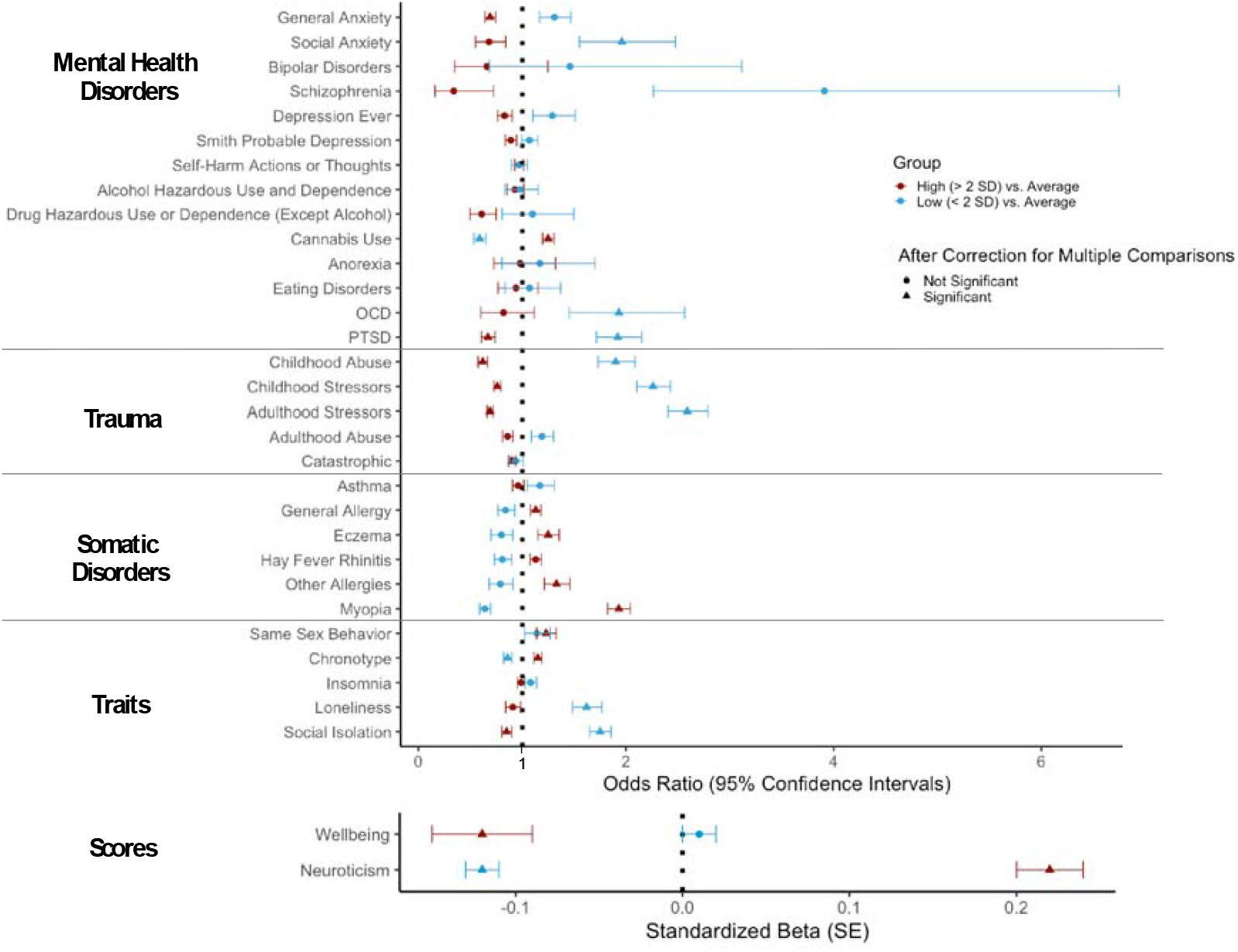
Group Differences in Prevalence between High and Average and Low and Average g-factor Groups across Phenotypes and Scores. OCD: Obsessive-Compulsive Disorder; PTSD: Post-Traumatic Stress Disorder. Correction for multiple comparisons varies by phenotype. See supplemental tables standard error (SE) for p-value thresholds for multiple comparison corrections. High g-factor: participants with a g-factor score 2SD above the mean. Low g-factor: participants with a g-factor score 2SD under the mean. Average g-factor: participants with a g-factor score between + or – 2SD from the mean.

#### 3.1.1. Mental Health Disorders

Compared to individuals in the average g-factor group, there was a 33% decrease in the odds of suffering from Post-Traumatic Stress Disorder (OR = 0.67) and a 31% decrease in the odds of having general anxiety (OR = 0.69) in the high g-factor group (Table 2). There was no significant difference across other mental health disorders (Figure 1).

**Table 2.**
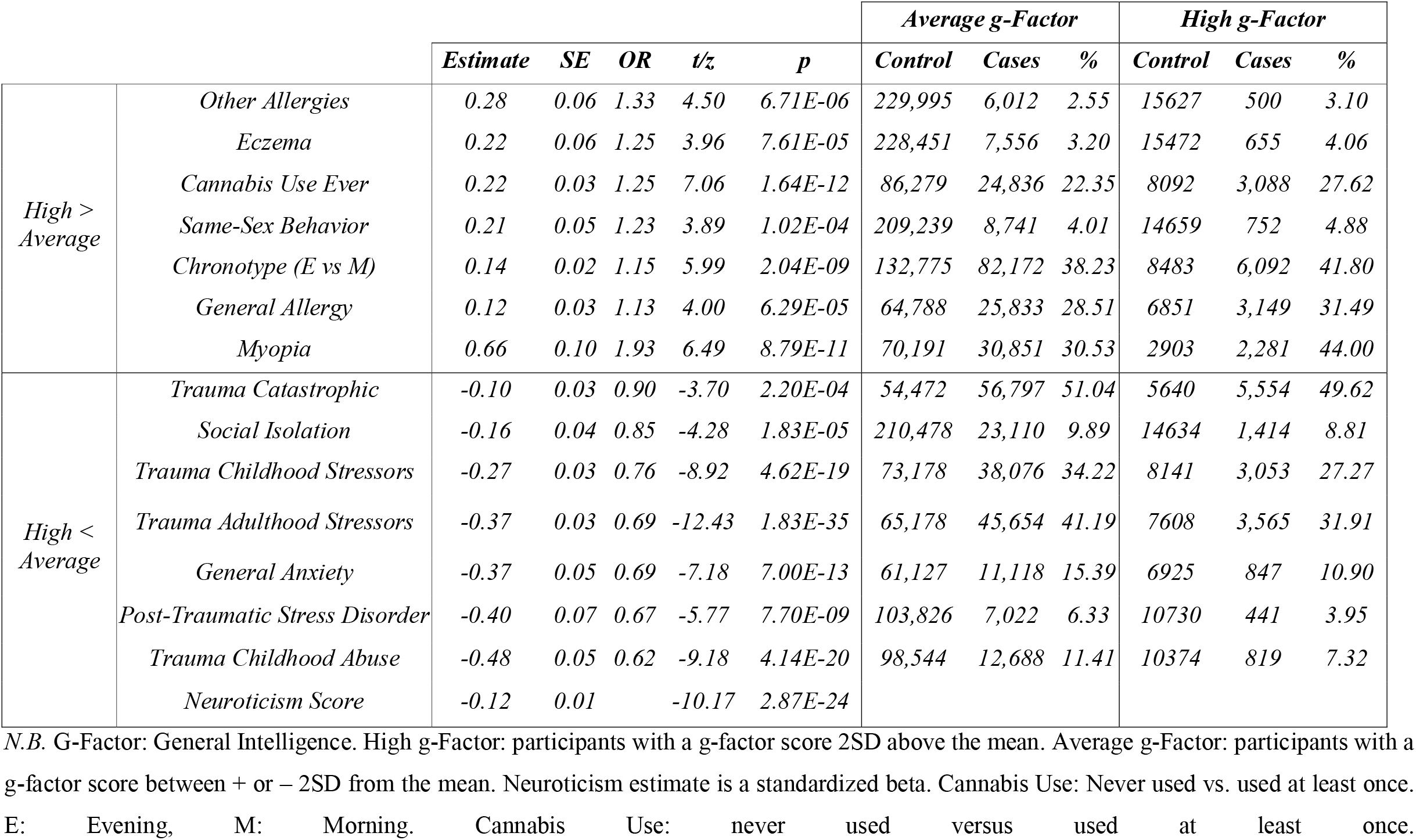
Phenotypes that differ in Prevalence between Average and High General Intelligence Groups.

#### 3.1.2. Somatic Phenotypes

Compared to individuals in the average g-factor group, the odds of having some type of allergy increased by 13% for individuals in the high g-factor group (OR=1.13). This was explained by their greater propensity to having eczema and other allergies (e.g., food; respectively 1.25 and 1.33 times more likely), which were included in the general allergy diagnosis (Table 2). The odds of being myopic increased by 93% in the high g-factor group (OR = 1.93) and this remained significant when controlling for educational attainment (OR = 1.75; Supplemental Section 2.2).

#### 3.1.3. Trauma

Compared to individuals in the average g-factor group, the odds of experiencing catastrophic trauma, adulthood stressors, childhood abuse, and childhood stressors decreased by 10% (OR=0.90), 31% (OR = 0.69), 38% (OR = 0.62), and 24% (OR = 0.76; Table 2) in the high g-factor group, respectively.

#### 3.1.4. Traits

Compared to individuals in the average g-factor group, the odds of feeling more socially isolated decreased by 15% in the high g-factor group (OR=0.85), whereas the odds of having an evening-like chronotype, ever engaging in same-sex behavior, and ever using cannabis increased by 15% (OR = 1.15), 23% (OR = 1.23), and 25% (OR=1.25) respectively in the high g-factor group. Figure S4 shows that there are more individuals with a higher-than-average g-factor that have used cannabis 1 to 100 times and that over 100 times there are no group differences. The high g-factor group had a lower neuroticism score than individuals in the average g-factor group (β = -0.12; Table 2).

### 3.2. Phenotypes with group differences between both the high and average and the low and average g-factor groups

Childhood stressors, childhood abuse, adulthood stressors, PTSD, and social isolation were more prevalent in the low g-factor group compared to the average g-factor group (Table S2) and were more prevalent in the average g-factor group compared to the high g-factor group (Table 2), suggesting that the prevalence of these phenotypes decreases with an increasing g-factor. The low g-factor group had a higher neuroticism score than the average g-factor group, which had a higher neuroticism score than the high g-factor group. The odds of ever trying cannabis and having an evening-like chronotype respectively decreased by 41% (OR = 0.59) and 14% (OR = 0.86) in the low g-factor group compared to the average g-factor group.

## 4. Discussion

We examined differences in the prevalence of mental health disorders, somatic and certain traits between individuals with high (2 SD above mean) and average g-factor scores (within 2 SD of the mean) in the UK Biobank (N ≃ 7,266 - 252,249). We contrasted these results with differences observed between individuals with low and average g-factor scores.

We found that the high g-factor group did not have more mental health disorders than the average g-factor group in general and that they were less likely to have general anxiety and PTSD. Individuals with higher intelligence were also less likely to have experienced trauma and stressors, except for adulthood abuse, which may be part of the explanation for the previous finding. The high g-factor group was also more neurotic and felt less socially isolated. In contrast, the low g-factor group was more neurotic, felt more socially isolated, and had a greater prevalence of trauma, stressors, and PTSD than the average g-factor group, suggesting that the prevalence of these phenotypes decreases with increasing intelligence. Among the few somatic disorders that were examined, we found that individuals with high intelligence were more myopic and had more allergies, although they had a lower prevalence of hay fever rhinitis, and asthma. Individuals with high intelligence were also more likely to present certain traits, such as having an afternoon-evening chronotype, ever tried cannabis, and ever engaged in same-sex behavior, whereas the low g-factor group was less likely to have ever tried cannabis and engaged in same-sex behavior than the average g-factor group. There were no differences between groups in the prevalence of insomnia.

The result of the present study contradicts several studies that reported an increased risk for various psychiatric disorders in individuals with high intelligence^5,24,26,28,46,47^. As noted in the introduction, these studies were generally based on small samples and suffered from major sampling bias or a lack of a control group^7,8^. Our results suggest that high intelligence is not a risk factor for psychiatric disorders and even a protective factor for general anxiety. We find that increasing intelligence is associated with a decrease in trauma exposure, and consequently PTSD. This is consistent with previous findings^48^ and with the association of childhood trauma with lower intelligence^49^.

With regards to somatic disorders, we replicate the increased risk of allergies in individuals with high intelligence^5,47,50^. One possible explanation for this association is that allergies and intelligence share neural correlates^51^. Another possibility is that more intelligent individuals with a higher g-factor live in more urban areas^52^, where allergies are more prevalent^53^, or that individuals with high intelligence are more aware of allergic symptoms and have better access to health care. However, the prevalence between groups did not differ across all allergies (e.g., Asthma, Hay Fever Rhinitis).

In line with a previous literature review^34^, the risk of myopia was greater for individuals with high intelligence. While near-work activities (e.g., reading and computer use) seem to be a risk factor for myopia^54,55^, this association appears to be distinct from that of higher intelligence and education level^29,54^. Although additional years of education contribute to an increase in the risk of developing myopia^56^, most of the evidence points toward shared genetic factors between intelligence and myopia^29^, which is consistent with our observation that the risk of myopia associated with a high g-factor only slightly decreased when adjusting for educational attainment.

Our results indicate more afternoon-evening chronotypes in individuals with high intelligence than in individuals with average intelligence, which could be explained by differences in the work schedules of the different g-factor groups^35^. In line with previous studies, we find that individuals with high intelligence are more likely to ever have engaged in same-sex behavior^38–41^. We note that this measure may not reflect sexual orientation, but sexual exploration. We also found an association between ever trying cannabis and intelligence, but this was only true when looking at individuals who consumed cannabis less than 101 times in their lifetime, not for more intensive consumption. Therefore, this measure may reflect a tendency to explore rather than a substance abuse disorder. One possibility is that individuals with higher intelligence, which is positively correlated with the “Openness to Experience” personality trait (r = 0.30^57^), may be more likely to seek out new experiences and explore alternative behaviors than the average.

First, as most neurodevelopmental and some psychotic disorders were not available or had too few cases in the UK Biobank, our results do not allow us to conclude on these psychiatric disorders. Second, UK Biobank has a “healthy volunteer” selection bias^58^. Therefore, the UK-Biobank sample has fewer psychiatric disorders^43,58^ and a higher g-factor score than the general population^31^. Although the prevalence of psychiatric disorders and traits differ from the general population, this should not affect the validity of the group comparisons. Third, the prevalence of psychiatric disorders and traits between g-factor groups may differ across the lifespan. However, here, we were interested in lifetime prevalence, which makes the UK Biobank, a prospective aging study, a good candidate for the question at hand.

High intelligence is not a risk factor for psychiatric disorders and is a protective factor for general anxiety and PTSD. Our results reinforce the idea that higher intelligence is an advantage. This does not imply that general intelligence is irrelevant for psychiatric evaluation: indeed, it may affect the presentation of symptoms, and the available resources for recovery.

## Data Availability

This research has been conducted using data from UK Biobank, a major biomedical database (http://www.ukbiobank.ac.uk/). Restrictions apply to the availability of these data, which were used under license for this study: application 46007.
Preregistration and code are available on OSF: https://osf.io/cywd6/?view_only=fa9f5091de124d96be3eb1a55a4e7f01

https://osf.io/cywd6/?view_only=fa9f5091de124d96be3eb1a55a4e7f01

## Acknowledgments

This work received support under the program “Investissements d’Avenir” launched by the French Government and implemented by l’Agence Nationale de la recherche (ANR) with the references ANR-17-EURE-0017 and ANR-10-IDEX-0001-02 PSL. This research has been conducted using the UK Biobank Resource. Declarations of interest: none.

