## Supplemental Information for "High Intelligence is not a Risk Factor for Mental Health Disorders"

### 1. Methods

Supplemental tables and files are available on OSF along with the code and preregistration: https://osf.io/cywd6/?view_only=fa9f5091de124d96be3eb1a55a4e7f01

#### 1.1. G-Factor Quality

In the sample that completed all cognitive tests, we correlated their g-factor scores calculated from all tests with their g-factor scores calculated from each possible combination of completed tests. In the sample that completed at least one of the 8 cognitive tests, we selected individuals who completed a combination of tests that would allow for a minimum g-factor score correlation of 0.70 (N= 261,701 participants) to obtain a sufficient number of participants to create g-factor groups with reliable G-factor scores. As discussed by Williams and colleagues (2022)^1^, the g-factor score should not be influenced by cognitive decline since tests show reasonable stability over time^2^ and declines in cognitive test scores before 65 years are small^3^.

#### 1.2. Sex Differences by G-factor Group

There were more males in the high g-factor compared to the average g-factor group (Table 1).

**Table 1. Prevalence of Males and Females across G-factor Groups.**

| **Sex/Group** | **Average g-factor** | | **High g-factor** | | **Low g-factor** | |
| --- | --- | --- | --- | --- | --- | --- |
| Both | 236,273 | 100.00% | 16,137 | 100.00% | 5,261 | 100.00% |
| Female | 130,613 | 55.28% | 7,017 | 43.48% | 4,983 | 56.63% |
| Male | 105,660 | 44.72% | 9,120 | 56.52% | 4,030 | 43.38% |
| 𝜒2 Test |  |  | 𝜒2 (1) = 847.30, p < 2.2e-16 | | 𝜒2 (1) = 6.48, p = 0.011 | |

*N.B.* g-factor: general intelligence factor. Chi-square test: Sex difference between High or Low g-factor group and Average g-factor Group.

#### 1.3. Phenotypes

We initially preregistered that we would examine 30 binary, ordinal, or continuous diagnoses for 17 phenotypes (Well-being: 1, Neuroticism: 1, Anxiety: 3, Trauma: 3, Depression: 4, Self-Harm: 1, Social Isolation: 1, Loneliness: 1, Bipolar Disorder: 1, Sleep Behaviors: 2, Eating Disorders: 2, Drug use and dependence: 3, Myopia:1, Asthma: 1, Allergy: 1, Sexual Behaviors: 3, Schizophrenia, delusion and psychotic experiences: 1, and Obsessive-Compulsive Disorder: 1).

To evaluate the relationship between the g-factor group and disorder severity, we planned on analyzing continuous phenotypes in addition to binary and ordinal ones for the same disorder. However, we were forced to exclude the CIDI, the PHQ9, the GAD scores, and the number of different and same-sex sexual partners because the regressions methods used did not fit the data well (linear, Poisson, quasi-Poisson, and negative binomial). Zero-inflation regressions did not converge due to the presence of an interaction with quadratic age.

Linear regression was not appropriate for some of the continuous phenotypes that we planned to investigate. Specifically, we did not find an appropriate regression method to model the CIDI severity distribution (Figure 1) and failed to fit Poisson, negative binomial, and or zero-inflated regression models on the remaining phenotypes (Figure 2). When the Poisson model could not model the overdispersion of the data, we ran quasi-Poisson and negative binomial regressions. The negative binomial regression sometimes led to under dispersion or did not appear to fix the dispersion problem. Since the quasi-Poisson and negative binomial methods do not have the same likelihood function, we failed to identify the best fitting model. Finally, the zero-inflated model did not converge due to the presence of the age2 by sex interaction. Please see the sex by age markdowns in Supplementary Files S1-6 for each of these analyses on each of the phenotypes in Table 2.

Instead, we used the CIDI, PHQ9, and GAD cut-offs from Davis and colleagues' (2020) study. However, because we wanted an alcohol diagnosis that indicated severe hazardous drinking or dependence, we used a threshold of 15 as indicated on the audit screening website [here](https://auditscreen.org/about/scoring-audit) instead of 8 as previously done^4^. We examined the specificity and sensitivity of the AUDIT score in light of the ICD10 diagnosis for harmful use and dependence (Supplemental File S7).

We further divided the trauma phenotype into more specific traumatic phenotypes after observing that a large number of participants were categorized as having a traumatic experience. Finally, instead of having one substance abuse disorder phenotype, we created a phenotype for alcohol addiction and one for all other substance dependence.

**Table 2. Distribution of Continuous Phenotype Scores and Counts**

| **Phenotype** | **Min** | **Q1** | **Median** | **Mean** | **Q3** | **Max** |
| --- | --- | --- | --- | --- | --- | --- |
| AUDIT Score | 0.0 | 2.0 | 4.0 | 4.9 | 7.0 | 40.0 |
| GAD Severity | 0.0 | 0.0 | 0.0 | 2.2 | 3.0 | 21.0 |
| PHQ9 Severity | 0.0 | 0.0 | 2.0 | 2.8 | 4.0 | 27.0 |
| CIDI Severity | 0.0 | 0.0 | 3.0 | 3.2 | 6.0 | 8.0 |
| N Same-Sex Partners | 0.0 | 1.00 | 2.0 | 27.3 | 5.0 | 15000 |
| N Different Sex Partners | 0.0 | 2.0 | 6.0 | 36.5 | 20.0 | 15000 |
| N Sex Partners | 0.0 | 3.0 | 9.0 | 63.8 | 23.0 | 30000 |

*N.B.* N: Number of individuals. AUDIT: Alcohol Use Disorders Identification Test. GAD: Generalized Anxiety Disorder. PHQ9: Patient Health Questionnaire. CIDI: Composite International Diagnostic Interview. N: Number.

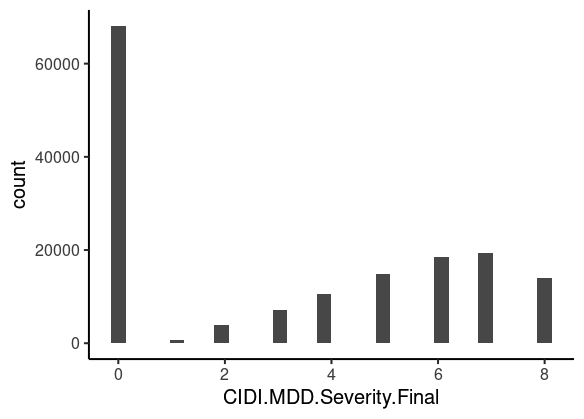

***Figure 1.*** Distribution of the CIDI Major Depression Disorder (MDD).

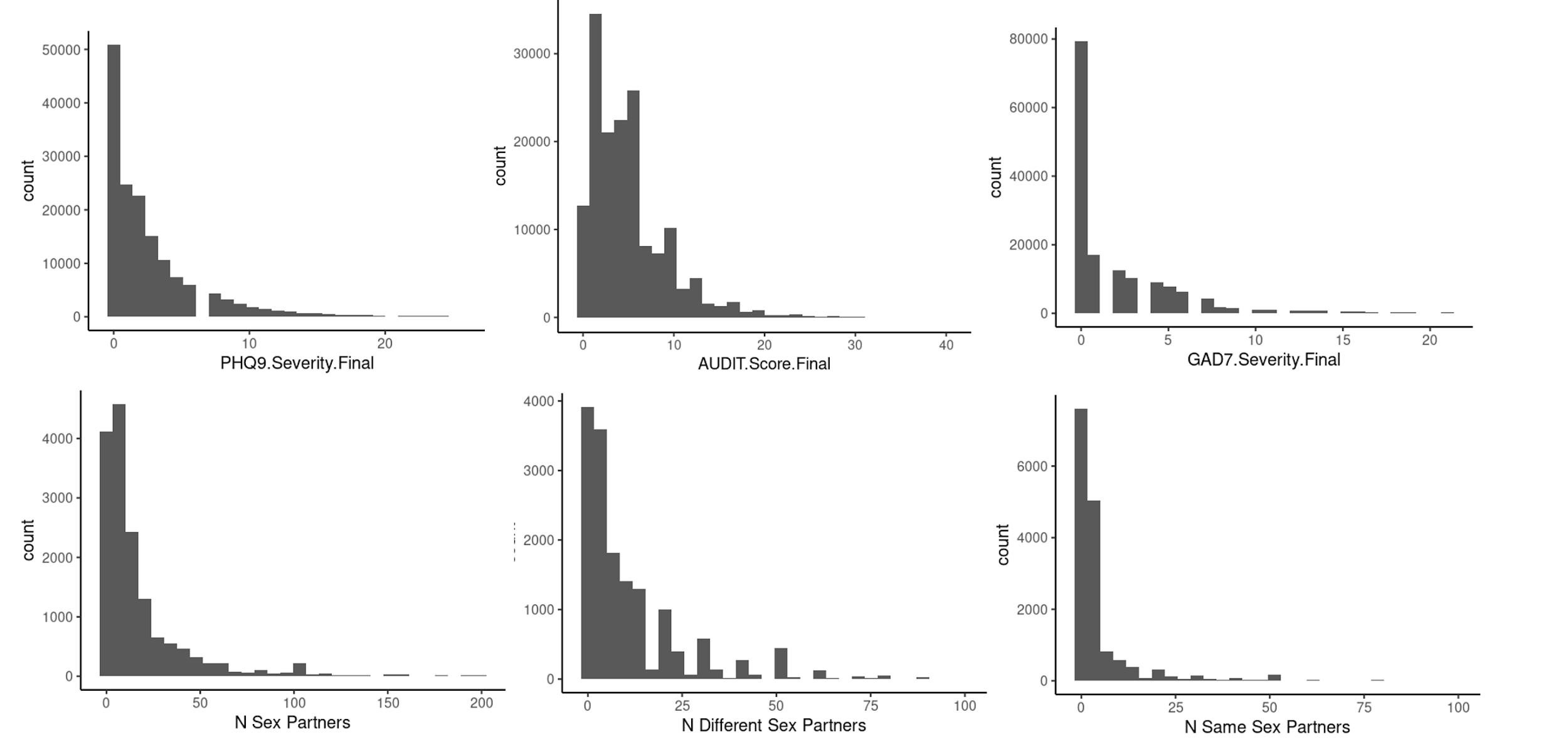

***Figure 2.*** Distribution of the PHQ9, the Alcohol Use Disorder Identification Test (AUDIT), and the GAD7 Scores and the number of total sexual partners (< 200), different sex partners (<100), and same sex partners (<100).

#### 1.4. Age

The age at the center was calculated using the date at which they attended the assessment center (field 53) and their birth month (field 52) and year (field 34). The age at which they were diagnosed was calculated using the date at which they received the inpatient diagnosis (ICD9 and ICD10: 41281 and 41280, respectively) and their birth month (field 52) and year (field 34). The online follow-up age was calculated as done by Davis and colleagues (2020), who created the UK Biobank Online MHQ, by using the age when attending the center (field 21003) and the date of completing the MHQ (field 20400). The age measures used to calculate max-age for each phenotype are indicated in the Tables SA1-17.

#### 1.5. Statistical Analyses

##### 1.5.1. Phenotypic Age & Sex Effects

To investigate general age and sex effects and interactions across phenotypes, we examined age and sex effects and interactions on a larger UK Biobank sample than the one included in the g-factor analyses. Participants were included in the sex and age analyses of a phenotype if they answered one of the questions used to establish that phenotype. If participants responded as having a phenotype across any of the questions used to create that phenotype, they were marked as having the phenotype, otherwise, they were marked as not having the phenotype.

We ran equation 1 to identify sex and age (linear and quadratic) effects and interactions on each phenotype. Logistic regressions were used for binary phenotypes, ordinal regressions for the Smith Probable Depression^5^ and the Cannabis Use phenotypes, and linear regressions for the well-being and neuroticism scores. Age was mean-centered.

If the main effects or interactions of the Age and Sex Analyses were not significant (p > 0.05), the main effects or interactions were removed from the g-factor group analyses. However, the main effects were maintained in the model if they were significant or if their interaction was significant.

***Equation 1:***

$Phenotype = \beta0 + \beta1* age + \beta2*{age}^{2} + \beta3 * sex + \beta4* age*sex + \beta5*{age}^{2}*sex$

##### 1.5.2. Group Analyses

If the question was asked several times, they were recorded as having the phenotype if they reported having the phenotype in any of the questions used to identify the phenotype. If participants responded as having the phenotype across any of the questions, they were marked as having the phenotype, otherwise, they were marked as not having the phenotype. However, the main effects were maintained in the model if they were significant or if their interaction was significant.

### 2. Results

#### 2.1. Sex and age effects and interactions

Sex and age effects varied across phenotypes (p < 0.05, Table S3). In brief, all sex and age effects and interactions were significant for the neuroticism score, social isolation, Loneliness, Eczema, Hay Fever Rhinitis, Insomnia, Chronotype Catastrophic Trauma, and Cannabis Use (Ordinal). Sex and age effects and interactions were significant except for the sex by linear age interaction for the General Allergy and Lifetime Depression Phenotypes. Sex and age effects and interactions were significant except for the sex by quadratic age interaction for the Asthma, Myopia, Childhood Abuse, Adult Abuse, Adult Stressors, General Anxiety, Self-Harm, Alcohol Hazardous Use or Dependence, and Smith’s Probable Depression (Ordinal) Phenotypes. Sex was not a significant predictor of Well-Being, Bipolar Disorder, or Social Anxiety.

#### 2.2. High vs. Average g-factor

Individuals with a High g-factor were less neurotic than individuals with an average g-factor (β= -0.12, SE= 0.01, p < 2.87E-24; Figure 3) and were more than 1.25 times more likely to have consumed cannabis at least once (i.e., comparing 0 to 1 and above; Figure 4).

In light of the positive association between education on myopia^6^, we ran exploratory analyses to examine whether the prevalence of myopia differed between high and average g-factor groups by controlling for education with Educational Qualification (data field 6138). We coded College or University as 2, A levels as 1, and GCSE, CSE, and NVQs as 0. We found that once we added this variable as a covariate the OR decreased from 1.93 to 1.75 (p = 1.21e-04) and was still significant after multiple comparison correction (p = 0.05/(5*32) = 3.13e-04).

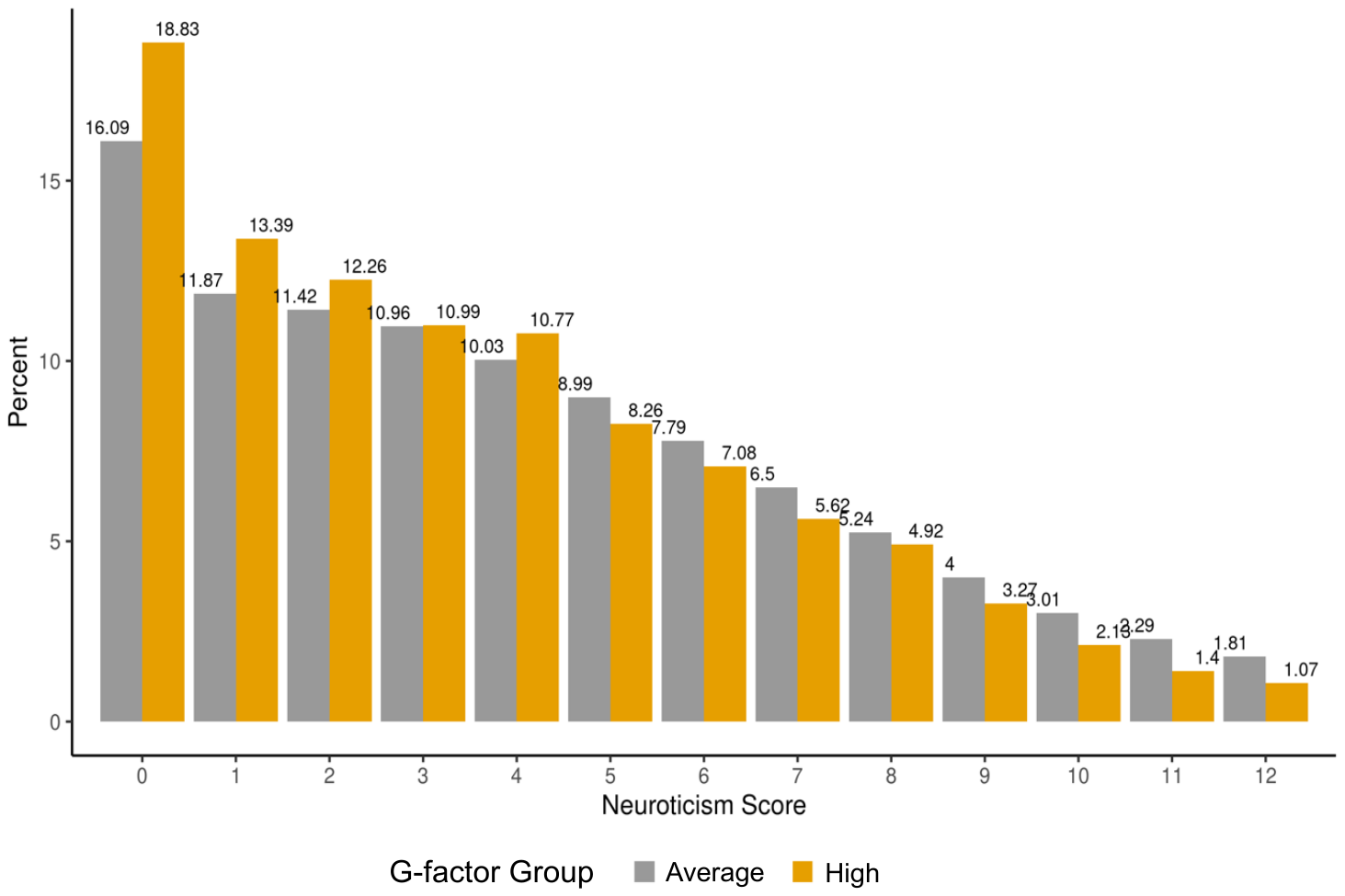

***Figure 3. Prevalence of High and Average g-factor Groups at each Neuroticism Score.***

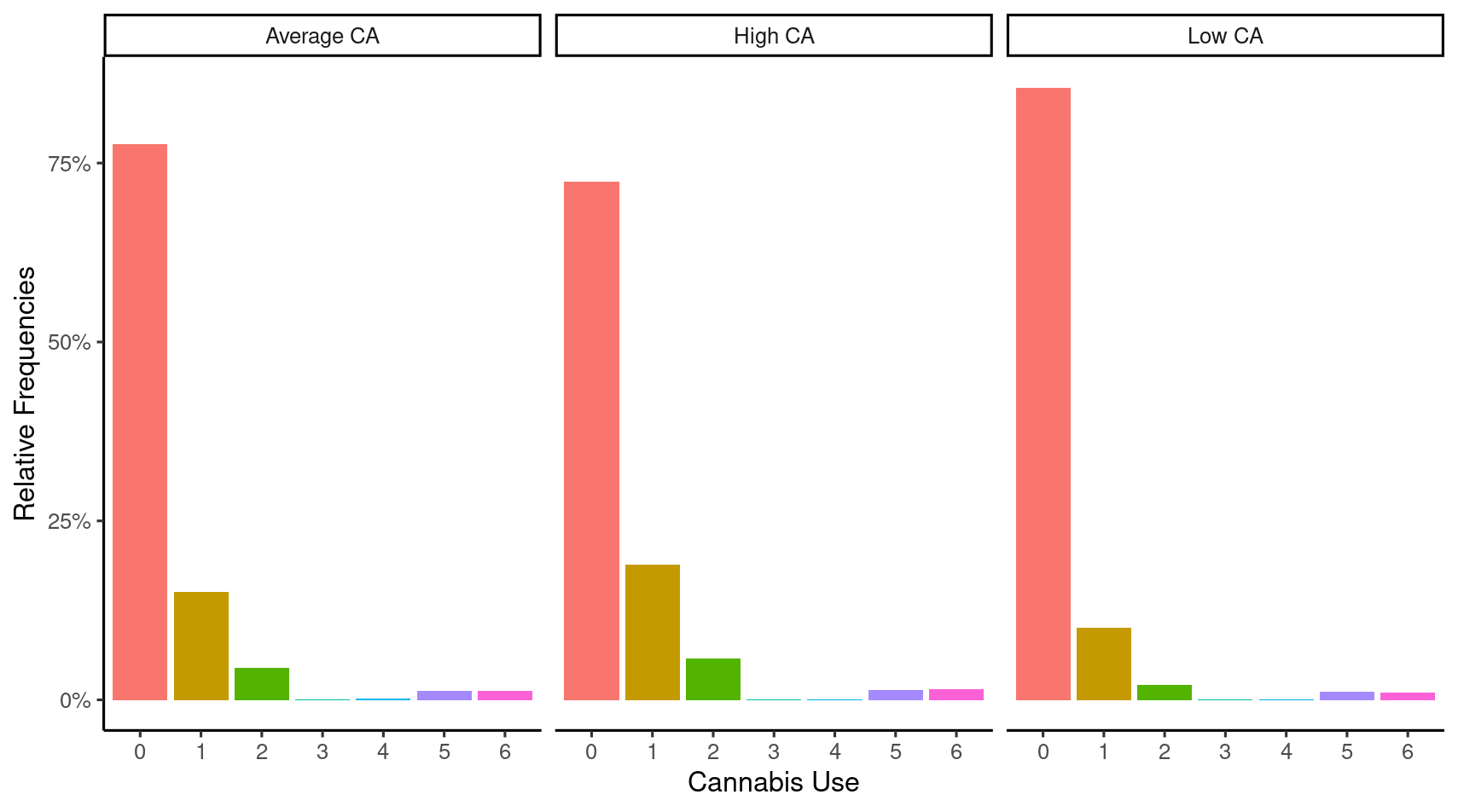

***Figure 4. Prevalence of High, Average, and Low g-factor Groups at each level of Cannabis Use.*** *G-factor: general intelligence factor. Group 0 (0 times): 20453 = 0; Group 1 (1-10 times): 20453 = 1 or 20453 = 2; Group 2 (11-100 times) : 20453 = 3; Group 3 (Several days a year, > 100 times): 20453 = 4 & 20454 = 1 Group 4 (Several days a month, > 100 times): 20453 = 4 & 20454 = 2 Group 5 (Several days a week, > 100 times) : 20453 = 4 & 20454 = 3 Group 6 (Every day, > 100 times) : 20453 = 4& 20454 = 4.*

#### 2.3. Low vs. Average g-factor Group

Across 32 phenotypes, the low g-factor differed from the average g-factor in 12 phenotypes (Table 3).

##### 2.3.1. Mental Health Disorders

Compared to individuals in the average g-factor group, the odds in the low g-factor group of having OCD, social anxiety, and PTSD increased by 170% (OR = 2.70), 96% (OR=1.96) and 92% (OR = 1.92), respectively.

##### 2.3.2. Trauma

In contrast to individuals in the average g-factor group, the odds of experiencing adulthood stressors, childhood stressors, and childhood abuse increased by 159% (OR = 2.59), 126% (OR=2.26) and 90% (OR = 1.90) in the low g-factor group, respectively.

##### 2.3.4. Traits

Individuals in the low g-factor group were respectively more likely to feel socially isolated (OR = 1.75) and lonely (OR = 1.62) compared to individuals in the average g-factor group. They were also less likely to have an afternoon-evening chronotype (OR = 0.86) and to have ever consumed cannabis (OR = 0.59). The low g-factor group had higher neuroticism scores (β = 0.22) compared to the average g-factor group. The low g-factor group also had lower well-being scores (β = -0.12) compared to the average g-factor group, but this main effect was driven by a significant Age by g-factor Group interaction for well-being (b = 0.07, SE = 0.02, p = 2.40e-4; Supplemental File S8). Wellbeing scores increased more with age in individuals with low g-factor (b = 0.12, SE = 0.02, OR = 1.13, p = 8.71e-11) than individuals with an average g-factor (b = 0.07, SE = 0.003, OR = 1.07, p = 2.17e-110; Supplemental File S8).

**Table 3 . Prevalence of Phenotype between Average and Low G-factor Groups**

|  |  |  |  |  |  |  | ***Average g-factor*** | | | ***Low g-factor*** | | |
| --- | --- | --- | --- | --- | --- | --- | --- | --- | --- | --- | --- | --- |
|  |  | ***Estimate*** | ***SE*** | ***OR*** | ***t/z*** | ***p*** | ***Control*** | ***Cases*** | ***%*** | ***Control*** | ***Cases*** | ***%*** |
| *Low > Average* | *Obsessive-Compulsive Disorder* | *0.99* | *0.16* | *2.70* | *6.34* | *2.23E-10* | *110570* | *694* | *0.62* | *3073* | *52* | *1.66* |
|  | *Trauma Adulthood Stressors* | *0.95* | *0.05* | *2.59* | *17.36* | *1.73E-67* | *65178* | *45654* | *41.19* | *1133* | *1948* | *63.23* |
|  | *Trauma Childhood Stressors* | *0.82* | *0.05* | *2.26* | *15.51* | *2.76E-54* | *73178* | *38076* | *34.22* | *1474* | *1650* | *52.82* |
|  | *Social Anxiety* | *0.67* | *0.14* | *1.96* | *4.80* | *1.58E-06* | *110638* | *675* | *0.61* | *3099* | *29* | *0.93* |
|  | *Post-Traumatic Stress Disorder* | *0.65* | *0.09* | *1.92* | *7.58* | *3.49E-14* | *103826* | *7022* | *6.33* | *2734* | *357* | *11.55* |
|  | *Trauma Childhood Abuse* | *0.64* | *0.07* | *1.90* | *9.46* | *3.00E-21* | *98544* | *12688* | *11.41* | *2567* | *553* | *17.72* |
|  | *Social Isolation* | *0.56* | *0.04* | *1.75* | *13.62* | *2.93E-42* | *210478* | *23110* | *9.89* | *7550* | *1363* | *15.29* |
|  | *Loneliness* | *0.48* | *0.06* | *1.62* | *8.09* | *6.13E-16* | *216552* | *10904* | *4.79* | *7796* | *579* | *6.91* |
|  | *Neuroticism Score* | *0.22* | *0.02* |  | *11.87* | *1.65E-32* |  |  |  |  |  |  |
| *Low < Average* | *Chronotype (E vs M)* | *-0.15* | *0.03* | *0.86* | *-4.67* | *3.08E-06* | *132775* | *82172* | *38.23* | *5518* | *2849* | *34.05* |
|  | *Cannabis Use* | *-0.52* | *0.07* | *0.59* | *-7.24* | *4.43E-13* | *86279* | *24836* | *22.35* | *2669* | *451* | *14.46* |
|  | *Wellbeing Score* | *-0.12* | *0.03* |  | *-4.48* | *7.45E-06* |  |  |  |  |  |  |

*N.B.* G-factor: general intelligence score. With NA: With missing values, participants answered at least one of the questions used to create a phenotype. Complete except Self-Report (SR) Instance 0-3: Participants responded to all questions at least once. E: Evening, M: Morning. Cannabis Use: never used versus used at least once.
