## Supplementary material for "High Intelligence is not a Risk Factor for Mental Health Disorders": Supplemental_File_S1_AUDIT_Age_Sex.html


### AUDIT\_Age\_Sex

### AUDIT, recent depression score - Age and Sex Effects

#### AUDIT Score Distribution

##### Distribution of Raw AUDIT Score

```
hist(Depression_DF_no_NA$AUDIT.Score.Final)
```

##### Distribution of log transformed AUDIT Score

```
Depression_DF_no_NA$AUDIT.Score.Final.log <- log(Depression_DF_no_NA$AUDIT.Score.Final)
hist(log(Depression_DF_no_NA$AUDIT.Score.Final))
```

The linear model may not be adequate. Distribution ressembles poisson distribution.

#### Regressions

##### 1. Linear Model

```
model <- lm(AUDIT.Score.Final~ Sex*scale(max_age_MHQ_alcohol, scale = FALSE) + Sex*I(scale(max_age_MHQ_alcohol, scale = FALSE)^2), data = Depression_DF_no_NA, na.action = na.exclude)
summary(model)
```

```
## 
## Call:
## lm(formula = AUDIT.Score.Final ~ Sex * scale(max_age_MHQ_alcohol, 
##     scale = FALSE) + Sex * I(scale(max_age_MHQ_alcohol, scale = FALSE)^2), 
##     data = Depression_DF_no_NA, na.action = na.exclude)
## 
## Residuals:
##    Min     1Q Median     3Q    Max 
## -6.724 -2.608 -0.725  1.523 33.676 
## 
## Coefficients:
##                                                      Estimate Std. Error
## (Intercept)                                         5.1366394  0.0142717
## Sex                                                -2.1396073  0.0285434
## scale(max_age_MHQ_alcohol, scale = FALSE)          -0.0846253  0.0013593
## I(scale(max_age_MHQ_alcohol, scale = FALSE)^2)     -0.0022644  0.0001671
## Sex:scale(max_age_MHQ_alcohol, scale = FALSE)      -0.0052733  0.0027185
## Sex:I(scale(max_age_MHQ_alcohol, scale = FALSE)^2)  0.0019608  0.0003342
##                                                    t value Pr(>|t|)    
## (Intercept)                                        359.918  < 2e-16 ***
## Sex                                                -74.960  < 2e-16 ***
## scale(max_age_MHQ_alcohol, scale = FALSE)          -62.259  < 2e-16 ***
## I(scale(max_age_MHQ_alcohol, scale = FALSE)^2)     -13.552  < 2e-16 ***
## Sex:scale(max_age_MHQ_alcohol, scale = FALSE)       -1.940   0.0524 .  
## Sex:I(scale(max_age_MHQ_alcohol, scale = FALSE)^2)   5.867 4.44e-09 ***
## ---
## Signif. codes:  0 '***' 0.001 '**' 0.01 '*' 0.05 '.' 0.1 ' ' 1
## 
## Residual standard error: 3.945 on 154805 degrees of freedom
##   (2431 observations deleted due to missingness)
## Multiple R-squared:  0.07792,    Adjusted R-squared:  0.07789 
## F-statistic:  2616 on 5 and 154805 DF,  p-value: < 2.2e-16
```

```
plot(model)
```

Residuals are not normally distributed. Let us try poisson regression.

##### Scaled

```
model <- lm(scale(AUDIT.Score.Final) ~ Sex*scale(max_age_MHQ_alcohol) + Sex*I(scale(max_age_MHQ_alcohol, scale = FALSE)^2), data = Depression_DF_no_NA, na.action = na.exclude)
summary(model)
```

```
## 
## Call:
## lm(formula = scale(AUDIT.Score.Final) ~ Sex * scale(max_age_MHQ_alcohol) + 
##     Sex * I(scale(max_age_MHQ_alcohol, scale = FALSE)^2), data = Depression_DF_no_NA, 
##     na.action = na.exclude)
## 
## Residuals:
##     Min      1Q  Median      3Q     Max 
## -1.5993 -0.6203 -0.1725  0.3622  8.0103 
## 
## Coefficients:
##                                                      Estimate Std. Error
## (Intercept)                                         4.547e-02  3.395e-03
## Sex                                                -5.089e-01  6.789e-03
## scale(max_age_MHQ_alcohol)                         -1.556e-01  2.499e-03
## I(scale(max_age_MHQ_alcohol, scale = FALSE)^2)     -5.386e-04  3.975e-05
## Sex:scale(max_age_MHQ_alcohol)                     -9.693e-03  4.997e-03
## Sex:I(scale(max_age_MHQ_alcohol, scale = FALSE)^2)  4.664e-04  7.949e-05
##                                                    t value Pr(>|t|)    
## (Intercept)                                         13.395  < 2e-16 ***
## Sex                                                -74.960  < 2e-16 ***
## scale(max_age_MHQ_alcohol)                         -62.259  < 2e-16 ***
## I(scale(max_age_MHQ_alcohol, scale = FALSE)^2)     -13.552  < 2e-16 ***
## Sex:scale(max_age_MHQ_alcohol)                      -1.940   0.0524 .  
## Sex:I(scale(max_age_MHQ_alcohol, scale = FALSE)^2)   5.867 4.44e-09 ***
## ---
## Signif. codes:  0 '***' 0.001 '**' 0.01 '*' 0.05 '.' 0.1 ' ' 1
## 
## Residual standard error: 0.9384 on 154805 degrees of freedom
##   (2431 observations deleted due to missingness)
## Multiple R-squared:  0.07792,    Adjusted R-squared:  0.07789 
## F-statistic:  2616 on 5 and 154805 DF,  p-value: < 2.2e-16
```

```
plot(model)
```

##### 2. Poisson Model

Poisson regression is often used for modeling count data.

Assumption : conditional variance is equal to the conditional mean -> test overdispersion

```
model1 <- glm(AUDIT.Score.Final~ Sex*scale(max_age_MHQ_alcohol, scale = FALSE) + Sex*I(scale(max_age_MHQ_alcohol, scale = FALSE)^2), data = Depression_DF_no_NA, family="poisson", na.action = na.exclude)
summary(model1)
```

```
## 
## Call:
## glm(formula = AUDIT.Score.Final ~ Sex * scale(max_age_MHQ_alcohol, 
##     scale = FALSE) + Sex * I(scale(max_age_MHQ_alcohol, scale = FALSE)^2), 
##     family = "poisson", data = Depression_DF_no_NA, na.action = na.exclude)
## 
## Deviance Residuals: 
##     Min       1Q   Median       3Q      Max  
## -3.6674  -1.3816  -0.3642   0.6911  10.1096  
## 
## Coefficients:
##                                                      Estimate Std. Error
## (Intercept)                                         1.614e+00  1.608e-03
## Sex                                                -4.239e-01  3.217e-03
## scale(max_age_MHQ_alcohol, scale = FALSE)          -1.840e-02  1.634e-04
## I(scale(max_age_MHQ_alcohol, scale = FALSE)^2)     -5.849e-04  1.940e-05
## Sex:scale(max_age_MHQ_alcohol, scale = FALSE)      -8.461e-03  3.268e-04
## Sex:I(scale(max_age_MHQ_alcohol, scale = FALSE)^2)  8.761e-05  3.881e-05
##                                                     z value Pr(>|z|)    
## (Intercept)                                        1003.621   <2e-16 ***
## Sex                                                -131.795   <2e-16 ***
## scale(max_age_MHQ_alcohol, scale = FALSE)          -112.578   <2e-16 ***
## I(scale(max_age_MHQ_alcohol, scale = FALSE)^2)      -30.143   <2e-16 ***
## Sex:scale(max_age_MHQ_alcohol, scale = FALSE)       -25.892   <2e-16 ***
## Sex:I(scale(max_age_MHQ_alcohol, scale = FALSE)^2)    2.257    0.024 *  
## ---
## Signif. codes:  0 '***' 0.001 '**' 0.01 '*' 0.05 '.' 0.1 ' ' 1
## 
## (Dispersion parameter for poisson family taken to be 1)
## 
##     Null deviance: 497837  on 154810  degrees of freedom
## Residual deviance: 455505  on 154805  degrees of freedom
##   (2431 observations deleted due to missingness)
## AIC: 924079
## 
## Number of Fisher Scoring iterations: 5
```

Our residual deviance is 625054 for 157014 degrees of freedom. The rule of thumb is ratio = 1, here : 625054/157014 = 3.98 - So we have moderate dispersion, which we can also test with a dispersion test

```
library("AER")
```

```
## Loading required package: car
```

```
## Loading required package: carData
```

```
## Registered S3 methods overwritten by 'car':
##   method                          from
##   influence.merMod                lme4
##   cooks.distance.influence.merMod lme4
##   dfbeta.influence.merMod         lme4
##   dfbetas.influence.merMod        lme4
```

```
## 
## Attaching package: 'car'
```

```
## The following object is masked from 'package:dplyr':
## 
##     recode
```

```
## Loading required package: lmtest
```

```
## Loading required package: zoo
```

```
## 
## Attaching package: 'zoo'
```

```
## The following objects are masked from 'package:base':
## 
##     as.Date, as.Date.numeric
```

```
## Loading required package: survival
```

```
dispersiontest(model1)
```

```
## 
##  Overdispersion test
## 
## data:  model1
## z = NA, p-value = NA
## alternative hypothesis: true dispersion is greater than 1
## sample estimates:
## dispersion 
##         NA
```

##### 3. “Fixing” overdispersion - negative binomial regression

Conditional variance exceedes the conditional mean -> test overdispersion

Maybe our distributional assumption was simply wrong, and we choose a different distribution

https://biometry.github.io/APES/LectureNotes/2016-JAGS/Overdispersion/OverdispersionJAGS.pdf https://stats.idre.ucla.edu/r/dae/negative-binomial-regression/

```
library(MASS)
```

```
## 
## Attaching package: 'MASS'
```

```
## The following object is masked from 'package:dplyr':
## 
##     select
```

```
model_2 <- glm.nb(AUDIT.Score.Final~ Sex*scale(max_age_MHQ_alcohol, scale = FALSE)+ Sex*I(scale(max_age_MHQ_alcohol, scale = FALSE)^2), data = Depression_DF_no_NA, na.action = na.exclude)
summary(model_2)
```

```
## 
## Call:
## glm.nb(formula = AUDIT.Score.Final ~ Sex * scale(max_age_MHQ_alcohol, 
##     scale = FALSE) + Sex * I(scale(max_age_MHQ_alcohol, scale = FALSE)^2), 
##     data = Depression_DF_no_NA, na.action = na.exclude, init.theta = 2.466437634, 
##     link = log)
## 
## Deviance Residuals: 
##     Min       1Q   Median       3Q      Max  
## -2.5474  -0.8802  -0.2164   0.4094   5.0207  
## 
## Coefficients:
##                                                      Estimate Std. Error
## (Intercept)                                         1.614e+00  2.812e-03
## Sex                                                -4.239e-01  5.625e-03
## scale(max_age_MHQ_alcohol, scale = FALSE)          -1.842e-02  2.738e-04
## I(scale(max_age_MHQ_alcohol, scale = FALSE)^2)     -5.891e-04  3.331e-05
## Sex:scale(max_age_MHQ_alcohol, scale = FALSE)      -8.463e-03  5.476e-04
## Sex:I(scale(max_age_MHQ_alcohol, scale = FALSE)^2)  8.741e-05  6.662e-05
##                                                    z value Pr(>|z|)    
## (Intercept)                                        574.003   <2e-16 ***
## Sex                                                -75.371   <2e-16 ***
## scale(max_age_MHQ_alcohol, scale = FALSE)          -67.275   <2e-16 ***
## I(scale(max_age_MHQ_alcohol, scale = FALSE)^2)     -17.686   <2e-16 ***
## Sex:scale(max_age_MHQ_alcohol, scale = FALSE)      -15.456   <2e-16 ***
## Sex:I(scale(max_age_MHQ_alcohol, scale = FALSE)^2)   1.312    0.189    
## ---
## Signif. codes:  0 '***' 0.001 '**' 0.01 '*' 0.05 '.' 0.1 ' ' 1
## 
## (Dispersion parameter for Negative Binomial(2.4664) family taken to be 1)
## 
##     Null deviance: 186579  on 154810  degrees of freedom
## Residual deviance: 172108  on 154805  degrees of freedom
##   (2431 observations deleted due to missingness)
## AIC: 794093
## 
## Number of Fisher Scoring iterations: 1
## 
## 
##               Theta:  2.4664 
##           Std. Err.:  0.0141 
## 
##  2 x log-likelihood:  -794079.0790
```

```
1 - pchisq(summary(model_2)$deviance,
           summary(model_2)$df.residual)
```

```
## [1] 0
```

The ratio of deviance 170198/157014 near 1 and thus fine. We can also check with a dispersion test.

### c. Let us use a likelihood ratio test to compare these two and test this model assumption

“As we mentioned earlier, negative binomial models assume the conditional means are not equal to the conditional variances. This inequality is captured by estimating a dispersion parameter (not shown in the output) that is held constant in a Poisson model. Thus, the Poisson model is actually nested in the negative binomial model. We can then use a likelihood ratio test to compare these two and test this model assumption.”

https://stats.idre.ucla.edu/r/dae/negative-binomial-regression/

```
pchisq(2 * (logLik(model_2) - logLik(model1)), df = 1, lower.tail = FALSE)
```

```
## 'log Lik.' 0 (df=7)
```

```
library("lmtest")
lrtest(model1, model_2)
```

```
## Likelihood ratio test
## 
## Model 1: AUDIT.Score.Final ~ Sex * scale(max_age_MHQ_alcohol, scale = FALSE) + 
##     Sex * I(scale(max_age_MHQ_alcohol, scale = FALSE)^2)
## Model 2: AUDIT.Score.Final ~ Sex * scale(max_age_MHQ_alcohol, scale = FALSE) + 
##     Sex * I(scale(max_age_MHQ_alcohol, scale = FALSE)^2)
##   #Df  LogLik Df  Chisq Pr(>Chisq)    
## 1   6 -462034                         
## 2   7 -397040  1 129988  < 2.2e-16 ***
## ---
## Signif. codes:  0 '***' 0.001 '**' 0.01 '*' 0.05 '.' 0.1 ' ' 1
```

In this example the associated chi-squared value estimated from 2\*(logLik(m1) – logLik(m3)) is 926.03 with one degree of freedom. This strongly suggests the negative binomial model, estimating the dispersion parameter, is more appropriate than the Poisson model.

### Save Negative Binomial Output

```
Model_summ_coef <- as.data.frame(summary(model_2)$coefficients)
Model_summ_coef$Model <- "AUDIT"
names(Model_summ_coef) <- c("Estimate", "SE", "t/z", "p", "Model")
fwrite( Model_summ_coef, "/scratch2/ukbio/biobank/High_IQ_Project/results/Sex_Age_Effects_AUDIT.csv")
```
