## Supplementary material for "High Intelligence is not a Risk Factor for Mental Health Disorders": Supplemental_File_S2_GAD_Age_Sex.html


### GAD\_Age\_Sex

### GAD7 anxiety score - Age and Sex Effects

#### GAD7 Score Distribution

##### Distribution of Raw GAD7 Score

```
hist(Anxiety_DF_no_NA$GAD7.Severity.Final)
```

##### Distribution of log transformed GAD7 Score

```
hist(log(Anxiety_DF_no_NA$GAD7.Severity.Final))
```

The linear model may not be adequate. Distribution resembles poisson distribution.

#### Regressions

##### 1. Linear Model

```
model <- lm(GAD7.Severity.Final~ Sex*scale(max_age_MHQ, scale = FALSE) + Sex*I(scale(max_age_MHQ, scale = FALSE)^2), data = Anxiety_DF_no_NA, na.action = na.exclude)
summary(model)
```

```
## 
## Call:
## lm(formula = GAD7.Severity.Final ~ Sex * scale(max_age_MHQ, scale = FALSE) + 
##     Sex * I(scale(max_age_MHQ, scale = FALSE)^2), data = Anxiety_DF_no_NA, 
##     na.action = na.exclude)
## 
## Residuals:
##     Min      1Q  Median      3Q     Max 
## -2.4735 -1.2951 -0.8697  0.3164 20.2917 
## 
## Coefficients:
##                                              Estimate Std. Error t value
## (Intercept)                                 1.2498394  0.0114184 109.459
## Sex                                         0.4563931  0.0228367  19.985
## scale(max_age_MHQ, scale = FALSE)          -0.0412460  0.0011077 -37.235
## I(scale(max_age_MHQ, scale = FALSE)^2)      0.0007295  0.0001344   5.429
## Sex:scale(max_age_MHQ, scale = FALSE)      -0.0083602  0.0022154  -3.774
## Sex:I(scale(max_age_MHQ, scale = FALSE)^2) -0.0004881  0.0002687  -1.816
##                                            Pr(>|t|)    
## (Intercept)                                 < 2e-16 ***
## Sex                                         < 2e-16 ***
## scale(max_age_MHQ, scale = FALSE)           < 2e-16 ***
## I(scale(max_age_MHQ, scale = FALSE)^2)     5.67e-08 ***
## Sex:scale(max_age_MHQ, scale = FALSE)      0.000161 ***
## Sex:I(scale(max_age_MHQ, scale = FALSE)^2) 0.069320 .  
## ---
## Signif. codes:  0 '***' 0.001 '**' 0.01 '*' 0.05 '.' 0.1 ' ' 1
## 
## Residual standard error: 2.664 on 107825 degrees of freedom
##   (49317 observations deleted due to missingness)
## Multiple R-squared:  0.02329,    Adjusted R-squared:  0.02324 
## F-statistic: 514.1 on 5 and 107825 DF,  p-value: < 2.2e-16
```

```
plot(model)
```

Residuals are not normally distributed. Let us try poisson regression.

##### 2. Poisson Model

Poisson regression is often used for modeling count data.

Assumption : conditional variance is equal to the conditional mean -> test overdispersion

```
model1 <- glm(GAD7.Severity.Final~ Sex*scale(max_age_MHQ, scale = FALSE) + Sex*I(scale(max_age_MHQ, scale = FALSE)^2), data = Anxiety_DF_no_NA, family="poisson", na.action = na.exclude)
summary(model1)
```

```
## 
## Call:
## glm(formula = GAD7.Severity.Final ~ Sex * scale(max_age_MHQ, 
##     scale = FALSE) + Sex * I(scale(max_age_MHQ, scale = FALSE)^2), 
##     family = "poisson", data = Anxiety_DF_no_NA, na.action = na.exclude)
## 
## Deviance Residuals: 
##     Min       1Q   Median       3Q      Max  
## -2.2244  -1.6024  -1.3038   0.2965  10.1345  
## 
## Coefficients:
##                                              Estimate Std. Error z value
## (Intercept)                                 2.068e-01  3.858e-03  53.604
## Sex                                         3.677e-01  7.717e-03  47.651
## scale(max_age_MHQ, scale = FALSE)          -3.244e-02  3.972e-04 -81.661
## I(scale(max_age_MHQ, scale = FALSE)^2)      6.211e-05  4.487e-05   1.384
## Sex:scale(max_age_MHQ, scale = FALSE)       3.513e-03  7.945e-04   4.422
## Sex:I(scale(max_age_MHQ, scale = FALSE)^2) -4.049e-04  8.974e-05  -4.512
##                                            Pr(>|z|)    
## (Intercept)                                 < 2e-16 ***
## Sex                                         < 2e-16 ***
## scale(max_age_MHQ, scale = FALSE)           < 2e-16 ***
## I(scale(max_age_MHQ, scale = FALSE)^2)        0.166    
## Sex:scale(max_age_MHQ, scale = FALSE)      9.77e-06 ***
## Sex:I(scale(max_age_MHQ, scale = FALSE)^2) 6.42e-06 ***
## ---
## Signif. codes:  0 '***' 0.001 '**' 0.01 '*' 0.05 '.' 0.1 ' ' 1
## 
## (Dispersion parameter for poisson family taken to be 1)
## 
##     Null deviance: 364525  on 107830  degrees of freedom
## Residual deviance: 350860  on 107825  degrees of freedom
##   (49317 observations deleted due to missingness)
## AIC: 469986
## 
## Number of Fisher Scoring iterations: 6
```

Our residual deviance is 625054 for 157014 degrees of freedom. The rule of thumb is ratio = 1, here : 625054/157014 = 3.98 - So we have moderate dispersion, which we can also test with a dispersion test

```
library("AER")
```

```
## Loading required package: car
```

```
## Loading required package: carData
```

```
## Registered S3 methods overwritten by 'car':
##   method                          from
##   influence.merMod                lme4
##   cooks.distance.influence.merMod lme4
##   dfbeta.influence.merMod         lme4
##   dfbetas.influence.merMod        lme4
```

```
## 
## Attaching package: 'car'
```

```
## The following object is masked from 'package:dplyr':
## 
##     recode
```

```
## Loading required package: lmtest
```

```
## Loading required package: zoo
```

```
## 
## Attaching package: 'zoo'
```

```
## The following objects are masked from 'package:base':
## 
##     as.Date, as.Date.numeric
```

```
## Loading required package: survival
```

```
dispersiontest(model1)
```

```
## 
##  Overdispersion test
## 
## data:  model1
## z = NA, p-value = NA
## alternative hypothesis: true dispersion is greater than 1
## sample estimates:
## dispersion 
##         NA
```

##### 3. “Fixing” overdispersionby using negative binomial regression by using negative binomial regression

Conditional variance exceedes the conditional mean -> test overdispersion Maybe our distributional assumption was simply wrong, and we choose a different distribution

https://biometry.github.io/APES/LectureNotes/2016-JAGS/Overdispersion/OverdispersionJAGS.pdf https://stats.idre.ucla.edu/r/dae/negative-binomial-regression/

```
library(MASS)
```

```
## 
## Attaching package: 'MASS'
```

```
## The following object is masked from 'package:dplyr':
## 
##     select
```

```
model_2 <- glm.nb(GAD7.Severity.Final~ Sex*scale(max_age_MHQ, scale = FALSE)+ Sex*I(scale(max_age_MHQ, scale = FALSE)^2), data = Anxiety_DF_no_NA, na.action = na.exclude)
summary(model_2)
```

```
## 
## Call:
## glm.nb(formula = GAD7.Severity.Final ~ Sex * scale(max_age_MHQ, 
##     scale = FALSE) + Sex * I(scale(max_age_MHQ, scale = FALSE)^2), 
##     data = Anxiety_DF_no_NA, na.action = na.exclude, init.theta = 0.3255206086, 
##     link = log)
## 
## Deviance Residuals: 
##     Min       1Q   Median       3Q      Max  
## -1.1894  -1.0184  -0.9136   0.1388   3.6445  
## 
## Coefficients:
##                                              Estimate Std. Error z value
## (Intercept)                                 1.984e-01  8.473e-03  23.410
## Sex                                         3.718e-01  1.695e-02  21.941
## scale(max_age_MHQ, scale = FALSE)          -3.202e-02  8.298e-04 -38.582
## I(scale(max_age_MHQ, scale = FALSE)^2)      2.041e-04  9.961e-05   2.049
## Sex:scale(max_age_MHQ, scale = FALSE)       3.560e-03  1.660e-03   2.145
## Sex:I(scale(max_age_MHQ, scale = FALSE)^2) -4.596e-04  1.992e-04  -2.307
##                                            Pr(>|z|)    
## (Intercept)                                  <2e-16 ***
## Sex                                          <2e-16 ***
## scale(max_age_MHQ, scale = FALSE)            <2e-16 ***
## I(scale(max_age_MHQ, scale = FALSE)^2)       0.0404 *  
## Sex:scale(max_age_MHQ, scale = FALSE)        0.0319 *  
## Sex:I(scale(max_age_MHQ, scale = FALSE)^2)   0.0210 *  
## ---
## Signif. codes:  0 '***' 0.001 '**' 0.01 '*' 0.05 '.' 0.1 ' ' 1
## 
## (Dispersion parameter for Negative Binomial(0.3255) family taken to be 1)
## 
##     Null deviance: 90684  on 107830  degrees of freedom
## Residual deviance: 87945  on 107825  degrees of freedom
##   (49317 observations deleted due to missingness)
## AIC: 314392
## 
## Number of Fisher Scoring iterations: 1
## 
## 
##               Theta:  0.32552 
##           Std. Err.:  0.00247 
## 
##  2 x log-likelihood:  -314378.41700
```

```
1 - pchisq(summary(model_2)$deviance,
           summary(model_2)$df.residual)
```

```
## [1] 1
```

The ratio of deviance under 1 (87945/107825). We can also check with a dispersion test. But we get an error suggesting this is not the appropriate regression model.

error: Error in securityAssertion(“Simulation from the model produced wrong dimension”, : Message from DHARMa: During the execution of a DHARMa function, some unexpected conditions occurred. Even if you didn’t get an error, your results may not be reliable. Please check with the help if you use the functions as intended. If you think that the error is not on your side, I would be grateful if you could report the problem at https://github.com/florianhartig/DHARMa/issues

Context: Simulation from the model produced wrong dimension

### c. Let us use a likelihood ratio test to compare these two and test this model assumption

“As we mentioned earlier, negative binomial models assume the conditional means are not equal to the conditional variances. This inequality is captured by estimating a dispersion parameter (not shown in the output) that is held constant in a Poisson model. Thus, the Poisson model is actually nested in the negative binomial model. We can then use a likelihood ratio test to compare these two and test this model assumption.”

https://stats.idre.ucla.edu/r/dae/negative-binomial-regression/

```
pchisq(2 * (logLik(model_2) - logLik(model1)), df = 1, lower.tail = FALSE)
```

```
## 'log Lik.' 0 (df=7)
```

```
library("lmtest")
lrtest(model1, model_2)
```

```
## Likelihood ratio test
## 
## Model 1: GAD7.Severity.Final ~ Sex * scale(max_age_MHQ, scale = FALSE) + 
##     Sex * I(scale(max_age_MHQ, scale = FALSE)^2)
## Model 2: GAD7.Severity.Final ~ Sex * scale(max_age_MHQ, scale = FALSE) + 
##     Sex * I(scale(max_age_MHQ, scale = FALSE)^2)
##   #Df  LogLik Df  Chisq Pr(>Chisq)    
## 1   6 -234987                         
## 2   7 -157189  1 155596  < 2.2e-16 ***
## ---
## Signif. codes:  0 '***' 0.001 '**' 0.01 '*' 0.05 '.' 0.1 ' ' 1
```

In this example the associated chi-squared value estimated from 2\*(logLik(m1) – logLik(m3)) is 926.03 with one degree of freedom. This strongly suggests the negative binomial model, estimating the dispersion parameter, is more appropriate than the Poisson model.
