## Supplementary material for "High Intelligence is not a Risk Factor for Mental Health Disorders": Supplemental_File_S3_Diff_Sex_N_Sex_Age.html

Diff\_Sex\_N\_Age\_Sex


### Diff\_Sex\_N\_Age\_Sex

### I. Diff\_Sex\_Partners\_N

#### a) No outlier removal

##### Distribution of Raw Neuroticism Score

```
hist(Sexual_Beh_DF$Diff_Sex_Partners_N)
```

```
length(Sexual_Beh_DF$Diff_Sex_Partners_N)
```

```
## [1] 502120
```

```
summary(Sexual_Beh_DF$Diff_Sex_Partners_N)
```

```
##      Min.   1st Qu.    Median      Mean   3rd Qu.      Max. 
##    -1.000     1.000     2.000     6.042     5.000 15000.000
```

#### Regressions

##### 1. Poisson Regression Model

Poisson regression is often used for modeling count data.

Assumption : conditional variance is equal to the conditional mean -> test overdispersion

```
model1 <- glm(Diff_Sex_Partners_N ~ Sex * scale(max_age_same_N, scale = FALSE) + Sex * I(scale((max_age_same_N), scale = FALSE)^2), 
                             data = Sexual_Beh_DF_no_NA, family = "poisson")
summary(model1)
```

```
## 
## Call:
## glm(formula = Diff_Sex_Partners_N ~ Sex * scale(max_age_same_N, 
##     scale = FALSE) + Sex * I(scale((max_age_same_N), scale = FALSE)^2), 
##     family = "poisson", data = Sexual_Beh_DF_no_NA)
## 
## Deviance Residuals: 
##    Min      1Q  Median      3Q     Max  
## -11.67   -2.43   -1.15    0.44  365.49  
## 
## Coefficients:
##                                                   Estimate Std. Error z value
## (Intercept)                                      1.873e+00  7.547e-04 2481.32
## Sex                                             -7.048e-01  1.509e-03 -466.95
## scale(max_age_same_N, scale = FALSE)             7.427e-02  2.196e-04  338.24
## I(scale((max_age_same_N), scale = FALSE)^2)     -8.797e-04  4.183e-06 -210.31
## Sex:scale(max_age_same_N, scale = FALSE)        -3.901e-02  4.392e-04  -88.83
## Sex:I(scale((max_age_same_N), scale = FALSE)^2)  4.443e-04  8.366e-06   53.10
##                                                 Pr(>|z|)    
## (Intercept)                                       <2e-16 ***
## Sex                                               <2e-16 ***
## scale(max_age_same_N, scale = FALSE)              <2e-16 ***
## I(scale((max_age_same_N), scale = FALSE)^2)       <2e-16 ***
## Sex:scale(max_age_same_N, scale = FALSE)          <2e-16 ***
## Sex:I(scale((max_age_same_N), scale = FALSE)^2)   <2e-16 ***
## ---
## Signif. codes:  0 '***' 0.001 '**' 0.01 '*' 0.05 '.' 0.1 ' ' 1
## 
## (Dispersion parameter for poisson family taken to be 1)
## 
##     Null deviance: 8806518  on 452054  degrees of freedom
## Residual deviance: 7208242  on 452049  degrees of freedom
## AIC: 8478757
## 
## Number of Fisher Scoring iterations: 7
```

Our residual deviance is 8275478 for 452049 degrees of freedom. The rule of thumb is ratio = 1, here : 8275478/452049 = 18.30 - So we have moderate dispersion, which we can also test with a dispersion test

```
library("AER")
```

```
## Loading required package: car
```

```
## Loading required package: carData
```

```
## Registered S3 methods overwritten by 'car':
##   method                          from
##   influence.merMod                lme4
##   cooks.distance.influence.merMod lme4
##   dfbeta.influence.merMod         lme4
##   dfbetas.influence.merMod        lme4
```

```
## 
## Attaching package: 'car'
```

```
## The following object is masked from 'package:dplyr':
## 
##     recode
```

```
## Loading required package: lmtest
```

```
## Loading required package: zoo
```

```
## 
## Attaching package: 'zoo'
```

```
## The following objects are masked from 'package:base':
## 
##     as.Date, as.Date.numeric
```

```
## Loading required package: survival
```

```
dispersiontest(model1)
```

```
## 
##  Overdispersion test
## 
## data:  model1
## z = 4.2158, p-value = 1.244e-05
## alternative hypothesis: true dispersion is greater than 1
## sample estimates:
## dispersion 
##   139.0959
```

```
1 - pchisq(summary(model1)$deviance,
           summary(model1)$df.residual)
```

```
## [1] 0
```

The GOF test indicates that the model fits the data if p > 0.05.

##### 3. “Fixing” overdispersion

### a. by using quasi-families regression

quasi-families augment the normal families by adding a dispersion parameter http://biometry.github.io/APES//LectureNotes/2016-JAGS/Overdispersion/OverdispersionJAGS.html

```
model <- glm(Diff_Sex_Partners_N~ Sex*max_age_same_N + Sex*I(max_age_same_N^2), data = Sexual_Beh_DF_no_NA, family="quasipoisson", na.action = na.exclude)
summary(model)
```

```
## 
## Call:
## glm(formula = Diff_Sex_Partners_N ~ Sex * max_age_same_N + Sex * 
##     I(max_age_same_N^2), family = "quasipoisson", data = Sexual_Beh_DF_no_NA, 
##     na.action = na.exclude)
## 
## Deviance Residuals: 
##    Min      1Q  Median      3Q     Max  
## -11.67   -2.43   -1.15    0.44  365.49  
## 
## Coefficients:
##                           Estimate Std. Error t value Pr(>|t|)    
## (Intercept)              1.798e+00  7.876e-03 228.244  < 2e-16 ***
## Sex                     -6.654e-01  1.575e-02 -42.243  < 2e-16 ***
## max_age_same_N           7.603e-02  2.687e-03  28.293  < 2e-16 ***
## I(max_age_same_N^2)     -8.797e-04  4.934e-05 -17.831  < 2e-16 ***
## Sex:max_age_same_N      -3.990e-02  5.375e-03  -7.424 1.14e-13 ***
## Sex:I(max_age_same_N^2)  4.442e-04  9.867e-05   4.502 6.72e-06 ***
## ---
## Signif. codes:  0 '***' 0.001 '**' 0.01 '*' 0.05 '.' 0.1 ' ' 1
## 
## (Dispersion parameter for quasipoisson family taken to be 139.1075)
## 
##     Null deviance: 8806518  on 452054  degrees of freedom
## Residual deviance: 7208242  on 452049  degrees of freedom
## AIC: NA
## 
## Number of Fisher Scoring iterations: 7
```

```
1 - pchisq(summary(model)$deviance,
           summary(model)$df.residual)
```

```
## [1] 0
```

The null hypothesis is that our model is correctly specified, and we have strong evidence to reject that hypothesis. So we have strong evidence that our model fits badly. Here we simulated the data, and we in fact know that the model we have fitted is the correct model. So here the deviance goodness of fit test has wrongly indicated that our model is incorrectly specified. But perhaps we were just unlucky – by chance 5% of the time the test will reject even when the null hypothesis is true.

You see that τ (dispersion parameter) is estimated at a value similar to those in the overdispersion tests above (as you’d expect). The main effect is the substantially larger errors for the estimates (the point estimates do not change), and hence potentially changed significances (though not here). (You can manually compute the corrected standard errors as Poisson-standard errors ·√τ .) Note that because this is no maximum likelihoo method (but a quasi-likelihood method), no likelihood and hence no AIC are available. No overdispersion tests can be conducted for quasi-family objects (neither in AER nor DHARMa)

### b. by using negative binomial regression

Conditional variance exceedes the conditional mean -> test overdispersion Maybe our distributional assumption was simply wrong, and we choose a different distribution

https://biometry.github.io/APES/LectureNotes/2016-JAGS/Overdispersion/OverdispersionJAGS.pdf https://stats.idre.ucla.edu/r/dae/negative-binomial-regression/

```
library(MASS)
```

```
## 
## Attaching package: 'MASS'
```

```
## The following object is masked from 'package:dplyr':
## 
##     select
```

```
model_2 <- glm.nb(Diff_Sex_Partners_N~ Sex*scale(max_age_same_N, scale = FALSE)+ Sex*I(scale(max_age_same_N, scale = FALSE)^2), data = Sexual_Beh_DF_no_NA, na.action = na.exclude)
summary(model_2)
```

```
## 
## Call:
## glm.nb(formula = Diff_Sex_Partners_N ~ Sex * scale(max_age_same_N, 
##     scale = FALSE) + Sex * I(scale(max_age_same_N, scale = FALSE)^2), 
##     data = Sexual_Beh_DF_no_NA, na.action = na.exclude, init.theta = 0.7487790746, 
##     link = log)
## 
## Deviance Residuals: 
##    Min      1Q  Median      3Q     Max  
## -2.598  -0.879  -0.497   0.139  43.095  
## 
## Coefficients:
##                                                 Estimate Std. Error z value
## (Intercept)                                    1.871e+00  2.845e-03  657.73
## Sex                                           -7.048e-01  5.690e-03 -123.88
## scale(max_age_same_N, scale = FALSE)           7.335e-02  1.087e-03   67.46
## I(scale(max_age_same_N, scale = FALSE)^2)     -8.626e-04  2.004e-05  -43.05
## Sex:scale(max_age_same_N, scale = FALSE)      -3.873e-02  2.175e-03  -17.81
## Sex:I(scale(max_age_same_N, scale = FALSE)^2)  4.410e-04  4.008e-05   11.01
##                                               Pr(>|z|)    
## (Intercept)                                     <2e-16 ***
## Sex                                             <2e-16 ***
## scale(max_age_same_N, scale = FALSE)            <2e-16 ***
## I(scale(max_age_same_N, scale = FALSE)^2)       <2e-16 ***
## Sex:scale(max_age_same_N, scale = FALSE)        <2e-16 ***
## Sex:I(scale(max_age_same_N, scale = FALSE)^2)   <2e-16 ***
## ---
## Signif. codes:  0 '***' 0.001 '**' 0.01 '*' 0.05 '.' 0.1 ' ' 1
## 
## (Dispersion parameter for Negative Binomial(0.7488) family taken to be 1)
## 
##     Null deviance: 595347  on 452054  degrees of freedom
## Residual deviance: 492054  on 452049  degrees of freedom
## AIC: 2552220
## 
## Number of Fisher Scoring iterations: 1
## 
## 
##               Theta:  0.74878 
##           Std. Err.:  0.00169 
## 
##  2 x log-likelihood:  -2552205.54200
```

```
1 - pchisq(summary(model_2)$deviance,
           summary(model_2)$df.residual)
```

```
## [1] 0
```

https://www.r-bloggers.com/2012/08/count-data-and-glms-choosing-among-poisson-negative-binomial-and-zero-inflated-models/ https://stats.stackexchange.com/questions/37732/when-someone-says-residual-deviance-df-should-1-for-a-poisson-model-how-appro

The ratio of deviance 495803/452049 = 1.09 and hence probably fine. We can also check with a dispersion test. The null hypothesis is that our model is correctly specified, and we have strong evidence to reject that hypothesis. So we have strong evidence that our model fits badly. Here we simulated the data, and we in fact know that the model we have fitted is the correct model. So here the deviance goodness of fit test has wrongly indicated that our model is incorrectly specified. But perhaps we were just unlucky – by chance 5% of the time the test will reject even when the null hypothesis is true.

```
library(DHARMa)
```

```
## This is DHARMa 0.4.1. For overview type '?DHARMa'. For recent changes, type news(package = 'DHARMa') Note: Syntax of plotResiduals has changed in 0.3.0, see ?plotResiduals for details
```

```
simulationOutput <- simulateResiduals(model_2)
testDispersion(simulationOutput)
```

```
## 
##  DHARMa nonparametric dispersion test via sd of residuals fitted vs.
##  simulated
## 
## data:  simulationOutput
## dispersion = 23.801, p-value < 2.2e-16
## alternative hypothesis: two.sided
```

```
testZeroInflation(simulationOutput)
```

```
## 
##  DHARMa zero-inflation test via comparison to expected zeros with
##  simulation under H0 = fitted model
## 
## data:  simulationOutput
## ratioObsSim = 0.50519, p-value < 2.2e-16
## alternative hypothesis: two.sided
```

https://stats.stackexchange.com/questions/490680/significant-dispersion-test

A word of warning that applies also to all other tests that follow: significance in hypothesis tests depends on at least 2 ingredients: strength of the signal, and number of data points. Hence, the p-value alone is not a good indicator of the extent to which your residuals deviate from assumptions. Specifically, if you have a lot of data points, residual diagnostics will nearly inevitably become significant, because having a perfectly fitting model is very unlikely. That, however, doesn’t necessarily mean that you need to change your model. The p-values confirm that there is a deviation from your null hypothesis. It is, however, in your discretion to decide whether this deviation is worth worrying about. If you see a dispersion parameter of 1.01, I would not worry, even if the test is significant. A significant value of 5, however, is clearly a reason to move to a model that accounts for overdispersion.

### c. Let us use a likelihood ratio test to compare these two and test this model assumption

“As we mentioned earlier, negative binomial models assume the conditional means are not equal to the conditional variances. This inequality is captured by estimating a dispersion parameter (not shown in the output) that is held constant in a Poisson model. Thus, the Poisson model is actually nested in the negative binomial model. We can then use a likelihood ratio test to compare these two and test this model assumption.”

https://stats.idre.ucla.edu/r/dae/negative-binomial-regression/

```
pchisq(2 * (logLik(model_2) - logLik(model1)), df = 1, lower.tail = FALSE)
```

```
## 'log Lik.' 0 (df=7)
```

```
library("lmtest")
lrtest(model1, model_2)
```

```
## Likelihood ratio test
## 
## Model 1: Diff_Sex_Partners_N ~ Sex * scale(max_age_same_N, scale = FALSE) + 
##     Sex * I(scale((max_age_same_N), scale = FALSE)^2)
## Model 2: Diff_Sex_Partners_N ~ Sex * scale(max_age_same_N, scale = FALSE) + 
##     Sex * I(scale(max_age_same_N, scale = FALSE)^2)
##   #Df   LogLik Df   Chisq Pr(>Chisq)    
## 1   6 -4239373                          
## 2   7 -1276103  1 5926539  < 2.2e-16 ***
## ---
## Signif. codes:  0 '***' 0.001 '**' 0.01 '*' 0.05 '.' 0.1 ' ' 1
```

In this example the associated chi-squared value estimated from 2\*(logLik(m1) – logLik(m3)) is 926.03 with one degree of freedom. This strongly suggests the negative binomial model, estimating the dispersion parameter, is more appropriate than the Poisson model.

#### B) Outlier removal 6 \* IQR

##### Distribution of Diff\_Sex\_Partners\_N

```
hist(Sexual_Beh_DT_na_outliers_DT$Diff_Sex_Partners_N)
```

```
length(Sexual_Beh_DT_na_outliers_DT$Diff_Sex_Partners_N)
```

```
## [1] 452055
```

```
summary(Sexual_Beh_DT_na_outliers_DT$Diff_Sex_Partners_N)
```

```
##    Min. 1st Qu.  Median    Mean 3rd Qu.    Max.    NA's 
##   0.000   1.000   3.000   4.603   6.000  36.000    8724
```

#### Regressions

##### 1. Poisson Regression Model

Poisson regression is often used for modeling count data.

Assumption : conditional variance is equal to the conditional mean -> test overdispersion

```
model1 <- glm(Diff_Sex_Partners_N ~ Sex * scale(max_age_same_N, scale = FALSE) + Sex * I(scale((max_age_same_N), scale = FALSE)^2), 
              data = Sexual_Beh_DT_na_outliers_DT, family = "poisson")
summary(model1)
```

```
## 
## Call:
## glm(formula = Diff_Sex_Partners_N ~ Sex * scale(max_age_same_N, 
##     scale = FALSE) + Sex * I(scale((max_age_same_N), scale = FALSE)^2), 
##     family = "poisson", data = Sexual_Beh_DT_na_outliers_DT)
## 
## Deviance Residuals: 
##     Min       1Q   Median       3Q      Max  
## -4.3908  -1.7062  -1.0102   0.5938   9.8849  
## 
## Coefficients:
##                                                   Estimate Std. Error z value
## (Intercept)                                      1.574e+00  9.905e-04 1588.86
## Sex                                             -3.066e-01  1.981e-03 -154.75
## scale(max_age_same_N, scale = FALSE)             3.390e-02  3.523e-04   96.21
## I(scale((max_age_same_N), scale = FALSE)^2)     -4.509e-04  6.573e-06  -68.60
## Sex:scale(max_age_same_N, scale = FALSE)         1.888e-02  7.046e-04   26.79
## Sex:I(scale((max_age_same_N), scale = FALSE)^2) -2.089e-04  1.315e-05  -15.89
##                                                 Pr(>|z|)    
## (Intercept)                                       <2e-16 ***
## Sex                                               <2e-16 ***
## scale(max_age_same_N, scale = FALSE)              <2e-16 ***
## I(scale((max_age_same_N), scale = FALSE)^2)       <2e-16 ***
## Sex:scale(max_age_same_N, scale = FALSE)          <2e-16 ***
## Sex:I(scale((max_age_same_N), scale = FALSE)^2)   <2e-16 ***
## ---
## Signif. codes:  0 '***' 0.001 '**' 0.01 '*' 0.05 '.' 0.1 ' ' 1
## 
## (Dispersion parameter for poisson family taken to be 1)
## 
##     Null deviance: 2363202  on 443330  degrees of freedom
## Residual deviance: 2274158  on 443325  degrees of freedom
##   (8724 observations deleted due to missingness)
## AIC: 3491641
## 
## Number of Fisher Scoring iterations: 5
```

Our residual deviance is 2284154 for 443393 degrees of freedom. The rule of thumb is ratio = 1, here : 2284154/443393 = 5.15 - So we have moderate dispersion, which we can also test with a dispersion test

```
library("AER")
dispersiontest(model1)
```

```
## 
##  Overdispersion test
## 
## data:  model1
## z = 198.55, p-value < 2.2e-16
## alternative hypothesis: true dispersion is greater than 1
## sample estimates:
## dispersion 
##    6.66609
```

##### 3. “Fixing” overdispersion

### a. by using quasi-families regression

quasi-families augment the normal families by adding a dispersion parameter http://biometry.github.io/APES//LectureNotes/2016-JAGS/Overdispersion/OverdispersionJAGS.html

```
model <- glm(Diff_Sex_Partners_N~ Sex*max_age_same_N + Sex*I(max_age_same_N^2), data = Sexual_Beh_DT_na_outliers_DT, family="quasipoisson", na.action = na.exclude)
summary(model)
```

```
## 
## Call:
## glm(formula = Diff_Sex_Partners_N ~ Sex * max_age_same_N + Sex * 
##     I(max_age_same_N^2), family = "quasipoisson", data = Sexual_Beh_DT_na_outliers_DT, 
##     na.action = na.exclude)
## 
## Deviance Residuals: 
##     Min       1Q   Median       3Q      Max  
## -4.3908  -1.7062  -1.0102   0.5938   9.8849  
## 
## Coefficients:
##                           Estimate Std. Error t value Pr(>|t|)    
## (Intercept)              1.539e+00  2.037e-03 755.641  < 2e-16 ***
## Sex                     -3.257e-01  4.075e-03 -79.923  < 2e-16 ***
## max_age_same_N           3.480e-02  9.432e-04  36.896  < 2e-16 ***
## I(max_age_same_N^2)     -4.509e-04  1.697e-05 -26.571  < 2e-16 ***
## Sex:max_age_same_N       1.930e-02  1.886e-03  10.229  < 2e-16 ***
## Sex:I(max_age_same_N^2) -2.089e-04  3.394e-05  -6.155 7.51e-10 ***
## ---
## Signif. codes:  0 '***' 0.001 '**' 0.01 '*' 0.05 '.' 0.1 ' ' 1
## 
## (Dispersion parameter for quasipoisson family taken to be 6.666355)
## 
##     Null deviance: 2363202  on 443330  degrees of freedom
## Residual deviance: 2274158  on 443325  degrees of freedom
##   (8724 observations deleted due to missingness)
## AIC: NA
## 
## Number of Fisher Scoring iterations: 5
```

You see that τ (dispersion parameter) is estimated at a value similar to those in the overdispersion tests above (as you’d expect). The main effect is the substantially larger errors for the estimates (the point estimates do not change), and hence potentially changed significances (though not here). (You can manually compute the corrected standard errors as Poisson-standard errors ·√τ .) Note that because this is no maximum likelihoo method (but a quasi-likelihood method), no likelihood and hence no AIC are available. No overdispersion tests can be conducted for quasi-family objects (neither in AER nor DHARMa)

### b. by using negative binomial regression

Conditional variance exceedes the conditional mean -> test overdispersion Maybe our distributional assumption was simply wrong, and we choose a different distribution

https://biometry.github.io/APES/LectureNotes/2016-JAGS/Overdispersion/OverdispersionJAGS.pdf https://stats.idre.ucla.edu/r/dae/negative-binomial-regression/

```
library(MASS)
model_2 <- glm.nb(Diff_Sex_Partners_N~ Sex*scale(max_age_same_N, scale = FALSE)+ Sex*I(scale(max_age_same_N, scale = FALSE)^2), data = Sexual_Beh_DT_na_outliers_DT, na.action = na.exclude)
summary(model_2)
```

```
## 
## Call:
## glm.nb(formula = Diff_Sex_Partners_N ~ Sex * scale(max_age_same_N, 
##     scale = FALSE) + Sex * I(scale(max_age_same_N, scale = FALSE)^2), 
##     data = Sexual_Beh_DT_na_outliers_DT, na.action = na.exclude, 
##     init.theta = 1.079014866, link = log)
## 
## Deviance Residuals: 
##     Min       1Q   Median       3Q      Max  
## -2.2219  -0.9342  -0.4904   0.2695   3.3973  
## 
## Coefficients:
##                                                 Estimate Std. Error z value
## (Intercept)                                    1.573e+00  2.583e-03 608.895
## Sex                                           -3.079e-01  5.166e-03 -59.611
## scale(max_age_same_N, scale = FALSE)           3.338e-02  1.006e-03  33.169
## I(scale(max_age_same_N, scale = FALSE)^2)     -4.408e-04  1.847e-05 -23.865
## Sex:scale(max_age_same_N, scale = FALSE)       1.823e-02  2.013e-03   9.056
## Sex:I(scale(max_age_same_N, scale = FALSE)^2) -1.958e-04  3.694e-05  -5.301
##                                               Pr(>|z|)    
## (Intercept)                                    < 2e-16 ***
## Sex                                            < 2e-16 ***
## scale(max_age_same_N, scale = FALSE)           < 2e-16 ***
## I(scale(max_age_same_N, scale = FALSE)^2)      < 2e-16 ***
## Sex:scale(max_age_same_N, scale = FALSE)       < 2e-16 ***
## Sex:I(scale(max_age_same_N, scale = FALSE)^2) 1.15e-07 ***
## ---
## Signif. codes:  0 '***' 0.001 '**' 0.01 '*' 0.05 '.' 0.1 ' ' 1
## 
## (Dispersion parameter for Negative Binomial(1.079) family taken to be 1)
## 
##     Null deviance: 494267  on 443330  degrees of freedom
## Residual deviance: 478072  on 443325  degrees of freedom
##   (8724 observations deleted due to missingness)
## AIC: 2314294
## 
## Number of Fisher Scoring iterations: 1
## 
## 
##               Theta:  1.07901 
##           Std. Err.:  0.00283 
## 
##  2 x log-likelihood:  -2314280.16800
```

```
1 - pchisq(summary(model_2)$deviance,
         summary(model_2)$df.residual)
```

```
## [1] 0
```

https://www.r-bloggers.com/2012/08/count-data-and-glms-choosing-among-poisson-negative-binomial-and-zero-inflated-models/ https://stats.stackexchange.com/questions/37732/when-someone-says-residual-deviance-df-should-1-for-a-poisson-model-how-appro

The ratio of deviance 477594/443393 = 1.07 We can also check with a dispersion test.

### `{r} # library(DHARMa) # simulationOutput <- simulateResiduals(model_2) # testDispersion(simulationOutput) # testZeroInflation(simulationOutput) #`

Error in securityAssertion(“Simulation from the model produced wrong dimension”, : Message from DHARMa: During the execution of a DHARMa function, some unexpected conditions occurred. Even if you didnt get an error, your results may not be reliable. Please check with the help if you use the functions as intended. If you think that the error is not on your side, I would be grateful if you could report the problem at https://github.com/florianhartig/DHARMa/issues Context: Simulation from the model produced wrong dimension

https://stats.stackexchange.com/questions/490680/significant-dispersion-test

A word of warning that applies also to all other tests that follow: significance in hypothesis tests depends on at least 2 ingredients: strength of the signal, and number of data points. Hence, the p-value alone is not a good indicator of the extent to which your residuals deviate from assumptions. Specifically, if you have a lot of data points, residual diagnostics will nearly inevitably become significant, because having a perfectly fitting model is very unlikely. That, however, doesn’t necessarily mean that you need to change your model. The p-values confirm that there is a deviation from your null hypothesis. It is, however, in your discretion to decide whether this deviation is worth worrying about. If you see a dispersion parameter of 1.01, I would not worry, even if the test is significant. A significant value of 5, however, is clearly a reason to move to a model that accounts for overdispersion.

### c. Let us use a likelihood ratio test to compare these two and test this model assumption

“As we mentioned earlier, negative binomial models assume the conditional means are not equal to the conditional variances. This inequality is captured by estimating a dispersion parameter (not shown in the output) that is held constant in a Poisson model. Thus, the Poisson model is actually nested in the negative binomial model. We can then use a likelihood ratio test to compare these two and test this model assumption.”

https://stats.idre.ucla.edu/r/dae/negative-binomial-regression/

```
pchisq(2 * (logLik(model_2) - logLik(model1)), df = 1, lower.tail = FALSE)
```

```
## 'log Lik.' 0 (df=7)
```

```
library("lmtest")
lrtest(model1, model_2)
```

```
## Likelihood ratio test
## 
## Model 1: Diff_Sex_Partners_N ~ Sex * scale(max_age_same_N, scale = FALSE) + 
##     Sex * I(scale((max_age_same_N), scale = FALSE)^2)
## Model 2: Diff_Sex_Partners_N ~ Sex * scale(max_age_same_N, scale = FALSE) + 
##     Sex * I(scale(max_age_same_N, scale = FALSE)^2)
##   #Df   LogLik Df   Chisq Pr(>Chisq)    
## 1   6 -1745815                          
## 2   7 -1157140  1 1177349  < 2.2e-16 ***
## ---
## Signif. codes:  0 '***' 0.001 '**' 0.01 '*' 0.05 '.' 0.1 ' ' 1
```

In this example the associated chi-squared value estimated from 2\*(logLik(m1) – logLik(m3)) is 926.03 with one degree of freedom. This strongly suggests the negative binomial model, estimating the dispersion parameter, is more appropriate than the Poisson model.

#### C) Outlier removal 3 \* IQR

##### Distribution of Diff\_Sex\_Partners\_N

```
hist(Sexual_Beh_DT_na_outliers_DT$Diff_Sex_Partners_N)
```

```
length(Sexual_Beh_DT_na_outliers_DT$Diff_Sex_Partners_N)
```

```
## [1] 452055
```

```
summary(Sexual_Beh_DT_na_outliers_DT$Diff_Sex_Partners_N)
```

```
##    Min. 1st Qu.  Median    Mean 3rd Qu.    Max.    NA's 
##   0.000   1.000   3.000   4.012   5.000  21.000   19497
```

#### Regressions

##### 1. Poisson Regression Model

Poisson regression is often used for modeling count data.

Assumption : conditional variance is equal to the conditional mean -> test overdispersion

```
model1 <- glm(Diff_Sex_Partners_N ~ Sex * scale(max_age_same_N, scale = FALSE) + Sex * I(scale((max_age_same_N), scale = FALSE)^2), 
              data = Sexual_Beh_DT_na_outliers_DT, family = "poisson")
summary(model1)
```

```
## 
## Call:
## glm(formula = Diff_Sex_Partners_N ~ Sex * scale(max_age_same_N, 
##     scale = FALSE) + Sex * I(scale((max_age_same_N), scale = FALSE)^2), 
##     family = "poisson", data = Sexual_Beh_DT_na_outliers_DT)
## 
## Deviance Residuals: 
##     Min       1Q   Median       3Q      Max  
## -3.8128  -1.5773  -0.8704   0.7554   6.3482  
## 
## Coefficients:
##                                                   Estimate Std. Error z value
## (Intercept)                                      1.431e+00  1.129e-03 1267.44
## Sex                                             -2.230e-01  2.259e-03  -98.71
## scale(max_age_same_N, scale = FALSE)             2.545e-02  4.198e-04   60.62
## I(scale((max_age_same_N), scale = FALSE)^2)     -3.396e-04  7.763e-06  -43.74
## Sex:scale(max_age_same_N, scale = FALSE)         1.589e-02  8.396e-04   18.93
## Sex:I(scale((max_age_same_N), scale = FALSE)^2) -1.611e-04  1.553e-05  -10.38
##                                                 Pr(>|z|)    
## (Intercept)                                       <2e-16 ***
## Sex                                               <2e-16 ***
## scale(max_age_same_N, scale = FALSE)              <2e-16 ***
## I(scale((max_age_same_N), scale = FALSE)^2)       <2e-16 ***
## Sex:scale(max_age_same_N, scale = FALSE)          <2e-16 ***
## Sex:I(scale((max_age_same_N), scale = FALSE)^2)   <2e-16 ***
## ---
## Signif. codes:  0 '***' 0.001 '**' 0.01 '*' 0.05 '.' 0.1 ' ' 1
## 
## (Dispersion parameter for poisson family taken to be 1)
## 
##     Null deviance: 1725710  on 432557  degrees of freedom
## Residual deviance: 1685364  on 432552  degrees of freedom
##   (19497 observations deleted due to missingness)
## AIC: 2847034
## 
## Number of Fisher Scoring iterations: 5
```

Our residual deviance is 19022 for 14440 degrees of freedom. The rule of thumb is ratio = 1, here : 19022/14440 = 1.317313 - So we have moderate dispersion, which we can also test with a dispersion test

```
library("AER")
dispersiontest(model1)
```

```
## 
##  Overdispersion test
## 
## data:  model1
## z = 238.64, p-value < 2.2e-16
## alternative hypothesis: true dispersion is greater than 1
## sample estimates:
## dispersion 
##   4.530123
```

##### 3. “Fixing” overdispersion

### a. by using quasi-families regression

quasi-families augment the normal families by adding a dispersion parameter http://biometry.github.io/APES//LectureNotes/2016-JAGS/Overdispersion/OverdispersionJAGS.html

```
model <- glm(Diff_Sex_Partners_N~ Sex*max_age_same_N + Sex*I(max_age_same_N^2), data = Sexual_Beh_DT_na_outliers_DT, family="quasipoisson", na.action = na.exclude)
summary(model)
```

```
## 
## Call:
## glm(formula = Diff_Sex_Partners_N ~ Sex * max_age_same_N + Sex * 
##     I(max_age_same_N^2), family = "quasipoisson", data = Sexual_Beh_DT_na_outliers_DT, 
##     na.action = na.exclude)
## 
## Deviance Residuals: 
##     Min       1Q   Median       3Q      Max  
## -3.8128  -1.5773  -0.8704   0.7554   6.3482  
## 
## Coefficients:
##                           Estimate Std. Error t value Pr(>|t|)    
## (Intercept)              1.406e+00  1.852e-03 759.078  < 2e-16 ***
## Sex                     -2.390e-01  3.703e-03 -64.538  < 2e-16 ***
## max_age_same_N           2.613e-02  9.261e-04  28.212  < 2e-16 ***
## I(max_age_same_N^2)     -3.396e-04  1.652e-05 -20.550  < 2e-16 ***
## Sex:max_age_same_N       1.622e-02  1.852e-03   8.755  < 2e-16 ***
## Sex:I(max_age_same_N^2) -1.611e-04  3.305e-05  -4.876 1.08e-06 ***
## ---
## Signif. codes:  0 '***' 0.001 '**' 0.01 '*' 0.05 '.' 0.1 ' ' 1
## 
## (Dispersion parameter for quasipoisson family taken to be 4.530229)
## 
##     Null deviance: 1725710  on 432557  degrees of freedom
## Residual deviance: 1685364  on 432552  degrees of freedom
##   (19497 observations deleted due to missingness)
## AIC: NA
## 
## Number of Fisher Scoring iterations: 5
```

You see that τ (dispersion parameter) is estimated at a value similar to those in the overdispersion tests above (as you’d expect). The main effect is the substantially larger errors for the estimates (the point estimates do not change), and hence potentially changed significances (though not here). (You can manually compute the corrected standard errors as Poisson-standard errors ·√τ .) Note that because this is no maximum likelihoo method (but a quasi-likelihood method), no likelihood and hence no AIC are available. No overdispersion tests can be conducted for quasi-family objects (neither in AER nor DHARMa)

### b. by using negative binomial regression

Conditional variance exceedes the conditional mean -> test overdispersion Maybe our distributional assumption was simply wrong, and we choose a different distribution

https://biometry.github.io/APES/LectureNotes/2016-JAGS/Overdispersion/OverdispersionJAGS.pdf https://stats.idre.ucla.edu/r/dae/negative-binomial-regression/

```
library(MASS)
model_2 <- glm.nb(Diff_Sex_Partners_N~ Sex*scale(max_age_same_N, scale = FALSE)+ Sex*I(scale(max_age_same_N, scale = FALSE)^2), data = Sexual_Beh_DT_na_outliers_DT, na.action = na.exclude)
summary(model_2)
```

```
## 
## Call:
## glm.nb(formula = Diff_Sex_Partners_N ~ Sex * scale(max_age_same_N, 
##     scale = FALSE) + Sex * I(scale(max_age_same_N, scale = FALSE)^2), 
##     data = Sexual_Beh_DT_na_outliers_DT, na.action = na.exclude, 
##     init.theta = 1.304803171, link = log)
## 
## Deviance Residuals: 
##     Min       1Q   Median       3Q      Max  
## -2.2144  -0.9375  -0.4838   0.3766   2.6043  
## 
## Coefficients:
##                                                 Estimate Std. Error z value
## (Intercept)                                    1.431e+00  2.513e-03 569.315
## Sex                                           -2.236e-01  5.027e-03 -44.487
## scale(max_age_same_N, scale = FALSE)           2.522e-02  9.902e-04  25.469
## I(scale(max_age_same_N, scale = FALSE)^2)     -3.351e-04  1.812e-05 -18.497
## Sex:scale(max_age_same_N, scale = FALSE)       1.558e-02  1.980e-03   7.868
## Sex:I(scale(max_age_same_N, scale = FALSE)^2) -1.549e-04  3.623e-05  -4.275
##                                               Pr(>|z|)    
## (Intercept)                                    < 2e-16 ***
## Sex                                            < 2e-16 ***
## scale(max_age_same_N, scale = FALSE)           < 2e-16 ***
## I(scale(max_age_same_N, scale = FALSE)^2)      < 2e-16 ***
## Sex:scale(max_age_same_N, scale = FALSE)      3.60e-15 ***
## Sex:I(scale(max_age_same_N, scale = FALSE)^2) 1.91e-05 ***
## ---
## Signif. codes:  0 '***' 0.001 '**' 0.01 '*' 0.05 '.' 0.1 ' ' 1
## 
## (Dispersion parameter for Negative Binomial(1.3048) family taken to be 1)
## 
##     Null deviance: 475325  on 432557  degrees of freedom
## Residual deviance: 465745  on 432552  degrees of freedom
##   (19497 observations deleted due to missingness)
## AIC: 2150463
## 
## Number of Fisher Scoring iterations: 1
## 
## 
##               Theta:  1.30480 
##           Std. Err.:  0.00379 
## 
##  2 x log-likelihood:  -2150449.04500
```

```
1 - pchisq(summary(model_2)$deviance,
         summary(model_2)$df.residual)
```

```
## [1] 0
```

https://www.r-bloggers.com/2012/08/count-data-and-glms-choosing-among-poisson-negative-binomial-and-zero-inflated-models/ https://stats.stackexchange.com/questions/37732/when-someone-says-residual-deviance-df-should-1-for-a-poisson-model-how-appro

The ratio of deviance 13067/14994 = 0.8714819. We can also check with a dispersion test.

### `{r} # library(DHARMa) # simulationOutput <- simulateResiduals(model_2) # testDispersion(simulationOutput) # testZeroInflation(simulationOutput) #`

Error in securityAssertion(“Simulation from the model produced wrong dimension”, Message from DHARMa: During the execution of a DHARMa function, some unexpected conditions occurred. Even if you didnt get an error, your results may not be reliable. Please check with the help if you use the functions as intended. If you think that the error is not on your side, I would be grateful if you could report the problem at https://github.com/florianhartig/DHARMa/issues Context: Simulation from the model produced wrong dimension https://stats.stackexchange.com/questions/490680/significant-dispersion-test

```
                       A word of warning that applies also to all other tests that follow: significance in hypothesis tests depends on at least 2 ingredients: strength of the signal, and number of data points. Hence, the p-value alone is not a good indicator of the extent to which your residuals deviate from assumptions. Specifically, if you have a lot of data points, residual diagnostics will nearly inevitably become significant, because having a perfectly fitting model is very unlikely. That, however, doesn’t necessarily mean that you need to change your model. The p-values confirm that there is a deviation from your null hypothesis. It is, however, in your discretion to decide whether this deviation is worth worrying about. If you see a dispersion parameter of 1.01, I would not worry, even if the test is significant. A significant value of 5, however, is clearly a reason to move to a model that accounts for overdispersion.
```

### c. Let us use a likelihood ratio test to compare these two and test this model assumption

```
                       "As we mentioned earlier, negative binomial models assume the conditional means are not equal to the conditional variances. This inequality is captured by estimating a dispersion parameter (not shown in the output) that is held constant in a Poisson model. Thus, the Poisson model is actually nested in the negative binomial model. We can then use a likelihood ratio test to compare these two and test this model assumption." 
                       
                       https://stats.idre.ucla.edu/r/dae/negative-binomial-regression/
```
