## Supplementary material for "High Intelligence is not a Risk Factor for Mental Health Disorders": Supplemental_File_S4_Same_Sex_N_Sex_Age.html


### Same\_sex\_N

### Same\_Sex\_Partners\_N

#### a) No outlier removal

#### Distribution of Same\_Sex\_Partners\_N

```
hist(Sexual_Beh_DF$Same_Sex_Partners_N)
```

```
length(Sexual_Beh_DF$Same_Sex_Partners_N)
```

```
## [1] 502120
```

```
summary(Sexual_Beh_DF$Same_Sex_Partners_N)
```

```
##     Min.  1st Qu.   Median     Mean  3rd Qu.     Max. 
## -1.0e+00 -1.0e+00 -1.0e+00 -8.1e-02 -1.0e+00  1.5e+04
```

##### 1. Poisson Regression Model

Poisson regression is often used for modeling count data.

Assumption : conditional variance is equal to the conditional mean -> test overdispersion

```
Sexual_Beh_DF_no_NA <- dplyr::filter(Sexual_Beh_DF_no_NA,Same_Sex_Partners_N != -1 )
model1 <- glm(Same_Sex_Partners_N ~ Sex * scale(max_age_same_N, scale = FALSE) + Sex * I(scale((max_age_same_N), scale = FALSE)^2), 
                             data = Sexual_Beh_DF_no_NA, family = "poisson")
summary(model1)
```

```
## 
## Call:
## glm(formula = Same_Sex_Partners_N ~ Sex * scale(max_age_same_N, 
##     scale = FALSE) + Sex * I(scale((max_age_same_N), scale = FALSE)^2), 
##     family = "poisson", data = Sexual_Beh_DF_no_NA)
## 
## Deviance Residuals: 
##    Min      1Q  Median      3Q     Max  
## -10.82   -8.65   -2.49   -0.82  369.45  
## 
## Coefficients:
##                                                   Estimate Std. Error  z value
## (Intercept)                                      2.570e+00  4.864e-03  528.283
## Sex                                             -2.904e+00  9.729e-03 -298.473
## scale(max_age_same_N, scale = FALSE)            -2.046e-02  4.435e-04  -46.132
## I(scale((max_age_same_N), scale = FALSE)^2)     -1.402e-03  5.143e-05  -27.255
## Sex:scale(max_age_same_N, scale = FALSE)         5.398e-03  8.869e-04    6.087
## Sex:I(scale((max_age_same_N), scale = FALSE)^2)  2.735e-03  1.029e-04   26.594
##                                                 Pr(>|z|)    
## (Intercept)                                      < 2e-16 ***
## Sex                                              < 2e-16 ***
## scale(max_age_same_N, scale = FALSE)             < 2e-16 ***
## I(scale((max_age_same_N), scale = FALSE)^2)      < 2e-16 ***
## Sex:scale(max_age_same_N, scale = FALSE)        1.15e-09 ***
## Sex:I(scale((max_age_same_N), scale = FALSE)^2)  < 2e-16 ***
## ---
## Signif. codes:  0 '***' 0.001 '**' 0.01 '*' 0.05 '.' 0.1 ' ' 1
## 
## (Dispersion parameter for poisson family taken to be 1)
## 
##     Null deviance: 2762586  on 16279  degrees of freedom
## Residual deviance: 2383909  on 16274  degrees of freedom
## AIC: 2427126
## 
## Number of Fisher Scoring iterations: 8
```

Our residual deviance is 2015675 for 16274 degrees of freedom. The rule of thumb is ratio = 1, here : 2015675/16274 = 123.858 - So we have moderate dispersion, which we can also test with a dispersion test

```
library("AER")
```

```
## Loading required package: car
```

```
## Loading required package: carData
```

```
## Registered S3 methods overwritten by 'car':
##   method                          from
##   influence.merMod                lme4
##   cooks.distance.influence.merMod lme4
##   dfbeta.influence.merMod         lme4
##   dfbetas.influence.merMod        lme4
```

```
## 
## Attaching package: 'car'
```

```
## The following object is masked from 'package:dplyr':
## 
##     recode
```

```
## Loading required package: lmtest
```

```
## Loading required package: zoo
```

```
## 
## Attaching package: 'zoo'
```

```
## The following objects are masked from 'package:base':
## 
##     as.Date, as.Date.numeric
```

```
## Loading required package: survival
```

```
dispersiontest(model1)
```

```
## 
##  Overdispersion test
## 
## data:  model1
## z = 4.0056, p-value = 3.093e-05
## alternative hypothesis: true dispersion is greater than 1
## sample estimates:
## dispersion 
##   1297.169
```

##### 2. “Fixing” overdispersion

###### a. by using quasi-families regression

quasi-families augment the normal families by adding a dispersion parameter http://biometry.github.io/APES//LectureNotes/2016-JAGS/Overdispersion/OverdispersionJAGS.html

```
model <- glm(Same_Sex_Partners_N~ Sex*max_age_same_N + Sex*I(max_age_same_N^2), data = Sexual_Beh_DF_no_NA, family="quasipoisson", na.action = na.exclude)
summary(model)
```

```
## 
## Call:
## glm(formula = Same_Sex_Partners_N ~ Sex * max_age_same_N + Sex * 
##     I(max_age_same_N^2), family = "quasipoisson", data = Sexual_Beh_DF_no_NA, 
##     na.action = na.exclude)
## 
## Deviance Residuals: 
##    Min      1Q  Median      3Q     Max  
## -10.82   -8.65   -2.49   -0.82  369.45  
## 
## Coefficients:
##                          Estimate Std. Error t value Pr(>|t|)
## (Intercept)             -0.481939   5.350358  -0.090    0.928
## Sex                      4.933929  10.700716   0.461    0.645
## max_age_same_N           0.132400   0.200737   0.660    0.510
## I(max_age_same_N^2)     -0.001402   0.001853  -0.757    0.449
## Sex:max_age_same_N      -0.292897   0.401475  -0.730    0.466
## Sex:I(max_age_same_N^2)  0.002735   0.003705   0.738    0.460
## 
## (Dispersion parameter for quasipoisson family taken to be 1297.668)
## 
##     Null deviance: 2762586  on 16279  degrees of freedom
## Residual deviance: 2383909  on 16274  degrees of freedom
## AIC: NA
## 
## Number of Fisher Scoring iterations: 8
```

You see that τ (dispersion parameter) is estimated at a value similar to those in the overdispersion tests above (as you’d expect). The main effect is the substantially larger errors for the estimates (the point estimates do not change), and hence potentially changed significances (though not here). (You can manually compute the corrected standard errors as Poisson-standard errors ·√τ .) Note that because this is no maximum likelihoo method (but a quasi-likelihood method), no likelihood and hence no AIC are available. No overdispersion tests can be conducted for quasi-family objects (neither in AER nor DHARMa)

###### b. by using negative binomial regression

Conditional variance exceedes the conditional mean -> test overdispersion Maybe our distributional assumption was simply wrong, and we choose a different distribution

https://biometry.github.io/APES/LectureNotes/2016-JAGS/Overdispersion/OverdispersionJAGS.pdf https://stats.idre.ucla.edu/r/dae/negative-binomial-regression/

```
library(MASS)
```

```
## 
## Attaching package: 'MASS'
```

```
## The following object is masked from 'package:dplyr':
## 
##     select
```

```
model_2 <- glm.nb(Same_Sex_Partners_N~ Sex*scale(max_age_same_N, scale = FALSE)+ Sex*I(scale(max_age_same_N, scale = FALSE)^2), data = Sexual_Beh_DF_no_NA, na.action = na.exclude)
summary(model_2)
```

```
## 
## Call:
## glm.nb(formula = Same_Sex_Partners_N ~ Sex * scale(max_age_same_N, 
##     scale = FALSE) + Sex * I(scale(max_age_same_N, scale = FALSE)^2), 
##     data = Sexual_Beh_DF_no_NA, na.action = na.exclude, init.theta = 0.3037063667, 
##     link = log)
## 
## Deviance Residuals: 
##     Min       1Q   Median       3Q      Max  
## -1.7795  -1.1913  -0.5477  -0.2220  12.7794  
## 
## Coefficients:
##                                                 Estimate Std. Error z value
## (Intercept)                                    2.5233632  0.0205166 122.991
## Sex                                           -2.8084939  0.0410332 -68.445
## scale(max_age_same_N, scale = FALSE)          -0.0200048  0.0018136 -11.030
## I(scale(max_age_same_N, scale = FALSE)^2)     -0.0007243  0.0002021  -3.584
## Sex:scale(max_age_same_N, scale = FALSE)       0.0044695  0.0036273   1.232
## Sex:I(scale(max_age_same_N, scale = FALSE)^2)  0.0013431  0.0004042   3.323
##                                               Pr(>|z|)    
## (Intercept)                                    < 2e-16 ***
## Sex                                            < 2e-16 ***
## scale(max_age_same_N, scale = FALSE)           < 2e-16 ***
## I(scale(max_age_same_N, scale = FALSE)^2)     0.000339 ***
## Sex:scale(max_age_same_N, scale = FALSE)      0.217881    
## Sex:I(scale(max_age_same_N, scale = FALSE)^2) 0.000891 ***
## ---
## Signif. codes:  0 '***' 0.001 '**' 0.01 '*' 0.05 '.' 0.1 ' ' 1
## 
## (Dispersion parameter for Negative Binomial(0.3037) family taken to be 1)
## 
##     Null deviance: 25346  on 16279  degrees of freedom
## Residual deviance: 18864  on 16274  degrees of freedom
## AIC: 102660
## 
## Number of Fisher Scoring iterations: 1
## 
## 
##               Theta:  0.30371 
##           Std. Err.:  0.00308 
## 
##  2 x log-likelihood:  -102645.83300
```

```
1 - pchisq(summary(model_2)$deviance,
           summary(model_2)$df.residual)
```

```
## [1] 0
```

https://www.r-bloggers.com/2012/08/count-data-and-glms-choosing-among-poisson-negative-binomial-and-zero-inflated-models/ https://stats.stackexchange.com/questions/37732/when-someone-says-residual-deviance-df-should-1-for-a-poisson-model-how-appro

The ratio of deviance 17744/16274 = 1.09 and hence probably fine. We can also check with a dispersion test.

```
library(DHARMa)
```

```
## This is DHARMa 0.4.1. For overview type '?DHARMa'. For recent changes, type news(package = 'DHARMa') Note: Syntax of plotResiduals has changed in 0.3.0, see ?plotResiduals for details
```

```
simulationOutput <- simulateResiduals(model_2)
testDispersion(simulationOutput)
```

```
## 
##  DHARMa nonparametric dispersion test via sd of residuals fitted vs.
##  simulated
## 
## data:  simulationOutput
## dispersion = 15.76, p-value < 2.2e-16
## alternative hypothesis: two.sided
```

```
testZeroInflation(simulationOutput)
```

```
## 
##  DHARMa zero-inflation test via comparison to expected zeros with
##  simulation under H0 = fitted model
## 
## data:  simulationOutput
## ratioObsSim = 0.46777, p-value < 2.2e-16
## alternative hypothesis: two.sided
```

https://stats.stackexchange.com/questions/490680/significant-dispersion-test

A word of warning that applies also to all other tests that follow: significance in hypothesis tests depends on at least 2 ingredients: strength of the signal, and number of data points. Hence, the p-value alone is not a good indicator of the extent to which your residuals deviate from assumptions. Specifically, if you have a lot of data points, residual diagnostics will nearly inevitably become significant, because having a perfectly fitting model is very unlikely. That, however, doesn’t necessarily mean that you need to change your model. The p-values confirm that there is a deviation from your null hypothesis. It is, however, in your discretion to decide whether this deviation is worth worrying about. If you see a dispersion parameter of 1.01, I would not worry, even if the test is significant. A significant value of 5, however, is clearly a reason to move to a model that accounts for overdispersion.

###### c. Let us use a likelihood ratio test to compare these two and test this model assumption

“As we mentioned earlier, negative binomial models assume the conditional means are not equal to the conditional variances. This inequality is captured by estimating a dispersion parameter (not shown in the output) that is held constant in a Poisson model. Thus, the Poisson model is actually nested in the negative binomial model. We can then use a likelihood ratio test to compare these two and test this model assumption.”

https://stats.idre.ucla.edu/r/dae/negative-binomial-regression/

```
pchisq(2 * (logLik(model_2) - logLik(model1)), df = 1, lower.tail = FALSE)
```

```
## 'log Lik.' 0 (df=7)
```

```
library("lmtest")
lrtest(model1, model_2)
```

```
## Likelihood ratio test
## 
## Model 1: Same_Sex_Partners_N ~ Sex * scale(max_age_same_N, scale = FALSE) + 
##     Sex * I(scale((max_age_same_N), scale = FALSE)^2)
## Model 2: Same_Sex_Partners_N ~ Sex * scale(max_age_same_N, scale = FALSE) + 
##     Sex * I(scale(max_age_same_N, scale = FALSE)^2)
##   #Df   LogLik Df   Chisq Pr(>Chisq)    
## 1   6 -1213557                          
## 2   7   -51323  1 2324469  < 2.2e-16 ***
## ---
## Signif. codes:  0 '***' 0.001 '**' 0.01 '*' 0.05 '.' 0.1 ' ' 1
```

In this example the associated chi-squared value estimated from 2\*(logLik(m1) – logLik(m3)) is 926.03 with one degree of freedom. This strongly suggests the negative binomial model, estimating the dispersion parameter, is more appropriate than the Poisson model.

#### B) Outlier removal 6 \* IQR

##### Distribution of Same\_Sex\_Partners\_N

```
hist(Sexual_Beh_DT_na_outliers_DT$Same_Sex_Partners_N)
```

```
length(Sexual_Beh_DT_na_outliers_DT$Same_Sex_Partners_N)
```

```
## [1] 16280
```

```
summary(Sexual_Beh_DT_na_outliers_DT$Same_Sex_Partners_N)
```

```
##    Min. 1st Qu.  Median    Mean 3rd Qu.    Max.    NA's 
##   0.000   1.000   1.000   3.247   4.000  29.000    1271
```

##### 1. Poisson Regression Model

Poisson regression is often used for modeling count data.

Assumption : conditional variance is equal to the conditional mean -> test overdispersion

```
model1 <- glm(Same_Sex_Partners_N ~ Sex * scale(max_age_same_N, scale = FALSE) + Sex * I(scale((max_age_same_N), scale = FALSE)^2), 
              data = Sexual_Beh_DT_na_outliers_DT, family = "poisson")
summary(model1)
```

```
## 
## Call:
## glm(formula = Same_Sex_Partners_N ~ Sex * scale(max_age_same_N, 
##     scale = FALSE) + Sex * I(scale((max_age_same_N), scale = FALSE)^2), 
##     family = "poisson", data = Sexual_Beh_DT_na_outliers_DT)
## 
## Deviance Residuals: 
##     Min       1Q   Median       3Q      Max  
## -3.0079  -1.5512  -0.9999   0.2817   9.2975  
## 
## Coefficients:
##                                                   Estimate Std. Error z value
## (Intercept)                                      1.152e+00  6.546e-03 176.061
## Sex                                             -3.629e-01  1.309e-02 -27.722
## scale(max_age_same_N, scale = FALSE)            -1.420e-02  5.772e-04 -24.598
## I(scale((max_age_same_N), scale = FALSE)^2)     -3.897e-05  6.591e-05  -0.591
## Sex:scale(max_age_same_N, scale = FALSE)        -7.327e-04  1.154e-03  -0.635
## Sex:I(scale((max_age_same_N), scale = FALSE)^2)  1.293e-04  1.318e-04   0.981
##                                                 Pr(>|z|)    
## (Intercept)                                       <2e-16 ***
## Sex                                               <2e-16 ***
## scale(max_age_same_N, scale = FALSE)              <2e-16 ***
## I(scale((max_age_same_N), scale = FALSE)^2)        0.554    
## Sex:scale(max_age_same_N, scale = FALSE)           0.526    
## Sex:I(scale((max_age_same_N), scale = FALSE)^2)    0.327    
## ---
## Signif. codes:  0 '***' 0.001 '**' 0.01 '*' 0.05 '.' 0.1 ' ' 1
## 
## (Dispersion parameter for poisson family taken to be 1)
## 
##     Null deviance: 69567  on 15008  degrees of freedom
## Residual deviance: 67701  on 15003  degrees of freedom
##   (1271 observations deleted due to missingness)
## AIC: 102606
## 
## Number of Fisher Scoring iterations: 6
```

Our residual deviance is 35060 for 14994 degrees of freedom. The rule of thumb is ratio = 1, here : 35060/14994 = 2.338269 - So we have moderate dispersion, which we can also test with a dispersion test

```
library("AER")
dispersiontest(model1)
```

```
## 
##  Overdispersion test
## 
## data:  model1
## z = 35.357, p-value < 2.2e-16
## alternative hypothesis: true dispersion is greater than 1
## sample estimates:
## dispersion 
##   6.266942
```

##### 2. “Fixing” overdispersion

###### a. by using quasi-families regression

quasi-families augment the normal families by adding a dispersion parameter http://biometry.github.io/APES//LectureNotes/2016-JAGS/Overdispersion/OverdispersionJAGS.html

```
model <- glm(Same_Sex_Partners_N~ Sex*max_age_same_N + Sex*I(max_age_same_N^2), data = Sexual_Beh_DT_na_outliers_DT, family="quasipoisson", na.action = na.exclude)
summary(model)
```

```
## 
## Call:
## glm(formula = Same_Sex_Partners_N ~ Sex * max_age_same_N + Sex * 
##     I(max_age_same_N^2), family = "quasipoisson", data = Sexual_Beh_DT_na_outliers_DT, 
##     na.action = na.exclude)
## 
## Deviance Residuals: 
##     Min       1Q   Median       3Q      Max  
## -3.0079  -1.5512  -0.9999   0.2817   9.2975  
## 
## Coefficients:
##                           Estimate Std. Error t value Pr(>|t|)    
## (Intercept)              1.811e+00  4.872e-01   3.717 0.000203 ***
## Sex                      6.148e-02  9.744e-01   0.063 0.949691    
## max_age_same_N          -9.950e-03  1.808e-02  -0.550 0.582174    
## I(max_age_same_N^2)     -3.897e-05  1.650e-04  -0.236 0.813338    
## Sex:max_age_same_N      -1.483e-02  3.617e-02  -0.410 0.681667    
## Sex:I(max_age_same_N^2)  1.293e-04  3.300e-04   0.392 0.695187    
## ---
## Signif. codes:  0 '***' 0.001 '**' 0.01 '*' 0.05 '.' 0.1 ' ' 1
## 
## (Dispersion parameter for quasipoisson family taken to be 6.269457)
## 
##     Null deviance: 69567  on 15008  degrees of freedom
## Residual deviance: 67701  on 15003  degrees of freedom
##   (1271 observations deleted due to missingness)
## AIC: NA
## 
## Number of Fisher Scoring iterations: 6
```

You see that τ (dispersion parameter) is estimated at a value similar to those in the overdispersion tests above (as you’d expect). The main effect is the substantially larger errors for the estimates (the point estimates do not change), and hence potentially changed significances (though not here). (You can manually compute the corrected standard errors as Poisson-standard errors ·√τ .) Note that because this is no maximum likelihoo method (but a quasi-likelihood method), no likelihood and hence no AIC are available. No overdispersion tests can be conducted for quasi-family objects (neither in AER nor DHARMa)

###### b. by using negative binomial regression

Conditional variance exceedes the conditional mean -> test overdispersion Maybe our distributional assumption was simply wrong, and we choose a different distribution

https://biometry.github.io/APES/LectureNotes/2016-JAGS/Overdispersion/OverdispersionJAGS.pdf https://stats.idre.ucla.edu/r/dae/negative-binomial-regression/

```
library(MASS)
model_2 <- glm.nb(Same_Sex_Partners_N~ Sex*scale(max_age_same_N, scale = FALSE)+ Sex*I(scale(max_age_same_N, scale = FALSE)^2), data = Sexual_Beh_DT_na_outliers_DT, na.action = na.exclude)
summary(model_2)
```

```
## 
## Call:
## glm.nb(formula = Same_Sex_Partners_N ~ Sex * scale(max_age_same_N, 
##     scale = FALSE) + Sex * I(scale(max_age_same_N, scale = FALSE)^2), 
##     data = Sexual_Beh_DT_na_outliers_DT, na.action = na.exclude, 
##     init.theta = 0.9162918089, link = log)
## 
## Deviance Residuals: 
##     Min       1Q   Median       3Q      Max  
## -1.8078  -0.8217  -0.5526   0.1278   3.5154  
## 
## Coefficients:
##                                                 Estimate Std. Error z value
## (Intercept)                                    1.152e+00  1.355e-02  84.961
## Sex                                           -3.622e-01  2.711e-02 -13.361
## scale(max_age_same_N, scale = FALSE)          -1.422e-02  1.200e-03 -11.844
## I(scale(max_age_same_N, scale = FALSE)^2)     -2.703e-05  1.336e-04  -0.202
## Sex:scale(max_age_same_N, scale = FALSE)      -6.932e-04  2.401e-03  -0.289
## Sex:I(scale(max_age_same_N, scale = FALSE)^2)  1.203e-04  2.672e-04   0.450
##                                               Pr(>|z|)    
## (Intercept)                                     <2e-16 ***
## Sex                                             <2e-16 ***
## scale(max_age_same_N, scale = FALSE)            <2e-16 ***
## I(scale(max_age_same_N, scale = FALSE)^2)        0.840    
## Sex:scale(max_age_same_N, scale = FALSE)         0.773    
## Sex:I(scale(max_age_same_N, scale = FALSE)^2)    0.653    
## ---
## Signif. codes:  0 '***' 0.001 '**' 0.01 '*' 0.05 '.' 0.1 ' ' 1
## 
## (Dispersion parameter for Negative Binomial(0.9163) family taken to be 1)
## 
##     Null deviance: 16251  on 15008  degrees of freedom
## Residual deviance: 15838  on 15003  degrees of freedom
##   (1271 observations deleted due to missingness)
## AIC: 69118
## 
## Number of Fisher Scoring iterations: 1
## 
## 
##               Theta:  0.9163 
##           Std. Err.:  0.0137 
## 
##  2 x log-likelihood:  -69104.2510
```

```
1 - pchisq(summary(model_2)$deviance,
           summary(model_2)$df.residual)
```

```
## [1] 1.086696e-06
```

https://www.r-bloggers.com/2012/08/count-data-and-glms-choosing-among-poisson-negative-binomial-and-zero-inflated-models/ https://stats.stackexchange.com/questions/37732/when-someone-says-residual-deviance-df-should-1-for-a-poisson-model-how-appro

https://stats.idre.ucla.edu/r/dae/negative-binomial-regression/

```
pchisq(2 * (logLik(model_2) - logLik(model1)), df = 1, lower.tail = FALSE)
```

```
## 'log Lik.' 0 (df=7)
```

```
library("lmtest")
lrtest(model1, model_2)
```

```
## Likelihood ratio test
## 
## Model 1: Same_Sex_Partners_N ~ Sex * scale(max_age_same_N, scale = FALSE) + 
##     Sex * I(scale((max_age_same_N), scale = FALSE)^2)
## Model 2: Same_Sex_Partners_N ~ Sex * scale(max_age_same_N, scale = FALSE) + 
##     Sex * I(scale(max_age_same_N, scale = FALSE)^2)
##   #Df LogLik Df Chisq Pr(>Chisq)    
## 1   6 -51297                        
## 2   7 -34552  1 33490  < 2.2e-16 ***
## ---
## Signif. codes:  0 '***' 0.001 '**' 0.01 '*' 0.05 '.' 0.1 ' ' 1
```

In this example the associated chi-squared value estimated from 2\*(logLik(m1) – logLik(m3)) is 926.03 with one degree of freedom. This strongly suggests the negative binomial model, estimating the dispersion parameter, is more appropriate than the Poisson model.

#### C) Outlier removal 3 \* IQR

##### Distribution of Same\_Sex\_Partners\_N

```
hist(Sexual_Beh_DT_na_outliers_DT$Same_Sex_Partners_N)
```

```
length(Sexual_Beh_DT_na_outliers_DT$Same_Sex_Partners_N)
```

```
## [1] 16280
```

```
summary(Sexual_Beh_DT_na_outliers_DT$Same_Sex_Partners_N)
```

```
##    Min. 1st Qu.  Median    Mean 3rd Qu.    Max.    NA's 
##   0.000   1.000   1.000   2.564   3.000  17.000    1810
```

##### 1. Poisson Regression Model

Poisson regression is often used for modeling count data.

Assumption : conditional variance is equal to the conditional mean -> test overdispersion

```
model1 <- glm(Same_Sex_Partners_N ~ Sex * scale(max_age_same_N, scale = FALSE) + Sex * I(scale((max_age_same_N), scale = FALSE)^2), 
              data = Sexual_Beh_DT_na_outliers_DT, family = "poisson")
summary(model1)
```

```
## 
## Call:
## glm(formula = Same_Sex_Partners_N ~ Sex * scale(max_age_same_N, 
##     scale = FALSE) + Sex * I(scale((max_age_same_N), scale = FALSE)^2), 
##     family = "poisson", data = Sexual_Beh_DT_na_outliers_DT)
## 
## Deviance Residuals: 
##     Min       1Q   Median       3Q      Max  
## -2.5041  -1.2110  -0.8824   0.3747   6.3347  
## 
## Coefficients:
##                                                   Estimate Std. Error z value
## (Intercept)                                      9.448e-01  7.312e-03 129.205
## Sex                                             -7.720e-02  1.462e-02  -5.279
## scale(max_age_same_N, scale = FALSE)            -1.192e-02  6.428e-04 -18.540
## I(scale((max_age_same_N), scale = FALSE)^2)     -9.847e-05  7.318e-05  -1.346
## Sex:scale(max_age_same_N, scale = FALSE)         1.584e-03  1.286e-03   1.232
## Sex:I(scale((max_age_same_N), scale = FALSE)^2)  1.181e-05  1.464e-04   0.081
##                                                 Pr(>|z|)    
## (Intercept)                                      < 2e-16 ***
## Sex                                              1.3e-07 ***
## scale(max_age_same_N, scale = FALSE)             < 2e-16 ***
## I(scale((max_age_same_N), scale = FALSE)^2)        0.178    
## Sex:scale(max_age_same_N, scale = FALSE)           0.218    
## Sex:I(scale((max_age_same_N), scale = FALSE)^2)    0.936    
## ---
## Signif. codes:  0 '***' 0.001 '**' 0.01 '*' 0.05 '.' 0.1 ' ' 1
## 
## (Dispersion parameter for poisson family taken to be 1)
## 
##     Null deviance: 42801  on 14469  degrees of freedom
## Residual deviance: 42397  on 14464  degrees of freedom
##   (1810 observations deleted due to missingness)
## AIC: 74656
## 
## Number of Fisher Scoring iterations: 5
```

Our residual deviance is 19022 for 14440 degrees of freedom. The rule of thumb is ratio = 1, here : 19022/14440 = 1.317313 - So we have moderate dispersion, which we can also test with a dispersion test

```
library("AER")
dispersiontest(model1)
```

```
## 
##  Overdispersion test
## 
## data:  model1
## z = 37.19, p-value < 2.2e-16
## alternative hypothesis: true dispersion is greater than 1
## sample estimates:
## dispersion 
##   3.638902
```

##### 2. “Fixing” overdispersion

###### a. by using quasi-families regression

quasi-families augment the normal families by adding a dispersion parameter http://biometry.github.io/APES//LectureNotes/2016-JAGS/Overdispersion/OverdispersionJAGS.html

```
model <- glm(Same_Sex_Partners_N~ Sex*max_age_same_N + Sex*I(max_age_same_N^2), data = Sexual_Beh_DT_na_outliers_DT, family="quasipoisson", na.action = na.exclude)
summary(model)
```

```
## 
## Call:
## glm(formula = Same_Sex_Partners_N ~ Sex * max_age_same_N + Sex * 
##     I(max_age_same_N^2), family = "quasipoisson", data = Sexual_Beh_DT_na_outliers_DT, 
##     na.action = na.exclude)
## 
## Deviance Residuals: 
##     Min       1Q   Median       3Q      Max  
## -2.5041  -1.2110  -0.8824   0.3747   6.3347  
## 
## Coefficients:
##                           Estimate Std. Error t value Pr(>|t|)   
## (Intercept)              1.302e+00  4.154e-01   3.134  0.00173 **
## Sex                     -1.285e-01  8.309e-01  -0.155  0.87711   
## max_age_same_N          -1.180e-03  1.536e-02  -0.077  0.93877   
## I(max_age_same_N^2)     -9.847e-05  1.396e-04  -0.705  0.48069   
## Sex:max_age_same_N       2.967e-04  3.072e-02   0.010  0.99229   
## Sex:I(max_age_same_N^2)  1.181e-05  2.792e-04   0.042  0.96628   
## ---
## Signif. codes:  0 '***' 0.001 '**' 0.01 '*' 0.05 '.' 0.1 ' ' 1
## 
## (Dispersion parameter for quasipoisson family taken to be 3.640659)
## 
##     Null deviance: 42801  on 14469  degrees of freedom
## Residual deviance: 42397  on 14464  degrees of freedom
##   (1810 observations deleted due to missingness)
## AIC: NA
## 
## Number of Fisher Scoring iterations: 5
```

You see that τ (dispersion parameter) is estimated at a value similar to those in the overdispersion tests above (as you’d expect). The main effect is the substantially larger errors for the estimates (the point estimates do not change), and hence potentially changed significances (though not here). (You can manually compute the corrected standard errors as Poisson-standard errors ·√τ .) Note that because this is no maximum likelihoo method (but a quasi-likelihood method), no likelihood and hence no AIC are available. No overdispersion tests can be conducted for quasi-family objects (neither in AER nor DHARMa)

###### b. by using negative binomial regression

Conditional variance exceedes the conditional mean -> test overdispersion Maybe our distributional assumption was simply wrong, and we choose a different distribution

https://biometry.github.io/APES/LectureNotes/2016-JAGS/Overdispersion/OverdispersionJAGS.pdf https://stats.idre.ucla.edu/r/dae/negative-binomial-regression/

```
library(MASS)
model_2 <- glm.nb(Same_Sex_Partners_N~ Sex*scale(max_age_same_N, scale = FALSE)+ Sex*I(scale(max_age_same_N, scale = FALSE)^2), data = Sexual_Beh_DT_na_outliers_DT, na.action = na.exclude)
summary(model_2)
```

```
## 
## Call:
## glm.nb(formula = Same_Sex_Partners_N ~ Sex * scale(max_age_same_N, 
##     scale = FALSE) + Sex * I(scale(max_age_same_N, scale = FALSE)^2), 
##     data = Sexual_Beh_DT_na_outliers_DT, na.action = na.exclude, 
##     init.theta = 1.291697398, link = log)
## 
## Deviance Residuals: 
##     Min       1Q   Median       3Q      Max  
## -1.7832  -0.7581  -0.5804   0.2161   3.0239  
## 
## Coefficients:
##                                                 Estimate Std. Error z value
## (Intercept)                                    9.450e-01  1.253e-02  75.399
## Sex                                           -7.813e-02  2.507e-02  -3.117
## scale(max_age_same_N, scale = FALSE)          -1.191e-02  1.108e-03 -10.745
## I(scale(max_age_same_N, scale = FALSE)^2)     -1.004e-04  1.236e-04  -0.812
## Sex:scale(max_age_same_N, scale = FALSE)       1.570e-03  2.216e-03   0.708
## Sex:I(scale(max_age_same_N, scale = FALSE)^2)  2.482e-05  2.473e-04   0.100
##                                               Pr(>|z|)    
## (Intercept)                                    < 2e-16 ***
## Sex                                            0.00183 ** 
## scale(max_age_same_N, scale = FALSE)           < 2e-16 ***
## I(scale(max_age_same_N, scale = FALSE)^2)      0.41698    
## Sex:scale(max_age_same_N, scale = FALSE)       0.47865    
## Sex:I(scale(max_age_same_N, scale = FALSE)^2)  0.92007    
## ---
## Signif. codes:  0 '***' 0.001 '**' 0.01 '*' 0.05 '.' 0.1 ' ' 1
## 
## (Dispersion parameter for Negative Binomial(1.2917) family taken to be 1)
## 
##     Null deviance: 15336  on 14469  degrees of freedom
## Residual deviance: 15200  on 14464  degrees of freedom
##   (1810 observations deleted due to missingness)
## AIC: 60904
## 
## Number of Fisher Scoring iterations: 1
## 
## 
##               Theta:  1.2917 
##           Std. Err.:  0.0234 
## 
##  2 x log-likelihood:  -60890.3580
```

```
1 - pchisq(summary(model_2)$deviance,
           summary(model_2)$df.residual)
```

```
## [1] 1.029767e-05
```

https://www.r-bloggers.com/2012/08/count-data-and-glms-choosing-among-poisson-negative-binomial-and-zero-inflated-models/ https://stats.stackexchange.com/questions/37732/when-someone-says-residual-deviance-df-should-1-for-a-poisson-model-how-appro
