## Supplementary material for "High Intelligence is not a Risk Factor for Mental Health Disorders": Supplemental_File_S5_Neuroticism_Age_Sex_new.html

Neuro\_Age\_Sex


### Neuro\_Age\_Sex

### Neuroticism - Age and Sex Effects

#### Neuroticism Score Distribution

##### Distribution of Raw Neuroticism Score

```
hist(Neuro_DF_no_NA$neuroticism_score)
```

##### Distribution of log transformed Neuroticism Score

```
hist(log(Neuro_DF_no_NA$neuroticism_score))
```

The linear model may not be adequate. Distribution ressembles poisson distribution.

#### Regressions

##### 1. Linear Model

```
model <- lm(neuroticism_score~ Sex*scale(Age_Center_0, scale = FALSE) + Sex*I(scale(Age_Center_0, scale = FALSE)^2), data = Neuro_DF_no_NA, na.action = na.exclude)
summary(model)
```

```
## 
## Call:
## lm(formula = neuroticism_score ~ Sex * scale(Age_Center_0, scale = FALSE) + 
##     Sex * I(scale(Age_Center_0, scale = FALSE)^2), data = Neuro_DF_no_NA, 
##     na.action = na.exclude)
## 
## Residuals:
##     Min      1Q  Median      3Q     Max 
## -4.9833 -2.7091 -0.4992  2.1770  9.0202 
## 
## Coefficients:
##                                               Estimate Std. Error t value
## (Intercept)                                  4.141e+00  7.415e-03 558.394
## Sex                                          9.742e-01  1.483e-02  65.686
## scale(Age_Center_0, scale = FALSE)          -4.160e-02  6.630e-04 -62.738
## I(scale(Age_Center_0, scale = FALSE)^2)     -8.774e-04  8.263e-05 -10.619
## Sex:scale(Age_Center_0, scale = FALSE)       1.933e-03  1.326e-03   1.457
## Sex:I(scale(Age_Center_0, scale = FALSE)^2) -5.619e-04  1.653e-04  -3.400
##                                             Pr(>|t|)    
## (Intercept)                                  < 2e-16 ***
## Sex                                          < 2e-16 ***
## scale(Age_Center_0, scale = FALSE)           < 2e-16 ***
## I(scale(Age_Center_0, scale = FALSE)^2)      < 2e-16 ***
## Sex:scale(Age_Center_0, scale = FALSE)      0.145003    
## Sex:I(scale(Age_Center_0, scale = FALSE)^2) 0.000674 ***
## ---
## Signif. codes:  0 '***' 0.001 '**' 0.01 '*' 0.05 '.' 0.1 ' ' 1
## 
## Residual standard error: 3.215 on 401290 degrees of freedom
## Multiple R-squared:  0.03125,    Adjusted R-squared:  0.03124 
## F-statistic:  2589 on 5 and 401290 DF,  p-value: < 2.2e-16
```

```
plot(model)
```

Residuals are not normally distributed. Let us try poisson regression.

##### 2. Poisson Model

Poisson regression is often used for modeling count data.

Assumption : conditional variance is equal to the conditional mean -> test overdispersion

```
model1 <- glm(neuroticism_score~ Sex*scale(Age_Center_0, scale = FALSE) + Sex*I(scale(Age_Center_0, scale = FALSE)^2), data = Neuro_DF_no_NA, family="poisson", na.action = na.exclude)
summary(model1)
```

```
## 
## Call:
## glm(formula = neuroticism_score ~ Sex * scale(Age_Center_0, scale = FALSE) + 
##     Sex * I(scale(Age_Center_0, scale = FALSE)^2), family = "poisson", 
##     data = Neuro_DF_no_NA, na.action = na.exclude)
## 
## Deviance Residuals: 
##     Min       1Q   Median       3Q      Max  
## -3.1527  -1.5160  -0.2541   0.9777   3.9144  
## 
## Coefficients:
##                                               Estimate Std. Error  z value
## (Intercept)                                  1.414e+00  1.147e-03 1232.214
## Sex                                          2.364e-01  2.295e-03  103.009
## scale(Age_Center_0, scale = FALSE)          -1.050e-02  1.051e-04  -99.917
## I(scale(Age_Center_0, scale = FALSE)^2)     -2.578e-04  1.286e-05  -20.049
## Sex:scale(Age_Center_0, scale = FALSE)       2.881e-03  2.102e-04   13.706
## Sex:I(scale(Age_Center_0, scale = FALSE)^2) -5.952e-05  2.571e-05   -2.315
##                                             Pr(>|z|)    
## (Intercept)                                   <2e-16 ***
## Sex                                           <2e-16 ***
## scale(Age_Center_0, scale = FALSE)            <2e-16 ***
## I(scale(Age_Center_0, scale = FALSE)^2)       <2e-16 ***
## Sex:scale(Age_Center_0, scale = FALSE)        <2e-16 ***
## Sex:I(scale(Age_Center_0, scale = FALSE)^2)   0.0206 *  
## ---
## Signif. codes:  0 '***' 0.001 '**' 0.01 '*' 0.05 '.' 0.1 ' ' 1
## 
## (Dispersion parameter for poisson family taken to be 1)
## 
##     Null deviance: 1188837  on 401295  degrees of freedom
## Residual deviance: 1155858  on 401290  degrees of freedom
## AIC: 2258320
## 
## Number of Fisher Scoring iterations: 5
```

Our residual deviance is 1155858 for 401290 degrees of freedom. The rule of thumb is ratio = 1, here : 1155858/401290 = 2.9 - So we have moderate dispersion, which we can also test with a dispersion test

```
library("AER")
```

```
## Loading required package: car
```

```
## Loading required package: carData
```

```
## Registered S3 methods overwritten by 'car':
##   method                          from
##   influence.merMod                lme4
##   cooks.distance.influence.merMod lme4
##   dfbeta.influence.merMod         lme4
##   dfbetas.influence.merMod        lme4
```

```
## 
## Attaching package: 'car'
```

```
## The following object is masked from 'package:dplyr':
## 
##     recode
```

```
## Loading required package: lmtest
```

```
## Loading required package: zoo
```

```
## 
## Attaching package: 'zoo'
```

```
## The following objects are masked from 'package:base':
## 
##     as.Date, as.Date.numeric
```

```
## Loading required package: survival
```

```
dispersiontest(model1)
```

```
## 
##  Overdispersion test
## 
## data:  model1
## z = 341.36, p-value < 2.2e-16
## alternative hypothesis: true dispersion is greater than 1
## sample estimates:
## dispersion 
##   2.542724
```

```
1 - pchisq(summary(model1)$deviance,
           summary(model1)$df.residual)
```

```
## [1] 0
```

The null hypothesis is that our model is correctly specified, and we have strong evidence to reject that hypothesis. So we have strong evidence that our model fits badly. Here we simulated the data, and we in fact know that the model we have fitted is the correct model. So here the deviance goodness of fit test has wrongly indicated that our model is incorrectly specified. But perhaps we were just unlucky – by chance 5% of the time the test will reject even when the null hypothesis is true.

##### 3. “Fixing” overdispersion

### a. by using quasi-families regression

quasi-families augment the normal families by adding a dispersion parameter http://biometry.github.io/APES//LectureNotes/2016-JAGS/Overdispersion/OverdispersionJAGS.html

```
model <- glm(neuroticism_score~ Sex*Age_Center_0 + Sex*I(Age_Center_0^2), data = Neuro_DF_no_NA, family="quasipoisson", na.action = na.exclude)
summary(model)
```

```
## 
## Call:
## glm(formula = neuroticism_score ~ Sex * Age_Center_0 + Sex * 
##     I(Age_Center_0^2), family = "quasipoisson", data = Neuro_DF_no_NA, 
##     na.action = na.exclude)
## 
## Deviance Residuals: 
##     Min       1Q   Median       3Q      Max  
## -3.1527  -1.5160  -0.2541   0.9777   3.9144  
## 
## Coefficients:
##                         Estimate Std. Error t value Pr(>|t|)    
## (Intercept)            1.177e+00  6.220e-02  18.914   <2e-16 ***
## Sex                   -1.204e-01  1.244e-01  -0.968    0.333    
## Age_Center_0           1.884e-02  2.277e-03   8.273   <2e-16 ***
## I(Age_Center_0^2)     -2.578e-04  2.050e-05 -12.573   <2e-16 ***
## Sex:Age_Center_0       9.656e-03  4.554e-03   2.120    0.034 *  
## Sex:I(Age_Center_0^2) -5.952e-05  4.100e-05  -1.452    0.147    
## ---
## Signif. codes:  0 '***' 0.001 '**' 0.01 '*' 0.05 '.' 0.1 ' ' 1
## 
## (Dispersion parameter for quasipoisson family taken to be 2.542767)
## 
##     Null deviance: 1188837  on 401295  degrees of freedom
## Residual deviance: 1155858  on 401290  degrees of freedom
## AIC: NA
## 
## Number of Fisher Scoring iterations: 5
```

```
1 - pchisq(summary(model)$deviance,
           summary(model)$df.residual)
```

```
## [1] 0
```

The GOF test indicates that the model fits the data if p > 0.05. The null hypothesis is that our model is correctly specified, and we have strong evidence to reject that hypothesis. So we have strong evidence that our model fits badly. Here we simulated the data, and we in fact know that the model we have fitted is the correct model. So here the deviance goodness of fit test has wrongly indicated that our model is incorrectly specified. But perhaps we were just unlucky – by chance 5% of the time the test will reject even when the null hypothesis is true.

You see that τ (dispersion parameter) is estimated at a value similar to those in the overdispersion tests above (as you’d expect). The main effect is the substantially larger errors for the estimates (the point estimates do not change), and hence potentially changed significances (though not here). (You can manually compute the corrected standard errors as Poisson-standard errors ·√τ .) Note that because this is no maximum likelihood method (but a quasi-likelihood method), no likelihood and hence no AIC are available. No overdispersion tests can be conducted for quasi-family objects (neither in AER nor DHARMa)

### b. by using negative binomial regression

Conditional variance exceedes the conditional mean -> test overdispersion Maybe our distributional assumption was simply wrong, and we choose a different distribution

https://biometry.github.io/APES/LectureNotes/2016-JAGS/Overdispersion/OverdispersionJAGS.pdf https://stats.idre.ucla.edu/r/dae/negative-binomial-regression/

```
library(MASS)
```

```
## 
## Attaching package: 'MASS'
```

```
## The following object is masked from 'package:dplyr':
## 
##     select
```

```
model_2 <- glm.nb(neuroticism_score~ Sex*scale(Age_Center_0, scale = FALSE)+ Sex*I(scale(Age_Center_0, scale = FALSE)^2), data = Neuro_DF_no_NA, na.action = na.exclude)
summary(model_2)
```

```
## 
## Call:
## glm.nb(formula = neuroticism_score ~ Sex * scale(Age_Center_0, 
##     scale = FALSE) + Sex * I(scale(Age_Center_0, scale = FALSE)^2), 
##     data = Neuro_DF_no_NA, na.action = na.exclude, init.theta = 1.965515583, 
##     link = log)
## 
## Deviance Residuals: 
##     Min       1Q   Median       3Q      Max  
## -2.2264  -1.0219  -0.1477   0.5342   2.0996  
## 
## Coefficients:
##                                               Estimate Std. Error z value
## (Intercept)                                  1.414e+00  2.006e-03 704.624
## Sex                                          2.363e-01  4.013e-03  58.890
## scale(Age_Center_0, scale = FALSE)          -1.050e-02  1.808e-04 -58.065
## I(scale(Age_Center_0, scale = FALSE)^2)     -2.569e-04  2.240e-05 -11.467
## Sex:scale(Age_Center_0, scale = FALSE)       2.885e-03  3.616e-04   7.977
## Sex:I(scale(Age_Center_0, scale = FALSE)^2) -5.845e-05  4.481e-05  -1.304
##                                             Pr(>|z|)    
## (Intercept)                                  < 2e-16 ***
## Sex                                          < 2e-16 ***
## scale(Age_Center_0, scale = FALSE)           < 2e-16 ***
## I(scale(Age_Center_0, scale = FALSE)^2)      < 2e-16 ***
## Sex:scale(Age_Center_0, scale = FALSE)       1.5e-15 ***
## Sex:I(scale(Age_Center_0, scale = FALSE)^2)    0.192    
## ---
## Signif. codes:  0 '***' 0.001 '**' 0.01 '*' 0.05 '.' 0.1 ' ' 1
## 
## (Dispersion parameter for Negative Binomial(1.9655) family taken to be 1)
## 
##     Null deviance: 486784  on 401295  degrees of freedom
## Residual deviance: 475993  on 401290  degrees of freedom
## AIC: 1986416
## 
## Number of Fisher Scoring iterations: 1
## 
## 
##               Theta:  1.96552 
##           Std. Err.:  0.00737 
## 
##  2 x log-likelihood:  -1986402.27400
```

```
1 - pchisq(summary(model_2)$deviance,
           summary(model_2)$df.residual)
```

```
## [1] 0
```

https://www.r-bloggers.com/2012/08/count-data-and-glms-choosing-among-poisson-negative-binomial-and-zero-inflated-models/ https://stats.stackexchange.com/questions/37732/when-someone-says-residual-deviance-df-should-1-for-a-poisson-model-how-appro

The ratio of deviance and df is near 1 and hence probably fine. We can also check with a dispersion test. BUT The GOF test indicates that the model does not fit the data (p > 0.05).

```
library(DHARMa)
```

```
## This is DHARMa 0.4.1. For overview type '?DHARMa'. For recent changes, type news(package = 'DHARMa') Note: Syntax of plotResiduals has changed in 0.3.0, see ?plotResiduals for details
```

```
simulationOutput <- simulateResiduals(model_2)
testDispersion(simulationOutput)
```

```
## 
##  DHARMa nonparametric dispersion test via sd of residuals fitted vs.
##  simulated
## 
## data:  simulationOutput
## dispersion = 0.80013, p-value < 2.2e-16
## alternative hypothesis: two.sided
```

```
testZeroInflation(simulationOutput)
```

```
## 
##  DHARMa zero-inflation test via comparison to expected zeros with
##  simulation under H0 = fitted model
## 
## data:  simulationOutput
## ratioObsSim = 1.3792, p-value < 2.2e-16
## alternative hypothesis: two.sided
```

https://stats.stackexchange.com/questions/490680/significant-dispersion-test

A word of warning that applies also to all other tests that follow: significance in hypothesis tests depends on at least 2 ingredients: strength of the signal, and number of data points. Hence, the p-value alone is not a good indicator of the extent to which your residuals deviate from assumptions. Specifically, if you have a lot of data points, residual diagnostics will nearly inevitably become significant, because having a perfectly fitting model is very unlikely. That, however, doesn’t necessarily mean that you need to change your model. The p-values confirm that there is a deviation from your null hypothesis. It is, however, in your discretion to decide whether this deviation is worth worrying about. If you see a dispersion parameter of 1.01, I would not worry, even if the test is significant. A significant value of 5, however, is clearly a reason to move to a model that accounts for overdispersion.

### c. Let us use a likelihood ratio test to compare these two and test this model assumption

“As we mentioned earlier, negative binomial models assume the conditional means are not equal to the conditional variances. This inequality is captured by estimating a dispersion parameter (not shown in the output) that is held constant in a Poisson model. Thus, the Poisson model is actually nested in the negative binomial model. We can then use a likelihood ratio test to compare these two and test this model assumption.”

https://stats.idre.ucla.edu/r/dae/negative-binomial-regression/

```
pchisq(2 * (logLik(model_2) - logLik(model1)), df = 1, lower.tail = FALSE)
```

```
## 'log Lik.' 0 (df=7)
```

```
library("lmtest")
lrtest(model1, model_2)
```

```
## Likelihood ratio test
## 
## Model 1: neuroticism_score ~ Sex * scale(Age_Center_0, scale = FALSE) + 
##     Sex * I(scale(Age_Center_0, scale = FALSE)^2)
## Model 2: neuroticism_score ~ Sex * scale(Age_Center_0, scale = FALSE) + 
##     Sex * I(scale(Age_Center_0, scale = FALSE)^2)
##   #Df   LogLik Df  Chisq Pr(>Chisq)    
## 1   6 -1129154                         
## 2   7  -993201  1 271906  < 2.2e-16 ***
## ---
## Signif. codes:  0 '***' 0.001 '**' 0.01 '*' 0.05 '.' 0.1 ' ' 1
```

In this example the associated chi-squared value estimated from 2\*(logLik(m1) – logLik(m3)) is 926.03 with one degree of freedom. This strongly suggests the negative binomial model, estimating the dispersion parameter, is more appropriate than the Poisson model.

### Save Negative Binomial Output

```
Summary_age_sex_model_binary_Depression_Ever <- as.data.frame(summary(model_2)$coefficients)
Summary_age_sex_model_binary_Depression_Ever$Model <- "Neuroticism_Score"
names(Summary_age_sex_model_binary_Depression_Ever) <- c("Estimate", "SE", "t/z", "p", "Model")
fwrite( Summary_age_sex_model_binary_Depression_Ever, "/scratch2/ukbio/biobank/High_IQ_Project/results/Sex_Age_Effects_Neuroticism.csv")
```
