## Supplementary material for "High Intelligence is not a Risk Factor for Mental Health Disorders": Supplemental_File_S7_AUDIT_Threshold.html

Alcohol\_AUDIT\_Threshold


### Alcohol\_AUDIT\_Threshold

### I- ICD10

```
## `summarise()` has grouped output by 'AUDIT.Score.Final'. You can override using the `.groups` argument.
```

```
## # A tibble: 79 × 3
## # Groups:   AUDIT.Score.Final [41]
##    AUDIT.Score.Final sex        mean
##                <int> <chr>     <dbl>
##  1                 0 Female 0.00112 
##  2                 0 Male   0.00297 
##  3                 1 Female 0.000579
##  4                 1 Male   0.00163 
##  5                 2 Female 0.00230 
##  6                 2 Male   0.00803 
##  7                 3 Female 0.00168 
##  8                 3 Male   0.00311 
##  9                 4 Female 0.00420 
## 10                 4 Male   0.0119  
## # … with 69 more rows
```

```
## `summarise()` has grouped output by 'AUDIT.Score.Final'. You can override using the `.groups` argument.
```

```
## # A tibble: 79 × 3
## # Groups:   AUDIT.Score.Final [41]
##    AUDIT.Score.Final sex        mean
##                <int> <chr>     <dbl>
##  1                 0 Female 0.000112
##  2                 0 Male   0.00135 
##  3                 1 Female 0.000166
##  4                 1 Male   0.000466
##  5                 2 Female 0.00123 
##  6                 2 Male   0.00485 
##  7                 3 Female 0.000219
##  8                 3 Male   0.000812
##  9                 4 Female 0.00125 
## 10                 4 Male   0.00462 
## # … with 69 more rows
```

```
## `summarise()` has grouped output by 'AUDIT.Score.Final'. You can override using the `.groups` argument.
```

```
## # A tibble: 79 × 3
## # Groups:   AUDIT.Score.Final [41]
##    AUDIT.Score.Final sex        mean
##                <int> <chr>     <dbl>
##  1                 0 Female 0.000781
##  2                 0 Male   0.00243 
##  3                 1 Female 0.000166
##  4                 1 Male   0.000466
##  5                 2 Female 0.000741
##  6                 2 Male   0.00284 
##  7                 3 Female 0.000876
##  8                 3 Male   0.000812
##  9                 4 Female 0.00192 
## 10                 4 Male   0.00586 
## # … with 69 more rows
```

```
##    AUDIT.Score.Final.8 Alcohol_ICD10      n
## 1:                   0             0 124094
## 2:                   0             1    566
## 3:                   1             0  31681
## 4:                   1             1    901
```

```
##    AUDIT.Score.Final.8 Alcohol_ICD10    sex     n
## 1:                   0             0 Female 76665
## 2:                   0             0   Male 47429
## 3:                   0             1 Female   205
## 4:                   0             1   Male   361
## 5:                   1             0 Female 11944
## 6:                   1             0   Male 19737
## 7:                   1             1 Female   224
## 8:                   1             1   Male   677
```

```
##    AUDIT.Score.Final.12 Alcohol_ICD10      n
## 1:                    0             0 144540
## 2:                    0             1    886
## 3:                    1             0  11235
## 4:                    1             1    581
```

```
##    AUDIT.Score.Final.12 Alcohol_ICD10    sex     n
## 1:                    0             0 Female 84542
## 2:                    0             0   Male 59998
## 3:                    0             1 Female   277
## 4:                    0             1   Male   609
## 5:                    1             0 Female  4067
## 6:                    1             0   Male  7168
## 7:                    1             1 Female   152
## 8:                    1             1   Male   429
```

```
##    AUDIT.Score.Final.15 Alcohol_ICD10      n
## 1:                    0             0 150354
## 2:                    0             1   1068
## 3:                    1             0   5421
## 4:                    1             1    399
```

```
##    AUDIT.Score.Final.15 Alcohol_ICD10    sex     n
## 1:                    0             0 Female 86581
## 2:                    0             0   Male 63773
## 3:                    0             1 Female   314
## 4:                    0             1   Male   754
## 5:                    1             0 Female  2028
## 6:                    1             0   Male  3393
## 7:                    1             1 Female   115
## 8:                    1             1   Male   284
```

```
##    AUDIT.Score.Final.20 Alcohol_ICD10      n
## 1:                    0             0 154269
## 2:                    0             1   1285
## 3:                    1             0   1506
## 4:                    1             1    182
```

```
##    AUDIT.Score.Final.20 Alcohol_ICD10    sex     n
## 1:                    0             0 Female 87960
## 2:                    0             0   Male 66309
## 3:                    0             1 Female   374
## 4:                    0             1   Male   911
## 5:                    1             0 Female   649
## 6:                    1             0   Male   857
## 7:                    1             1 Female    55
## 8:                    1             1   Male   127
```

### II - AUC curve

#### 1. across sexes

The AUC shows whether the prediction of icd10 diagnosis based on each audit score identifies more true positive results (e.g., the proportion of people with icd10 diagnoses who have an audit score; on the y axis in Figure 2) than false positive results (e.g., the proportion of people without ICD10 diagnosis who have an audit score; on the x axis in Figure 2).

AUC (up to AUC = 1 showing perfect accuracy).

We found that the AUC for having a mental health problem was 0.776 (Figure 2). This AUC represents a 77% probability (i.e., only 27% above chance) that a person from the UKB with an ICD10 diagnosis had a higher audit score than a UKB participant without an ICD10 diagnosis. In other words, the audit score accurately distinguish a UK Biobank person who had an icd10 diagnosis from a a UKB person who did not.

```
## Type 'citation("pROC")' for a citation.
```

```
## 
## Attaching package: 'pROC'
```

```
## The following objects are masked from 'package:stats':
## 
##     cov, smooth, var
```

```
## randomForest 4.6-14
```

```
## Type rfNews() to see new features/changes/bug fixes.
```

```
## 
## Attaching package: 'randomForest'
```

```
## The following object is masked from 'package:dplyr':
## 
##     combine
```

```
## The following object is masked from 'package:ggplot2':
## 
##     margin
```

```
## 
## Call:
## glm(formula = Alcohol_ICD10 ~ AUDIT.Score.Final, family = binomial, 
##     data = A)
## 
## Deviance Residuals: 
##     Min       1Q   Median       3Q      Max  
## -1.5260  -0.1342  -0.1038  -0.0875   3.4380  
## 
## Coefficients:
##                    Estimate Std. Error z value Pr(>|z|)    
## (Intercept)       -5.907211   0.045516  -129.8   <2e-16 ***
## AUDIT.Score.Final  0.171729   0.003571    48.1   <2e-16 ***
## ---
## Signif. codes:  0 '***' 0.001 '**' 0.01 '*' 0.05 '.' 0.1 ' ' 1
## 
## (Dispersion parameter for binomial family taken to be 1)
## 
##     Null deviance: 16635  on 157241  degrees of freedom
## Residual deviance: 14847  on 157240  degrees of freedom
## AIC: 14851
## 
## Number of Fisher Scoring iterations: 8
```

```
## Setting levels: control = 0, case = 1
```

```
## Setting direction: controls < cases
```

```
## 
## Call:
## roc.default(response = A$Alcohol_ICD10, predictor = A$AUDIT.Score.Final,     plot = T)
## 
## Data: A$AUDIT.Score.Final in 155775 controls (A$Alcohol_ICD10 0) < 1467 cases (A$Alcohol_ICD10 1).
## Area under the curve: 0.7761
```

```
## Setting levels: control = 0, case = 1
## Setting direction: controls < cases
```

```
## 
## Call:
## roc.default(response = A$Alcohol_ICD10, predictor = A$AUDIT.Score.Final,     percent = T, plot = T, xlab = "False Positive Percentage",     ylab = "True Positive Percentage", legacy.axes = T, print.auc = TRUE)
## 
## Data: A$AUDIT.Score.Final in 155775 controls (A$Alcohol_ICD10 0) < 1467 cases (A$Alcohol_ICD10 1).
## Area under the curve: 77.61%
```

```
## Setting levels: control = 0, case = 1
## Setting direction: controls < cases
```

```
##         tpp      fpp sensitivities specificities thresholds
## 5  89.29789 56.27989     0.8929789     0.4372011        3.5
## 6  78.18678 41.97785     0.7818678     0.5802215        4.5
## 7  72.86980 32.59958     0.7286980     0.6740042        5.5
## 8  66.73483 25.50538     0.6673483     0.7449462        6.5
## 9  61.41786 20.33767     0.6141786     0.7966233        7.5
## 10 55.07839 15.73808     0.5507839     0.8426192        8.5
```

```
##             tpp          fpp sensitivities specificities thresholds
## 1  100.00000000 1.000000e+02  1.0000000000    0.00000000       -Inf
## 2   98.56850716 9.187996e+01  0.9856850716    0.08120045        0.5
## 3   97.61417860 8.137827e+01  0.9761417860    0.18621730        1.5
## 4   92.43353783 6.978912e+01  0.9243353783    0.30210881        2.5
## 5   89.29788684 5.627989e+01  0.8929788684    0.43720109        3.5
## 6   78.18677573 4.197785e+01  0.7818677573    0.58022147        4.5
## 7   72.86980232 3.259958e+01  0.7286980232    0.67400417        5.5
## 8   66.73483299 2.550538e+01  0.6673483299    0.74494624        6.5
## 9   61.41785958 2.033767e+01  0.6141785958    0.79662333        7.5
## 10  55.07839127 1.573808e+01  0.5507839127    0.84261916        8.5
## 11  49.69325153 1.204494e+01  0.4969325153    0.87955063        9.5
## 12  44.44444444 9.281335e+00  0.4444444444    0.90718665       10.5
## 13  39.60463531 7.212325e+00  0.3960463531    0.92787675       11.5
## 14  35.24199046 5.625421e+00  0.3524199046    0.94374579       12.5
## 15  31.28834356 4.436527e+00  0.3128834356    0.95563473       13.5
## 16  27.19836401 3.480019e+00  0.2719836401    0.96519981       14.5
## 17  23.51738241 2.666667e+00  0.2351738241    0.97333333       15.5
## 18  19.29107021 2.069010e+00  0.1929107021    0.97930990       16.5
## 19  16.08725290 1.604879e+00  0.1608725290    0.98395121       17.5
## 20  13.56509884 1.240250e+00  0.1356509884    0.98759750       18.5
## 21  12.40627130 9.667790e-01  0.1240627130    0.99033221       19.5
## 22  11.04294479 7.356765e-01  0.1104294479    0.99264324       20.5
## 23   9.95228357 5.662013e-01  0.0995228357    0.99433799       21.5
## 24   8.72528971 4.236880e-01  0.0872528971    0.99576312       22.5
## 25   7.36196319 3.113465e-01  0.0736196319    0.99688654       23.5
## 26   5.72597137 2.291767e-01  0.0572597137    0.99770823       24.5
## 27   4.83980913 1.656235e-01  0.0483980913    0.99834377       25.5
## 28   3.88548057 1.219708e-01  0.0388548057    0.99878029       26.5
## 29   3.13565099 8.152784e-02  0.0313565099    0.99918472       27.5
## 30   2.65848671 6.034344e-02  0.0265848671    0.99939657       28.5
## 31   2.18132243 4.301075e-02  0.0218132243    0.99956989       29.5
## 32   1.97682345 2.567806e-02  0.0197682345    0.99974322       30.5
## 33   1.29516019 1.797464e-02  0.0129516019    0.99982025       31.5
## 34   1.09066121 1.091318e-02  0.0109066121    0.99989087       32.5
## 35   0.88616224 7.061467e-03  0.0088616224    0.99992939       33.5
## 36   0.74982958 5.135612e-03  0.0074982958    0.99994864       34.5
## 37   0.54533061 4.493661e-03  0.0054533061    0.99995506       35.5
## 38   0.27266530 1.925855e-03  0.0027266530    0.99998074       36.5
## 39   0.20449898 1.925855e-03  0.0020449898    0.99998074       37.5
## 40   0.06816633 6.419515e-04  0.0006816633    0.99999358       38.5
## 41   0.06816633 0.000000e+00  0.0006816633    1.00000000       39.5
## 42   0.00000000 0.000000e+00  0.0000000000    1.00000000        Inf
```

#### 2. Males

```
## 
## Call:
## glm(formula = Alcohol_ICD10 ~ AUDIT.Score.Final, family = binomial, 
##     data = A1)
## 
## Deviance Residuals: 
##     Min       1Q   Median       3Q      Max  
## -1.4873  -0.1733  -0.1369  -0.1170   3.3043  
## 
## Coefficients:
##                    Estimate Std. Error z value Pr(>|z|)    
## (Intercept)       -5.454770   0.058760  -92.83   <2e-16 ***
## AUDIT.Score.Final  0.157925   0.004615   34.22   <2e-16 ***
## ---
## Signif. codes:  0 '***' 0.001 '**' 0.01 '*' 0.05 '.' 0.1 ' ' 1
## 
## (Dispersion parameter for binomial family taken to be 1)
## 
##     Null deviance: 10748.6  on 68203  degrees of freedom
## Residual deviance:  9780.2  on 68202  degrees of freedom
## AIC: 9784.2
## 
## Number of Fisher Scoring iterations: 7
```

```
## Setting levels: control = 0, case = 1
```

```
## Setting direction: controls < cases
```

```
## 
## Call:
## roc.default(response = A1$Alcohol_ICD10, predictor = A1$AUDIT.Score.Final,     plot = T)
## 
## Data: A1$AUDIT.Score.Final in 67166 controls (A1$Alcohol_ICD10 0) < 1038 cases (A1$Alcohol_ICD10 1).
## Area under the curve: 0.7389
```

```
## Setting levels: control = 0, case = 1
## Setting direction: controls < cases
```

```
## 
## Call:
## roc.default(response = A1$Alcohol_ICD10, predictor = A1$AUDIT.Score.Final,     percent = T, plot = T, xlab = "False Positive Percentage",     ylab = "True Positive Percentage", legacy.axes = T, print.auc = TRUE)
## 
## Data: A1$AUDIT.Score.Final in 67166 controls (A1$Alcohol_ICD10 0) < 1038 cases (A1$Alcohol_ICD10 1).
## Area under the curve: 73.89%
```

```
## Setting levels: control = 0, case = 1
## Setting direction: controls < cases
```

```
##         tpp      fpp sensitivities specificities thresholds
## 6  81.21387 55.27201     0.8121387     0.4472799        4.5
## 7  76.10790 44.71608     0.7610790     0.5528392        5.5
## 8  70.71291 35.98696     0.7071291     0.6401304        6.5
## 9  65.22158 29.38540     0.6522158     0.7061460        7.5
## 10 58.76686 23.14266     0.5876686     0.7685734        8.5
## 11 52.40848 17.81109     0.5240848     0.8218891        9.5
```

```
##             tpp          fpp sensitivities specificities thresholds
## 1  100.00000000 1.000000e+02  1.0000000000    0.00000000       -Inf
## 2   98.94026975 9.450168e+01  0.9894026975    0.05498318        0.5
## 3   98.26589595 8.812197e+01  0.9826589595    0.11878034        1.5
## 4   93.64161850 7.929161e+01  0.9364161850    0.20708394        2.5
## 5   91.42581888 6.833070e+01  0.9142581888    0.31669297        3.5
## 6   81.21387283 5.527201e+01  0.8121387283    0.44727987        4.5
## 7   76.10789981 4.471608e+01  0.7610789981    0.55283923        5.5
## 8   70.71290944 3.598696e+01  0.7071290944    0.64013042        6.5
## 9   65.22157996 2.938540e+01  0.6522157996    0.70614597        7.5
## 10  58.76685934 2.314266e+01  0.5876685934    0.76857339        8.5
## 11  52.40847784 1.781109e+01  0.5240847784    0.82188905        9.5
## 12  46.72447013 1.369890e+01  0.4672447013    0.86301105       10.5
## 13  41.32947977 1.067207e+01  0.4132947977    0.89327934       11.5
## 14  36.12716763 8.289909e+00  0.3612716763    0.91710091       12.5
## 15  32.17726397 6.521157e+00  0.3217726397    0.93478843       13.5
## 16  27.36030829 5.051663e+00  0.2736030829    0.94948337       14.5
## 17  23.89210019 3.823363e+00  0.2389210019    0.96176637       15.5
## 18  19.46050096 2.897299e+00  0.1946050096    0.97102701       16.5
## 19  15.99229287 2.204985e+00  0.1599229287    0.97795015       17.5
## 20  13.19845857 1.666021e+00  0.1319845857    0.98333979       18.5
## 21  12.23506744 1.275943e+00  0.1223506744    0.98724057       19.5
## 22  10.88631985 9.379746e-01  0.1088631985    0.99062025       20.5
## 23  10.21194605 7.086919e-01  0.1021194605    0.99291308       21.5
## 24   8.86319846 5.225858e-01  0.0886319846    0.99477414       22.5
## 25   7.51445087 3.662567e-01  0.0751445087    0.99633743       23.5
## 26   5.97302505 2.575708e-01  0.0597302505    0.99742429       24.5
## 27   4.91329480 1.786618e-01  0.0491329480    0.99821338       25.5
## 28   3.94990366 1.265521e-01  0.0394990366    0.99873448       26.5
## 29   3.08285164 9.677515e-02  0.0308285164    0.99903225       27.5
## 30   2.69749518 6.848703e-02  0.0269749518    0.99931513       28.5
## 31   2.02312139 5.210970e-02  0.0202312139    0.99947890       29.5
## 32   1.83044316 3.424352e-02  0.0183044316    0.99965756       30.5
## 33   1.15606936 2.084388e-02  0.0115606936    0.99979156       31.5
## 34   0.96339114 1.191079e-02  0.0096339114    0.99988089       32.5
## 35   0.86705202 7.444243e-03  0.0086705202    0.99992556       33.5
## 36   0.67437380 5.955394e-03  0.0067437380    0.99994045       34.5
## 37   0.48169557 5.955394e-03  0.0048169557    0.99994045       35.5
## 38   0.19267823 2.977697e-03  0.0019267823    0.99997022       37.0
## 39   0.09633911 1.488849e-03  0.0009633911    0.99998511       38.5
## 40   0.09633911 0.000000e+00  0.0009633911    1.00000000       39.5
## 41   0.00000000 0.000000e+00  0.0000000000    1.00000000        Inf
```

#### 3. Females

```
## 
## Call:
## glm(formula = Alcohol_ICD10 ~ AUDIT.Score.Final, family = binomial, 
##     data = A1)
## 
## Deviance Residuals: 
##     Min       1Q   Median       3Q      Max  
## -1.2889  -0.0878  -0.0737  -0.0674   3.5885  
## 
## Coefficients:
##                    Estimate Std. Error z value Pr(>|z|)    
## (Intercept)       -6.437194   0.076652  -83.98   <2e-16 ***
## AUDIT.Score.Final  0.176201   0.006095   28.91   <2e-16 ***
## ---
## Signif. codes:  0 '***' 0.001 '**' 0.01 '*' 0.05 '.' 0.1 ' ' 1
## 
## (Dispersion parameter for binomial family taken to be 1)
## 
##     Null deviance: 5433.7  on 89037  degrees of freedom
## Residual deviance: 4860.1  on 89036  degrees of freedom
## AIC: 4864.1
## 
## Number of Fisher Scoring iterations: 8
```

```
## Setting levels: control = 0, case = 1
```

```
## Setting direction: controls < cases
```

```
## 
## Call:
## roc.default(response = A1$Alcohol_ICD10, predictor = A1$AUDIT.Score.Final,     plot = T)
## 
## Data: A1$AUDIT.Score.Final in 88609 controls (A1$Alcohol_ICD10 0) < 429 cases (A1$Alcohol_ICD10 1).
## Area under the curve: 0.7773
```

```
## Setting levels: control = 0, case = 1
## Setting direction: controls < cases
```

```
## 
## Call:
## roc.default(response = A1$Alcohol_ICD10, predictor = A1$AUDIT.Score.Final,     percent = T, plot = T, xlab = "False Positive Percentage",     ylab = "True Positive Percentage", legacy.axes = T, print.auc = TRUE)
## 
## Data: A1$AUDIT.Score.Final in 88609 controls (A1$Alcohol_ICD10 0) < 429 cases (A1$Alcohol_ICD10 1).
## Area under the curve: 77.73%
```

```
## Setting levels: control = 0, case = 1
## Setting direction: controls < cases
```

```
##        tpp      fpp sensitivities specificities thresholds
## 4 89.51049 62.58619     0.8951049     0.3741381        2.5
## 5 84.14918 47.14532     0.8414918     0.5285468        3.5
## 6 70.86247 31.90082     0.7086247     0.6809918        4.5
## 7 65.03497 23.41523     0.6503497     0.7658477        5.5
## 8 57.10956 17.56029     0.5710956     0.8243971        6.5
## 9 52.21445 13.47944     0.5221445     0.8652056        7.5
```

```
##            tpp          fpp sensitivities specificities thresholds
## 1  100.0000000 1.000000e+02   1.000000000     0.0000000       -Inf
## 2   97.6689977 8.989267e+01   0.976689977     0.1010733        0.5
## 3   96.0372960 7.626652e+01   0.960372960     0.2373348        1.5
## 4   89.5104895 6.258619e+01   0.895104895     0.3741381        2.5
## 5   84.1491841 4.714532e+01   0.841491841     0.5285468        3.5
## 6   70.8624709 3.190082e+01   0.708624709     0.6809918        4.5
## 7   65.0349650 2.341523e+01   0.650349650     0.7658477        5.5
## 8   57.1095571 1.756029e+01   0.571095571     0.8243971        6.5
## 9   52.2144522 1.347944e+01   0.522144522     0.8652056        7.5
## 10  46.1538462 1.012538e+01   0.461538462     0.8987462        8.5
## 11  43.1235431 7.674164e+00   0.431235431     0.9232584        9.5
## 12  38.9277389 5.932806e+00   0.389277389     0.9406719       10.5
## 13  35.4312354 4.589827e+00   0.354312354     0.9541017       11.5
## 14  33.1002331 3.605729e+00   0.331002331     0.9639427       12.5
## 15  29.1375291 2.856369e+00   0.291375291     0.9714363       13.5
## 16  26.8065268 2.288707e+00   0.268065268     0.9771129       14.5
## 17  22.6107226 1.789886e+00   0.226107226     0.9821011       15.5
## 18  18.8811189 1.441163e+00   0.188811189     0.9855884       16.5
## 19  16.3170163 1.149996e+00   0.163170163     0.9885000       17.5
## 20  14.4522145 9.175140e-01   0.144522145     0.9908249       18.5
## 21  12.8205128 7.324312e-01   0.128205128     0.9926757       19.5
## 22  11.4219114 5.823336e-01   0.114219114     0.9941767       20.5
## 23   9.3240093 4.581927e-01   0.093240093     0.9954181       21.5
## 24   8.3916084 3.487230e-01   0.083916084     0.9965128       22.5
## 25   6.9930070 2.697243e-01   0.069930070     0.9973028       23.5
## 26   5.1282051 2.076539e-01   0.051282051     0.9979235       24.5
## 27   4.6620047 1.557404e-01   0.046620047     0.9984426       25.5
## 28   3.7296037 1.184981e-01   0.037296037     0.9988150       26.5
## 29   3.2634033 6.997032e-02   0.032634033     0.9993003       27.5
## 30   2.5641026 5.417057e-02   0.025641026     0.9994583       28.5
## 31   2.5641026 3.611371e-02   0.025641026     0.9996389       29.5
## 32   2.3310023 1.918541e-02   0.023310023     0.9998081       30.5
## 33   1.6317016 1.579975e-02   0.016317016     0.9998420       31.5
## 34   1.3986014 1.015698e-02   0.013986014     0.9998984       32.5
## 35   0.9324009 6.771321e-03   0.009324009     0.9999323       33.5
## 36   0.9324009 4.514214e-03   0.009324009     0.9999549       34.5
## 37   0.6993007 3.385661e-03   0.006993007     0.9999661       35.5
## 38   0.4662005 1.128554e-03   0.004662005     0.9999887       36.5
## 39   0.2331002 1.128554e-03   0.002331002     0.9999887       37.5
## 40   0.0000000 0.000000e+00   0.000000000     1.0000000        Inf
```
