## Supplementary material for "High Intelligence is not a Risk Factor for Mental Health Disorders": Supplemental_Files_S8_Interactions_WellBeing.html

Cognitive Ability (CA) Group Significant Interactions with Age


### Cognitive Ability (CA) Group Significant Interactions with Age

#### I- Well-being - CA Group by Age (Average vs. Low)

```
## [1] "Average_CA" "High_CA"    "Low_CA"
```

```
##                                             Estimate          SE         t/z
## (Intercept)                              0.016404613 0.004296955   3.8177297
## CA_GroupHigh_CA                          0.013379548 0.013899663   0.9625808
## CA_GroupLow_CA                          -0.119798710 0.026736800  -4.4806676
## scale(max_age_MHQ)                       0.071659724 0.003190524  22.4601730
## Sex                                      0.018447293 0.006156951   2.9961736
## I(scale(max_age_MHQ)^2)                 -0.019158947 0.002997828  -6.3909425
## CA_GroupHigh_CA:scale(max_age_MHQ)       0.002261923 0.010676606   0.2118579
## CA_GroupLow_CA:scale(max_age_MHQ)        0.069955643 0.019046507   3.6728857
## CA_GroupHigh_CA:Sex                     -0.047734029 0.020276751  -2.3541261
## CA_GroupLow_CA:Sex                       0.010575502 0.037987288   0.2783958
## scale(max_age_MHQ):Sex                  -0.064411243 0.006151204 -10.4713228
## CA_GroupHigh_CA:I(scale(max_age_MHQ)^2)  0.008221997 0.009647441   0.8522464
## CA_GroupLow_CA:I(scale(max_age_MHQ)^2)  -0.010879404 0.018271177  -0.5954408
## CA_GroupHigh_CA:scale(max_age_MHQ):Sex  -0.008761748 0.020227681  -0.4331563
## CA_GroupLow_CA:scale(max_age_MHQ):Sex    0.019973116 0.037278297   0.5357840
##                                                     p      Model
## (Intercept)                              1.347519e-04 Well-being
## CA_GroupHigh_CA                          3.357599e-01 Well-being
## CA_GroupLow_CA                           7.447745e-06 Well-being
## scale(max_age_MHQ)                      1.715919e-111 Well-being
## Sex                                      2.734456e-03 Well-being
## I(scale(max_age_MHQ)^2)                  1.654583e-10 Well-being
## CA_GroupHigh_CA:scale(max_age_MHQ)       8.322183e-01 Well-being
## CA_GroupLow_CA:scale(max_age_MHQ)        2.399292e-04 Well-being
## CA_GroupHigh_CA:Sex                      1.856789e-02 Well-being
## CA_GroupLow_CA:Sex                       7.807090e-01 Well-being
## scale(max_age_MHQ):Sex                   1.199672e-25 Well-being
## CA_GroupHigh_CA:I(scale(max_age_MHQ)^2)  3.940790e-01 Well-being
## CA_GroupLow_CA:I(scale(max_age_MHQ)^2)   5.515500e-01 Well-being
## CA_GroupHigh_CA:scale(max_age_MHQ):Sex   6.649020e-01 Well-being
## CA_GroupLow_CA:scale(max_age_MHQ):Sex    5.921088e-01 Well-being
```

```
## `geom_smooth()` using formula 'y ~ x'
```

#### Age under median

```
## [1] "Median max age =  65"
```

```
##                                                               Estimate
## (Intercept)                                              -0.0249063606
## CA_GroupHigh_CA                                           0.0175026153
## CA_GroupLow_CA                                           -0.2265603563
## Sex                                                       0.0688789390
## scale(max_age_MHQ, scale = FALSE)                         0.0129550207
## I(scale((max_age_MHQ), scale = FALSE)^2)                  0.0008108164
## CA_GroupHigh_CA:Sex                                      -0.0464351426
## CA_GroupLow_CA:Sex                                        0.0296300479
## CA_GroupHigh_CA:scale(max_age_MHQ, scale = FALSE)         0.0005061278
## CA_GroupLow_CA:scale(max_age_MHQ, scale = FALSE)          0.0098426114
## CA_GroupHigh_CA:I(scale((max_age_MHQ), scale = FALSE)^2)  0.0001305262
## CA_GroupLow_CA:I(scale((max_age_MHQ), scale = FALSE)^2)   0.0012418134
##                                                                    SE
## (Intercept)                                              0.0062597060
## CA_GroupHigh_CA                                          0.0204373669
## CA_GroupLow_CA                                           0.0379318390
## Sex                                                      0.0086388550
## scale(max_age_MHQ, scale = FALSE)                        0.0008794176
## I(scale((max_age_MHQ), scale = FALSE)^2)                 0.0001785870
## CA_GroupHigh_CA:Sex                                      0.0274840984
## CA_GroupLow_CA:Sex                                       0.0526365675
## CA_GroupHigh_CA:scale(max_age_MHQ, scale = FALSE)        0.0028473113
## CA_GroupLow_CA:scale(max_age_MHQ, scale = FALSE)         0.0053620085
## CA_GroupHigh_CA:I(scale((max_age_MHQ), scale = FALSE)^2) 0.0005713422
## CA_GroupLow_CA:I(scale((max_age_MHQ), scale = FALSE)^2)  0.0010981322
##                                                                 t/z
## (Intercept)                                              -3.9788387
## CA_GroupHigh_CA                                           0.8564027
## CA_GroupLow_CA                                           -5.9728282
## Sex                                                       7.9731561
## scale(max_age_MHQ, scale = FALSE)                        14.7313647
## I(scale((max_age_MHQ), scale = FALSE)^2)                  4.5401772
## CA_GroupHigh_CA:Sex                                      -1.6895276
## CA_GroupLow_CA:Sex                                        0.5629176
## CA_GroupHigh_CA:scale(max_age_MHQ, scale = FALSE)         0.1777564
## CA_GroupLow_CA:scale(max_age_MHQ, scale = FALSE)          1.8356202
## CA_GroupHigh_CA:I(scale((max_age_MHQ), scale = FALSE)^2)  0.2284554
## CA_GroupLow_CA:I(scale((max_age_MHQ), scale = FALSE)^2)   1.1308415
##                                                                     p
## (Intercept)                                              6.932935e-05
## CA_GroupHigh_CA                                          3.917783e-01
## CA_GroupLow_CA                                           2.344087e-09
## Sex                                                      1.572241e-15
## scale(max_age_MHQ, scale = FALSE)                        4.884576e-49
## I(scale((max_age_MHQ), scale = FALSE)^2)                 5.630982e-06
## CA_GroupHigh_CA:Sex                                      9.112327e-02
## CA_GroupLow_CA:Sex                                       5.734930e-01
## CA_GroupHigh_CA:scale(max_age_MHQ, scale = FALSE)        8.589148e-01
## CA_GroupLow_CA:scale(max_age_MHQ, scale = FALSE)         6.641850e-02
## CA_GroupHigh_CA:I(scale((max_age_MHQ), scale = FALSE)^2) 8.192930e-01
## CA_GroupLow_CA:I(scale((max_age_MHQ), scale = FALSE)^2)  2.581261e-01
##                                                               Model
## (Intercept)                                              Well-being
## CA_GroupHigh_CA                                          Well-being
## CA_GroupLow_CA                                           Well-being
## Sex                                                      Well-being
## scale(max_age_MHQ, scale = FALSE)                        Well-being
## I(scale((max_age_MHQ), scale = FALSE)^2)                 Well-being
## CA_GroupHigh_CA:Sex                                      Well-being
## CA_GroupLow_CA:Sex                                       Well-being
## CA_GroupHigh_CA:scale(max_age_MHQ, scale = FALSE)        Well-being
## CA_GroupLow_CA:scale(max_age_MHQ, scale = FALSE)         Well-being
## CA_GroupHigh_CA:I(scale((max_age_MHQ), scale = FALSE)^2) Well-being
## CA_GroupLow_CA:I(scale((max_age_MHQ), scale = FALSE)^2)  Well-being
```

```
## `geom_smooth()` using formula 'y ~ x'
```

#### Age over median

```
## [1] "Median max age =  65"
```

```
##                                                               Estimate
## (Intercept)                                               0.0117226317
## CA_GroupHigh_CA                                           0.0156112614
## CA_GroupLow_CA                                           -0.0527064339
## Sex                                                      -0.0414844170
## scale(max_age_MHQ, scale = FALSE)                         0.0004186762
## I(scale((max_age_MHQ), scale = FALSE)^2)                 -0.0011500145
## CA_GroupHigh_CA:Sex                                      -0.0468037129
## CA_GroupLow_CA:Sex                                       -0.0149158392
## CA_GroupHigh_CA:scale(max_age_MHQ, scale = FALSE)         0.0041912220
## CA_GroupLow_CA:scale(max_age_MHQ, scale = FALSE)         -0.0096068919
## CA_GroupHigh_CA:I(scale((max_age_MHQ), scale = FALSE)^2)  0.0006815719
## CA_GroupLow_CA:I(scale((max_age_MHQ), scale = FALSE)^2)   0.0008995977
##                                                                    SE
## (Intercept)                                              0.0060661949
## CA_GroupHigh_CA                                          0.0208312196
## CA_GroupLow_CA                                           0.0373263926
## Sex                                                      0.0088118034
## scale(max_age_MHQ, scale = FALSE)                        0.0014737209
## I(scale((max_age_MHQ), scale = FALSE)^2)                 0.0004099812
## CA_GroupHigh_CA:Sex                                      0.0301085138
## CA_GroupLow_CA:Sex                                       0.0551070063
## CA_GroupHigh_CA:scale(max_age_MHQ, scale = FALSE)        0.0049045748
## CA_GroupLow_CA:scale(max_age_MHQ, scale = FALSE)         0.0091155642
## CA_GroupHigh_CA:I(scale((max_age_MHQ), scale = FALSE)^2) 0.0013427996
## CA_GroupLow_CA:I(scale((max_age_MHQ), scale = FALSE)^2)  0.0024860874
##                                                                 t/z
## (Intercept)                                               1.9324522
## CA_GroupHigh_CA                                           0.7494166
## CA_GroupLow_CA                                           -1.4120420
## Sex                                                      -4.7078237
## scale(max_age_MHQ, scale = FALSE)                         0.2840946
## I(scale((max_age_MHQ), scale = FALSE)^2)                 -2.8050421
## CA_GroupHigh_CA:Sex                                      -1.5545009
## CA_GroupLow_CA:Sex                                       -0.2706705
## CA_GroupHigh_CA:scale(max_age_MHQ, scale = FALSE)         0.8545536
## CA_GroupLow_CA:scale(max_age_MHQ, scale = FALSE)         -1.0538999
## CA_GroupHigh_CA:I(scale((max_age_MHQ), scale = FALSE)^2)  0.5075753
## CA_GroupLow_CA:I(scale((max_age_MHQ), scale = FALSE)^2)   0.3618528
##                                                                     p
## (Intercept)                                              5.330855e-02
## CA_GroupHigh_CA                                          4.536092e-01
## CA_GroupLow_CA                                           1.579429e-01
## Sex                                                      2.509499e-06
## scale(max_age_MHQ, scale = FALSE)                        7.763389e-01
## I(scale((max_age_MHQ), scale = FALSE)^2)                 5.032665e-03
## CA_GroupHigh_CA:Sex                                      1.200704e-01
## CA_GroupLow_CA:Sex                                       7.866454e-01
## CA_GroupHigh_CA:scale(max_age_MHQ, scale = FALSE)        3.928018e-01
## CA_GroupLow_CA:scale(max_age_MHQ, scale = FALSE)         2.919331e-01
## CA_GroupHigh_CA:I(scale((max_age_MHQ), scale = FALSE)^2) 6.117531e-01
## CA_GroupLow_CA:I(scale((max_age_MHQ), scale = FALSE)^2)  7.174633e-01
##                                                               Model
## (Intercept)                                              Well-being
## CA_GroupHigh_CA                                          Well-being
## CA_GroupLow_CA                                           Well-being
## Sex                                                      Well-being
## scale(max_age_MHQ, scale = FALSE)                        Well-being
## I(scale((max_age_MHQ), scale = FALSE)^2)                 Well-being
## CA_GroupHigh_CA:Sex                                      Well-being
## CA_GroupLow_CA:Sex                                       Well-being
## CA_GroupHigh_CA:scale(max_age_MHQ, scale = FALSE)        Well-being
## CA_GroupLow_CA:scale(max_age_MHQ, scale = FALSE)         Well-being
## CA_GroupHigh_CA:I(scale((max_age_MHQ), scale = FALSE)^2) Well-being
## CA_GroupLow_CA:I(scale((max_age_MHQ), scale = FALSE)^2)  Well-being
```

```
## `geom_smooth()` using formula 'y ~ x'
```

#### Average CA Results

```
##                            Estimate          SE        t/z             p
## (Intercept)              0.01519169 0.004296898   3.535503  4.071696e-04
## scale(max_age_MHQ)       0.07140002 0.003194698  22.349540 2.172925e-110
## Sex                      0.01822676 0.006159386   2.959184  3.085215e-03
## I(scale(max_age_MHQ)^2) -0.01910673 0.002994769  -6.380034  1.777614e-10
## scale(max_age_MHQ):Sex  -0.06428339 0.006149491 -10.453449  1.453161e-25
##                              Model
## (Intercept)             Well-being
## scale(max_age_MHQ)      Well-being
## Sex                     Well-being
## I(scale(max_age_MHQ)^2) Well-being
## scale(max_age_MHQ):Sex  Well-being
```

```
## [1] "beta =  0.0714000224485863 SE = 0.00319469763176148 OR = 1.07401076836668 p =  2.1729246786779e-110"
```

#### Low CA Results

```
##                            Estimate         SE        t/z            p
## (Intercept)              0.02359570 0.02618383  0.9011557 3.675786e-01
## scale(max_age_MHQ)       0.12393633 0.01903355  6.5114684 8.706314e-11
## Sex                      0.02448490 0.03729585  0.6565047 5.115503e-01
## I(scale(max_age_MHQ)^2) -0.02675916 0.01817421 -1.4723698 1.410270e-01
## scale(max_age_MHQ):Sex  -0.03933544 0.03683905 -1.0677648 2.857134e-01
##                              Model
## (Intercept)             Well-being
## scale(max_age_MHQ)      Well-being
## Sex                     Well-being
## I(scale(max_age_MHQ)^2) Well-being
## scale(max_age_MHQ):Sex  Well-being
```

```
## [1] "beta =  0.12393633373694 SE = 0.019033546180348 OR = 1.13194380207315 p =  8.70631384130346e-11"
```
